# Associations between cannabis use, tobacco use and co-use with brain volume: a systematic review and meta-analysis

**DOI:** 10.1101/2025.07.21.25331928

**Authors:** Katherine Sawyer, Tom P Freeman, Martine Skumlien, Esther Walton, Thomas Lancaster, Jorien L Treur, Valentina Lorenzetti, Anna K M Blackwell, Chloe Burke, Richie J Carr, Constantinos Constantinides, Maisie Cox, Sarah Dance, Shadi Daryan, Sorcha Hamilton, Tom A. Jenkins, Gemma M. J. Taylor

**Affiliations:** Addiction and Mental Health Group, University of Bath; Department of Addictions, King’s College London; Department of Psychology, University of Bath; Department of Psychiatry, Amsterdam UMC, location University of Amsterdam; Neuroscience of Addiction and Mental Health Program, Healthy Brain and Mind Research Centre, Australian Catholic University; University of Bath; University of Bristol; University of Bath; Addiction and Mental Health Group, Centre for Motivation and Behaviour Change, University of Bath; Addiction and Mental Health Group, Bath Centre for Mindfulness and Community, University of Bath; Centre for Public Health Research, Population Health Sciences, Bristol Medical School, University of Bristol; Addiction and Mental Health Group, University of Bath; NIHR Oxford Biomedical Research Centre, John Radcliffe Hospital, Oxford OX3 9DU, United Kingdom Oxford Biomedical Research Centre, John Radcliffe Hospital, Oxford OX3 9DU, United Kingdom

**Author notes:** **Corresponding Author:** Katherine Sawyer, Department of Psychology, 10 West, 2.10, University of Bath, Bath, BA2 7AY.

**Keywords:** Cannabis, Tobacco, Co-use, Brain volume, Neuroimaging, MRI, Systematic review, Meta-analysis

## Abstract

**Background and Aims:** Cannabis is the most widely used illicit drug worldwide and is often co-used with tobacco, the leading cause of preventable death. Although cannabis and tobacco have distinct neurobiological actions, their associations with brain volumes are unclear. We aimed to systematically review brain volumes associated with cannabis use, tobacco use, and their co-use.

**Design:** Systematic review and meta-analysis (CRD42022356982).

**Setting:** SCOPUS, PubMed and PsycINFO were searched up to 5 September 2024

**Participants:** Searches yielded 103 studies: 57 investigated cannabis use, 45 investigated tobacco use, and one investigated tobacco and cannabis co-use.

**Measurements:** We extracted adjusted and unadjusted estimates. Random effects meta-analyses were stratified by exposure and study design across 33 brain regions. Risk of bias was assessed using a modified version of the Newcastle-Ottawa scale.

**Findings:** Meta-analysis of adjusted estimates from cross-sectional studies indicated smaller amygdala volumes (k = 17, g = 0.13, 95%CI [0.03, 0.23]) in people who use cannabis compared to controls. Relative to controls, people who smoked tobacco had smaller volumes in the amygdala (k = 5, g = 0.17, 95%CI [0.04, 0.31]), insula (k = 5, g = 0.17, 95%CI [0.06, 0.27]), pallidum (k = 5, g = 0.17, 95%CI [0.13, 0.21]) and total grey matter volume (TGMV) (k = 7, g = 0.17, 95%CI [0.04, 0.30]). Longitudinal studies indicated a larger decrease in TGMV in people who smoke tobacco (k = 5, g = 0.05, 95%CI [0.01, 0.10]) relative to controls.

**Conclusions:** There was evidence that cannabis use was associated with smaller volume in the amygdala. Tobacco use was associated with smaller amygdala, insula, pallidum and total grey matter volume.

## INTRODUCTION

Cannabis and tobacco use are highly prevalent and associated with negative health outcomes. Cannabis alone was used by approximately 228 million people globally in 2022, around 4.4% of the global adult population (1). Cannabis use is increasing in prevalence and potency (2,3). Cannabis products with higher potency carry a greater risk of mental-ill health and addiction compared to lower potency products (4). Meanwhile, tobacco was used by approximately 1.18 billion people in 2020, around 30% of the global population (5). Over 8 million deaths a year are attributed to tobacco smoking (6,7). Tobacco smoking also exacerbates health inequalities, leading to higher levels of harm in more deprived and vulnerable populations (8). Cannabis can be co-used with tobacco, either separately within the same time period (e.g., same day, cigarettes and cannabis separately), or co-administered, where both tobacco and cannabis are delivered together (e.g., through ‘spliffs’ or ‘joints’ in European terms) (9).

Tobacco and cannabis act on distinct pathways in the brain. When cannabis is consumed, tetrahydrocannabinol (THC, the primary psychoactive component of cannabis) acts on cannabinoid type one (CB1) receptors, which are found at high levels in the hippocampus, basal ganglia and cerebellum and moderate levels in the prefrontal cortex, amygdala, and hypothalamus (10,11). Nicotine, the main psychoactive component of tobacco, acts on nicotinic acetylcholine receptors (nAChRs) found throughout the brain, including the hippocampus, ventral tegmental area, nucleus accumbens, prefrontal cortex and amygdala (12–14).

Despite high rates of co-use and similar associations with grey matter volume, previous systematic reviews have focused on brain volume and either cannabis use (15–18) or tobacco use individually (19–22). A recent umbrella review of meta-analyses of Voxel Based Morphometry (VBM) tobacco studies found consistently smaller volumes in the prefrontal cortex, insula and cingulate cortex in people who smoke tobacco chronically, compared to controls (23). Meta-analyses of structural neuroimaging cannabis studies have found smaller volumes of the hippocampus, lateral and medial OFC (15) and cerebellum (17) in people who regularly use cannabis compared to controls. However, no significant differences were found in adolescents (18). Only one narrative review has synthesised the evidence on the association between cannabis and tobacco co-use on brain structure (24), and identified one study reporting smaller hippocampal volume for co-users of nicotine and cannabis, compared to controls and nicotine only users (25).

Mendelian randomisation (MR) is a method which is increasingly being used to test causal relationships. MR uses genetic variation that predicts modifiable exposures as instrumental variables, thereby accounting for unobserved confounding and reverse causation (26). To date, there are no systematic reviews of Mendelian randomisation studies examining the associations between tobacco or cannabis use and brain structure, nor any that integrate these findings with evidence from observational studies.

Evidence from both individual studies and reviews generally suggests an association between cannabis and tobacco use and reduced grey matter volume; however, findings regarding specific brain regions remain heterogeneous and inconsistent. Previous reviews focussed on only cross-sectional studies which reduce potential for causal inference. Finally, despite high rates of co-use, there is still limited investigation of studies of cannabis and tobacco co-use, or even comparisons of results between cannabis and tobacco studies using similar meta-analysis methods and exposure definitions. Therefore, we present, to our knowledge, the first systematic review and meta-analysis of the associations between cannabis use, tobacco use, and/or co-use with brain volume, across different study designs.

### Aims

In this systematic review and meta-analysis, we aimed to synthesise the literature examining brain volumetric differences between people who use cannabis and/or tobacco compared to non-using controls, across cross-sectional, longitudinal and MR study designs.

## METHODS

This review was preregistered on PROSPERO (CRD42022356982) and was reported according to the Preferred Reporting Items for Systematic Reviews and Meta-Analyses (PRISMA) statement (27)(supplementary file 1, table 1).

**Table 1:**
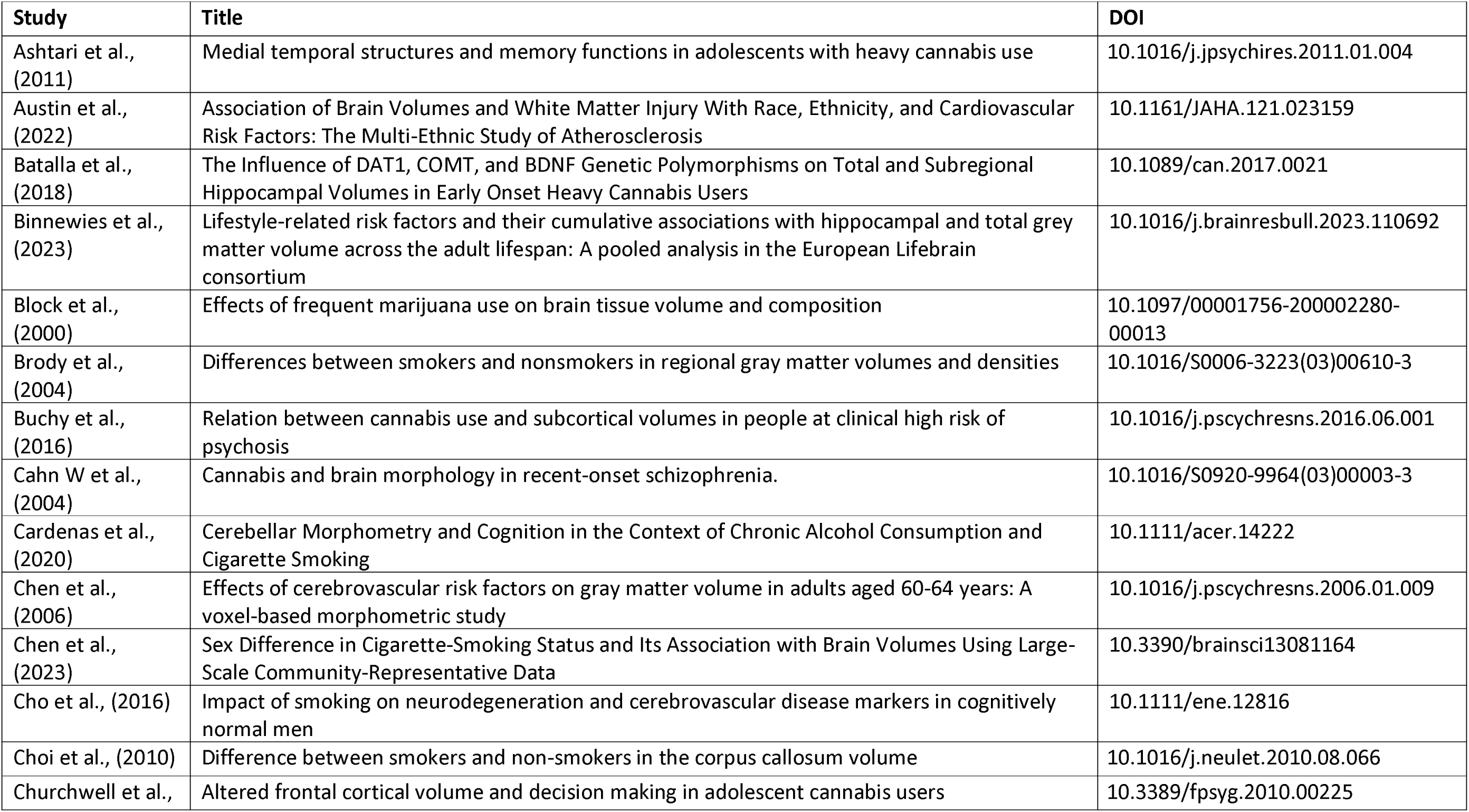
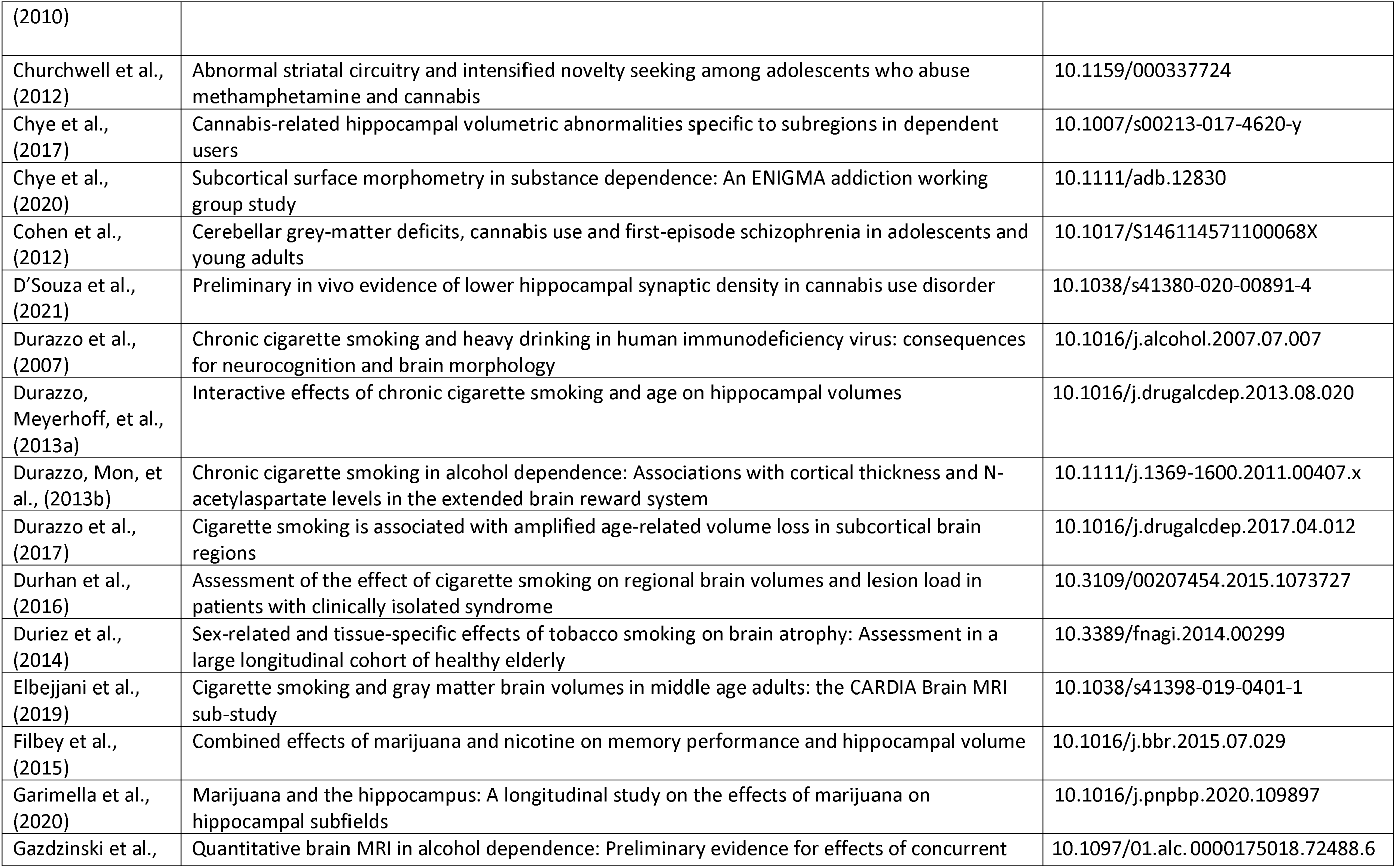
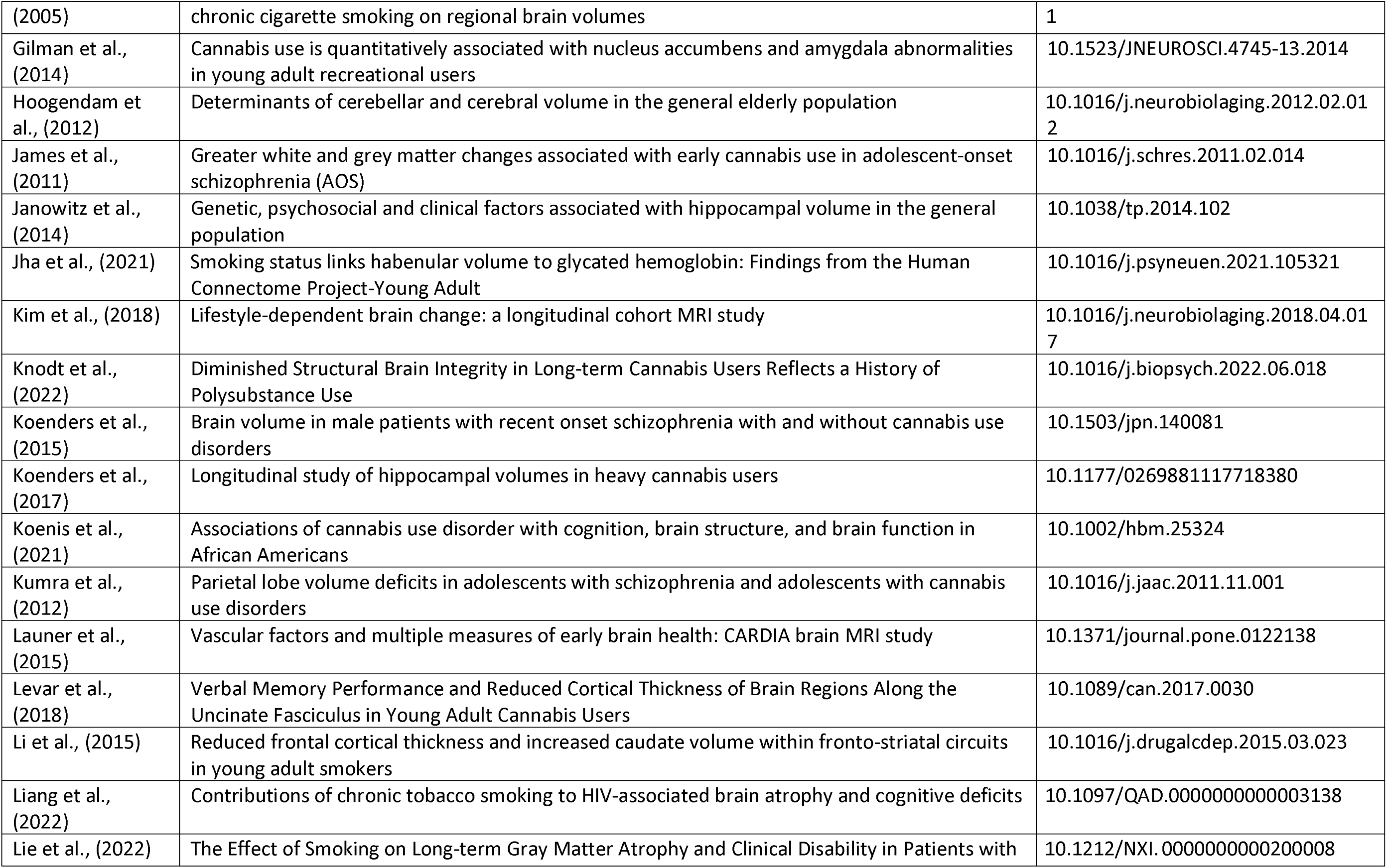
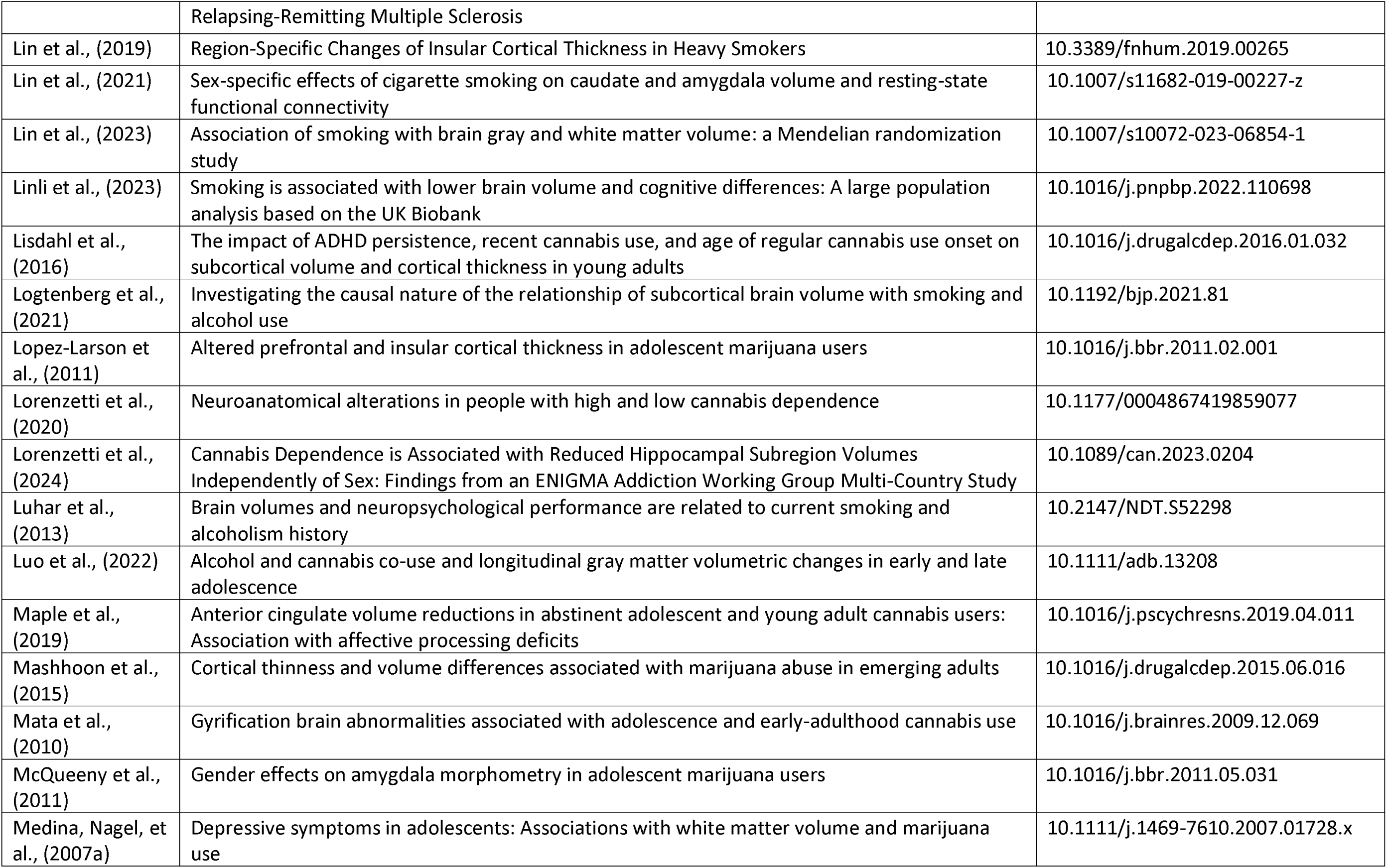
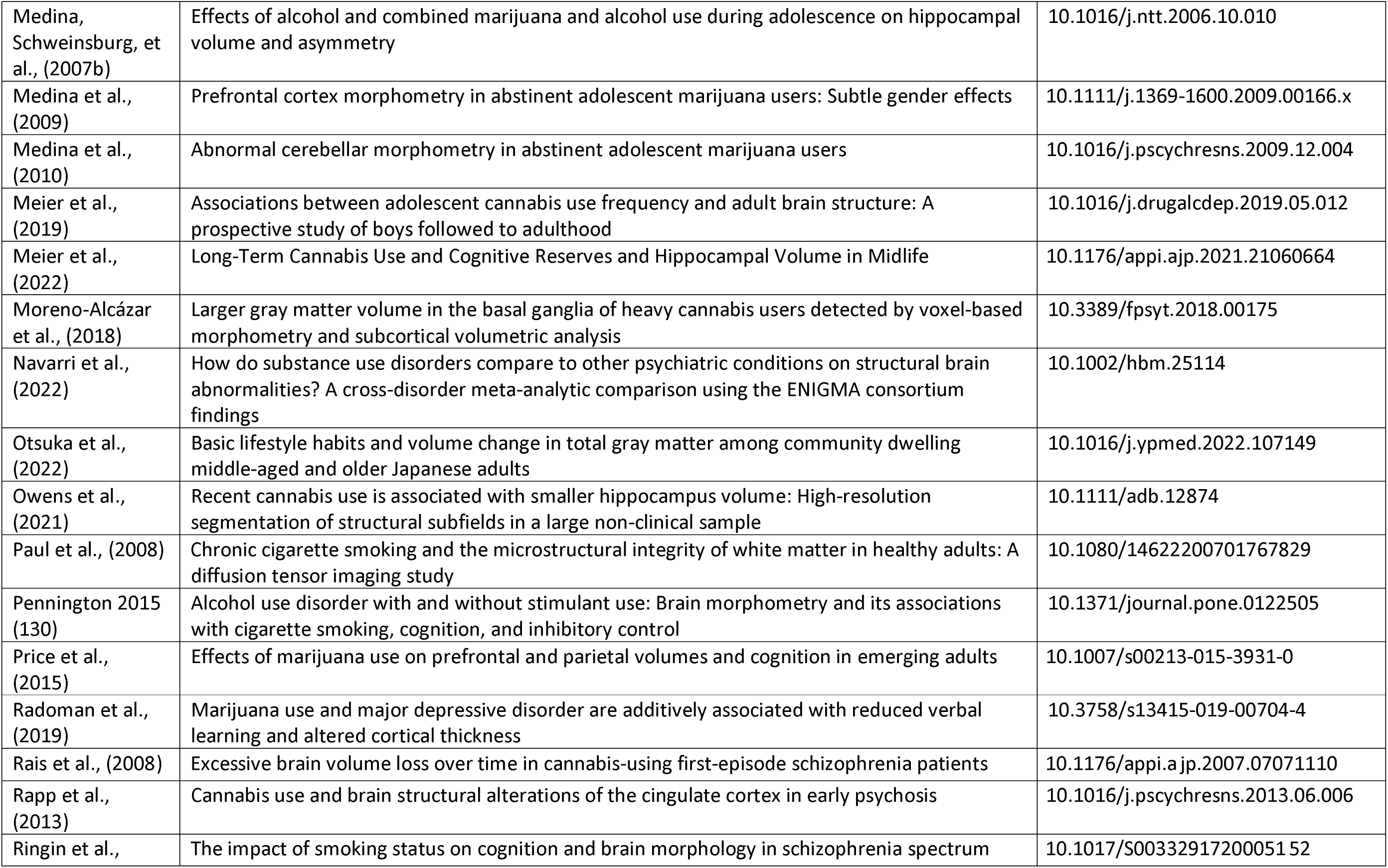
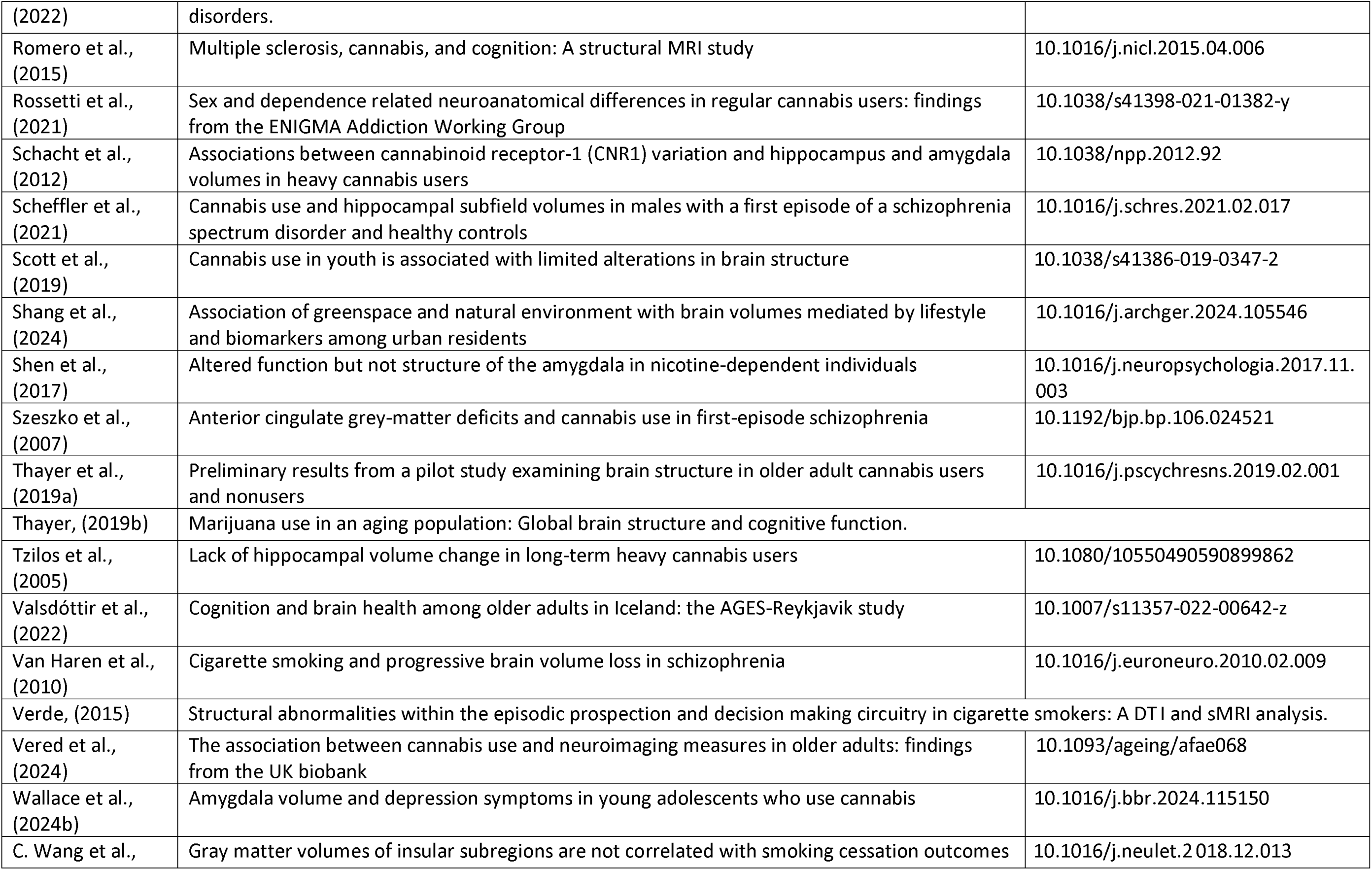
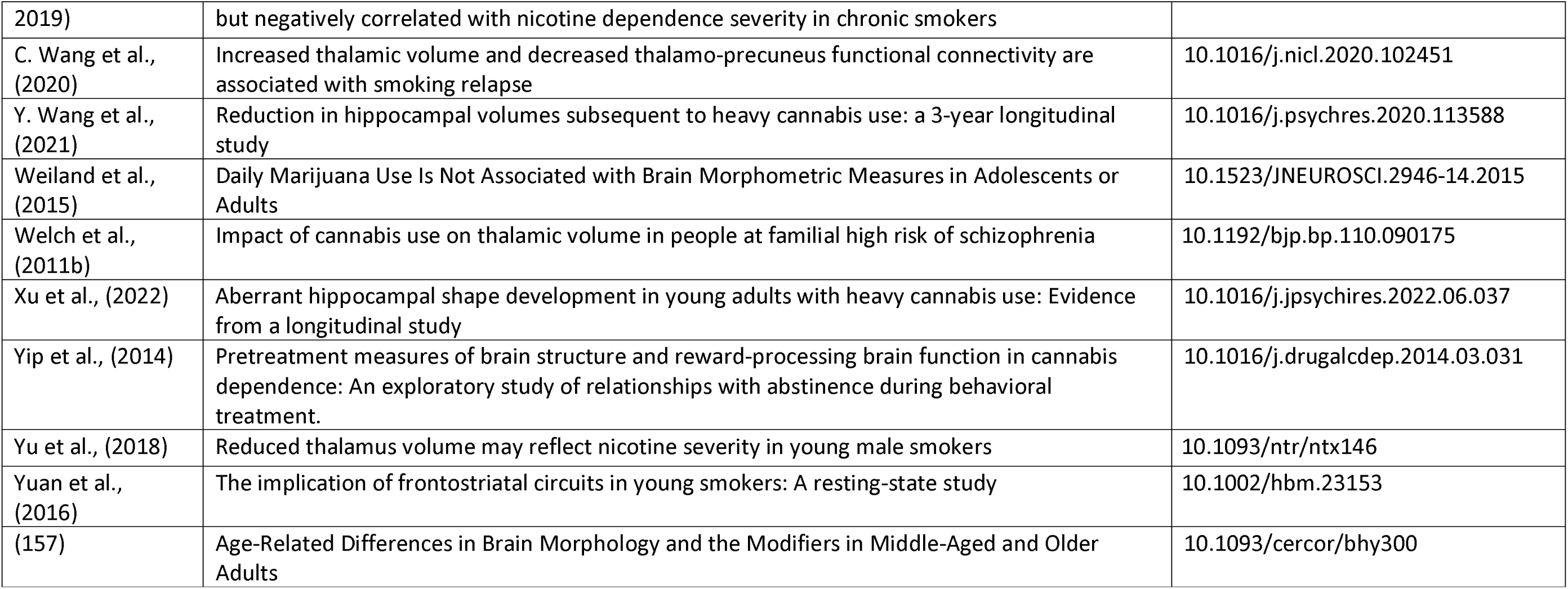
Studies included in the review.

### Eligibility criteria

Studies were included if they met the following criteria: (1) journal article, conference abstract or dissertation; (2) human participants; (3) measured brain volume using T1-weighted structural MRI; (4) primary use of current tobacco use, cannabis use and/or co-use and a comparator group which was non-exposed (as per study definition); (5) observational studies (cross-sectional or longitudinal), or instrumental variable studies such as Mendelian randomisation studies.

Exclusion criteria were: (1) primary substance of use was not cannabis or tobacco (but use of other substances was allowed); (2) pre/post-natal tobacco/cannabis exposure; (3) studies which reported only voxel-wise results; (4) studies where the control group was low frequency of use or absence of cannabis or tobacco abuse, dependence, or use disorder. There were no restrictions on date or language, age, psychiatric diagnosis, or population type.

Volume can be estimated by either voxel-based techniques (i.e., voxel-based morphometry analysing voxel by voxel differences) or summary representations of specific structures within the brain (28). Here we included summary representations of whole regions only to minimise methodological heterogeneity and support meta-analysis.

### Information sources and study selection

One author (KS) searched SCOPUS, PubMed and PsycINFO on 14^th^ September 2022 for relevant research articles and updated this list on 5th September 2024 (supplementary file 1, tables 2-4 for search strategy). Further cross-referencing from searching relevant systematic reviews in the field (15,18) was conducted. Studies in non-English language were translated using Google translate. All title/abstracts and full texts were independently screened by two authors and any discrepancies were resolved by discussion or, if necessary, contacting a third author. KS screened all studies, with second screening by either AB, CB, RC, CC, S Dance, S Daryan or TJ.

**Table 2:**
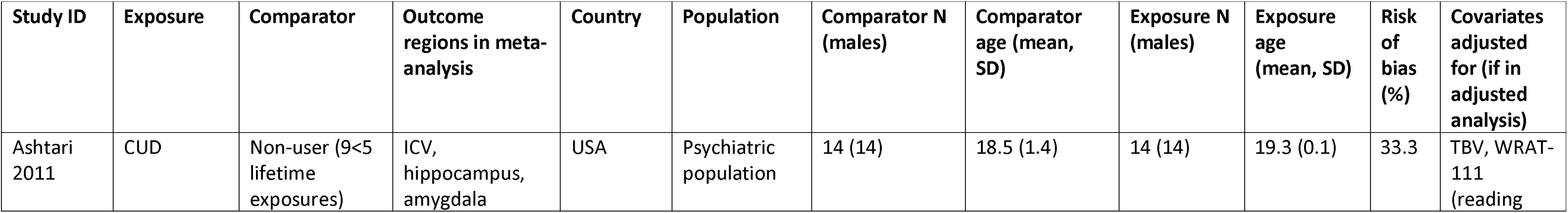
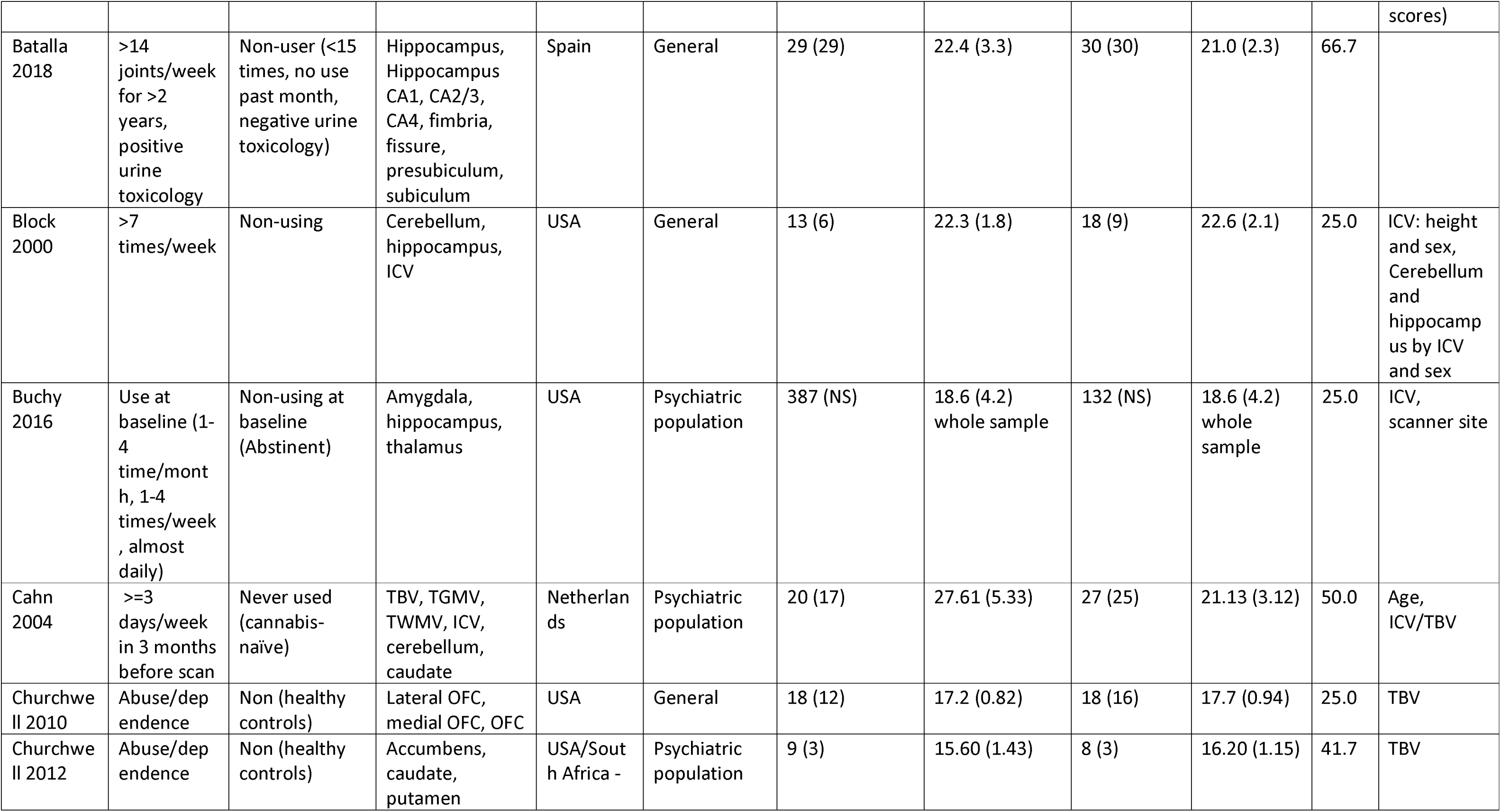
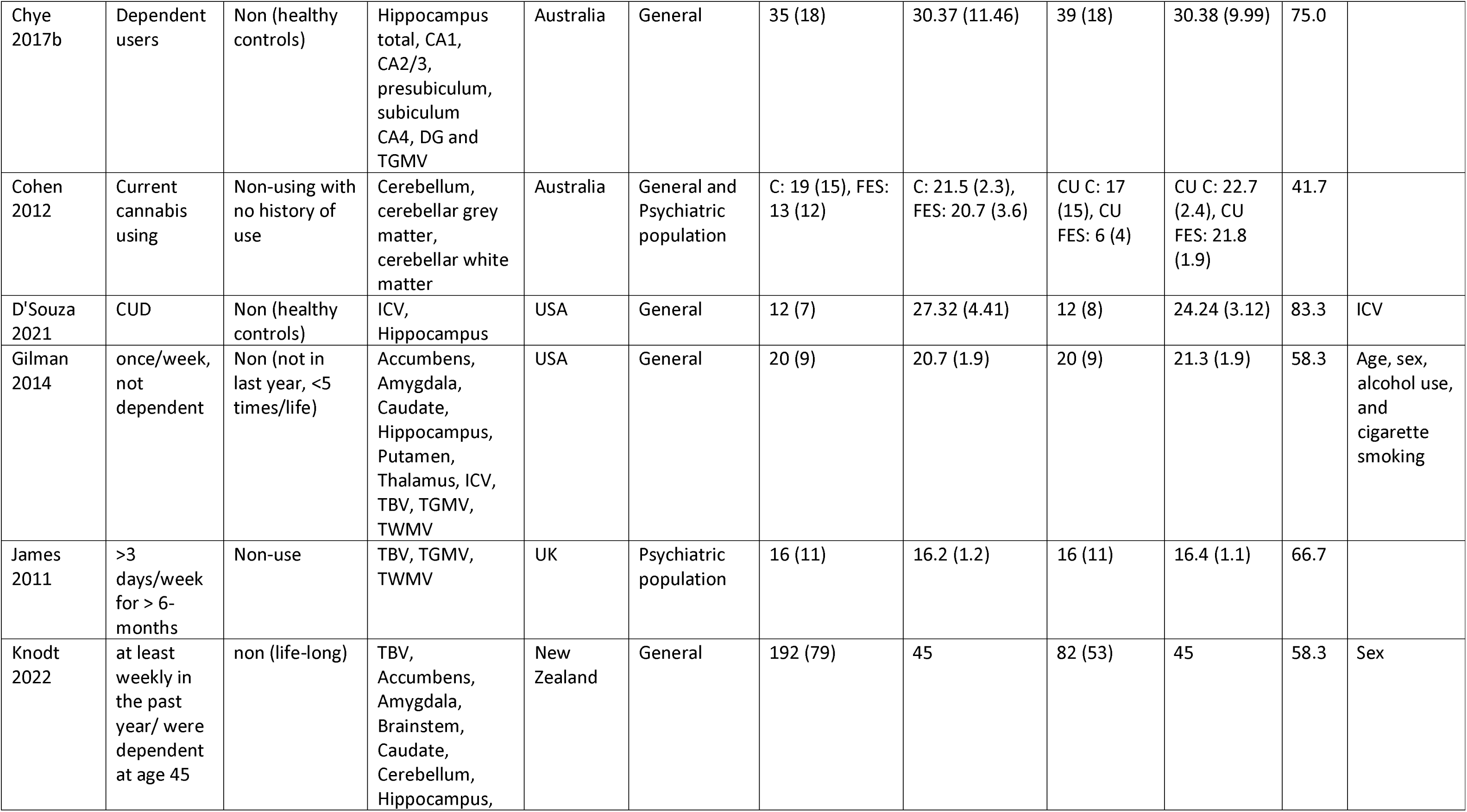
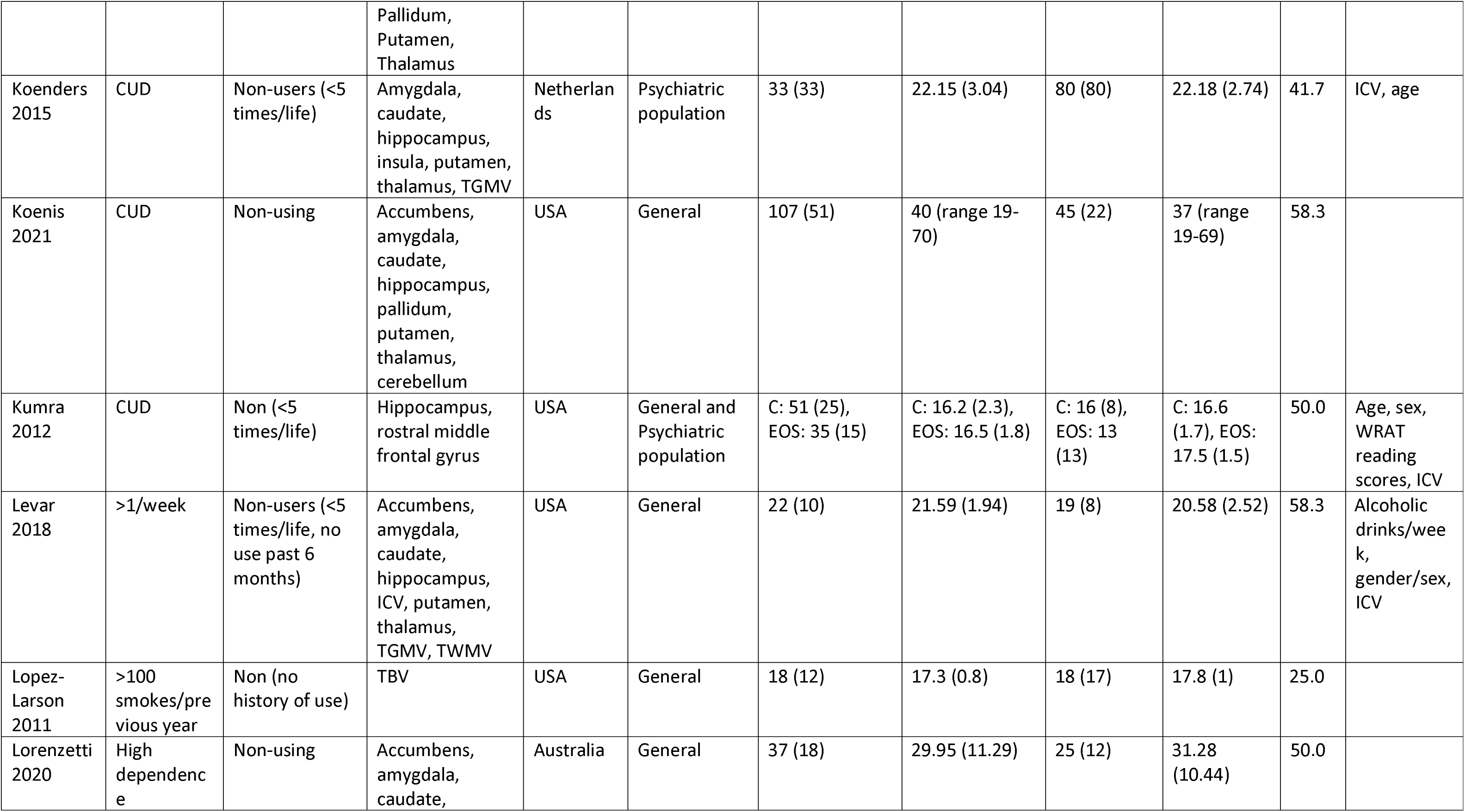
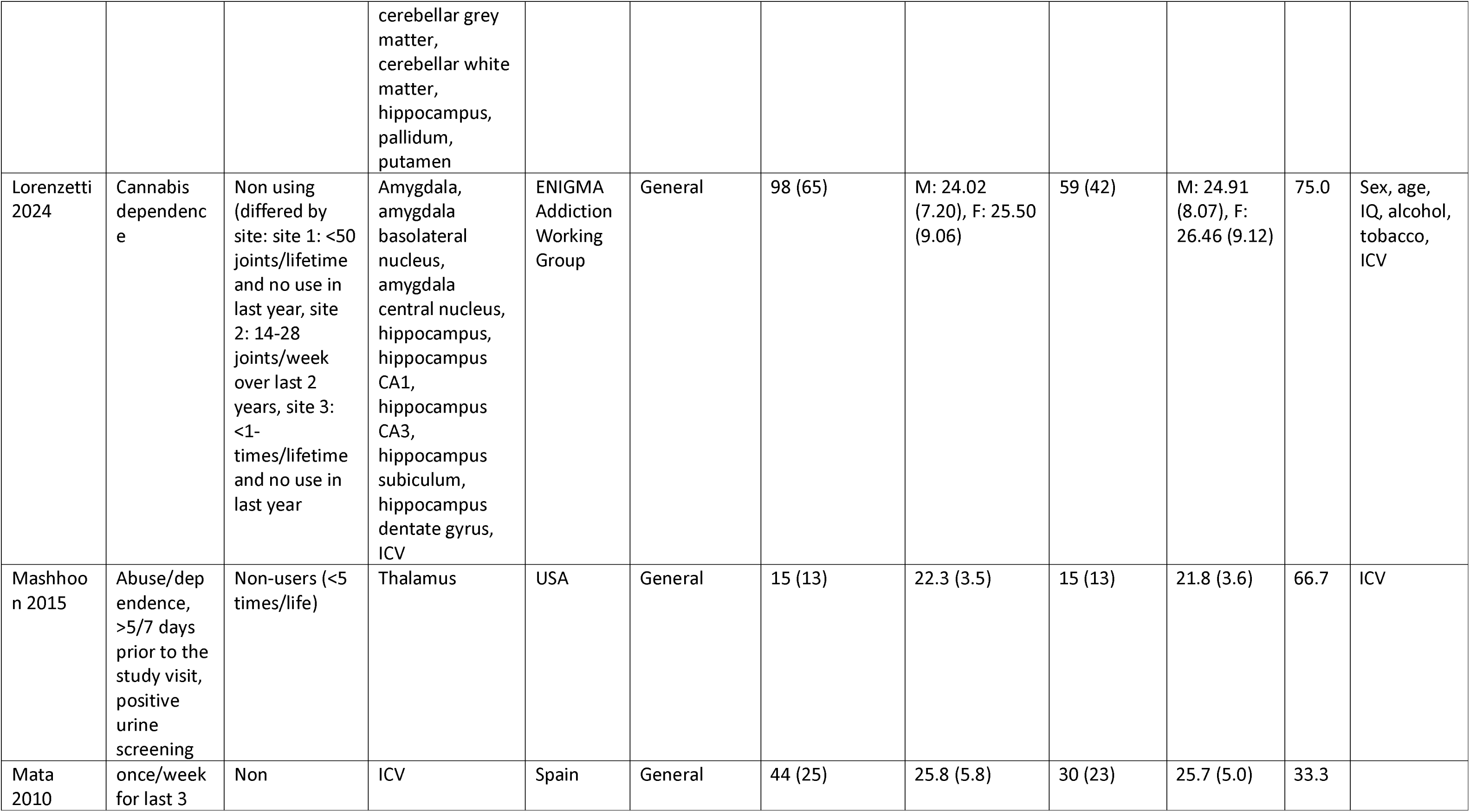
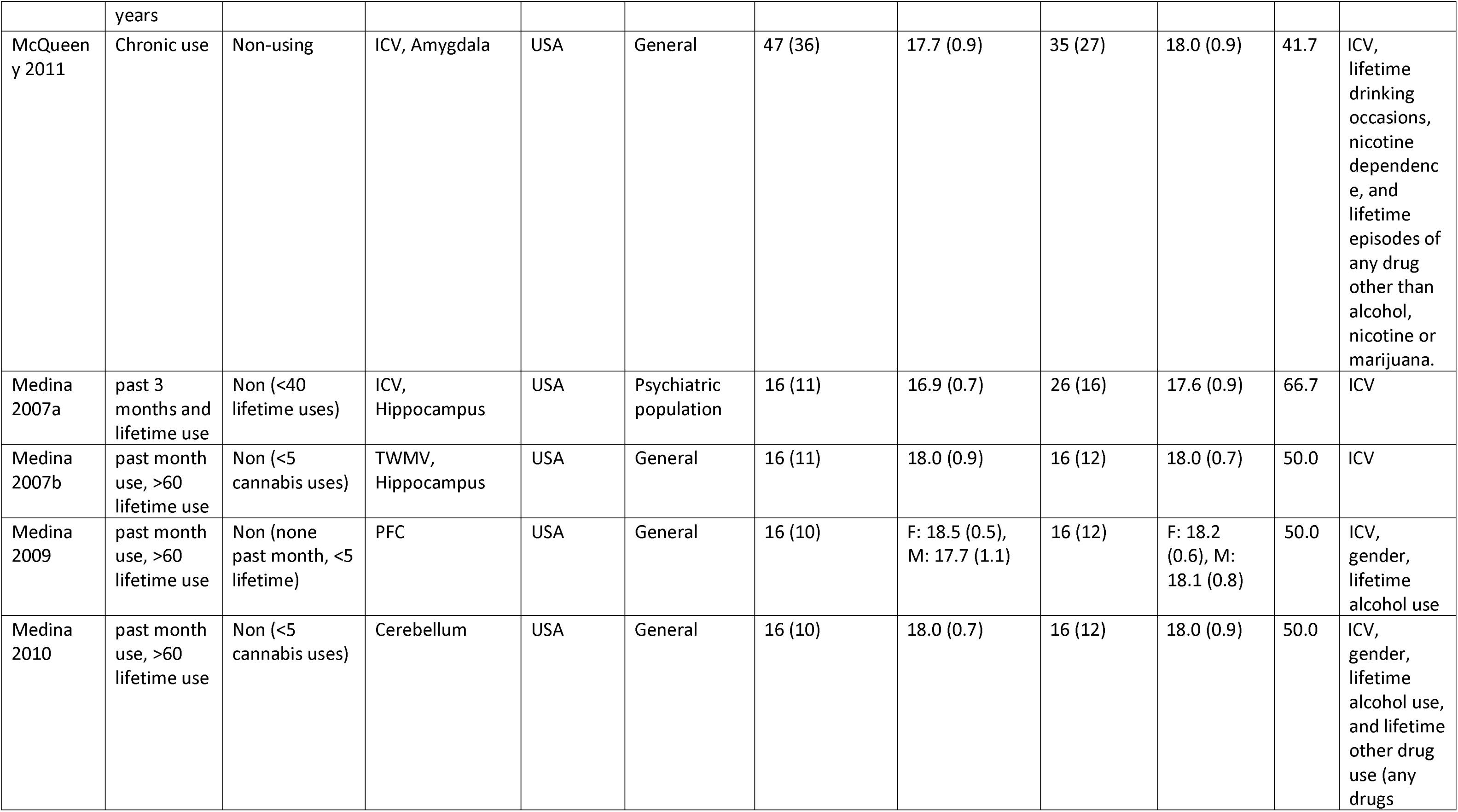
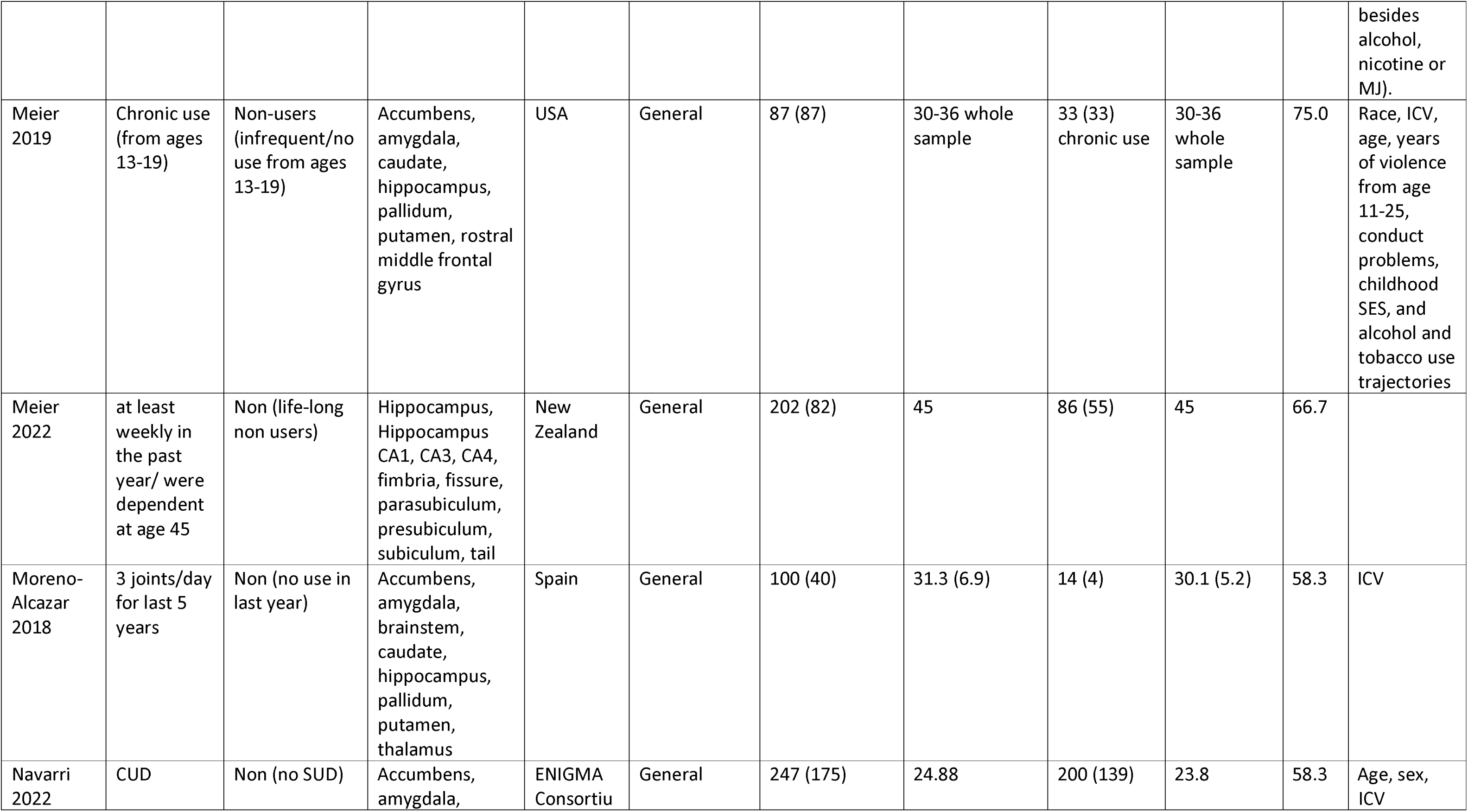
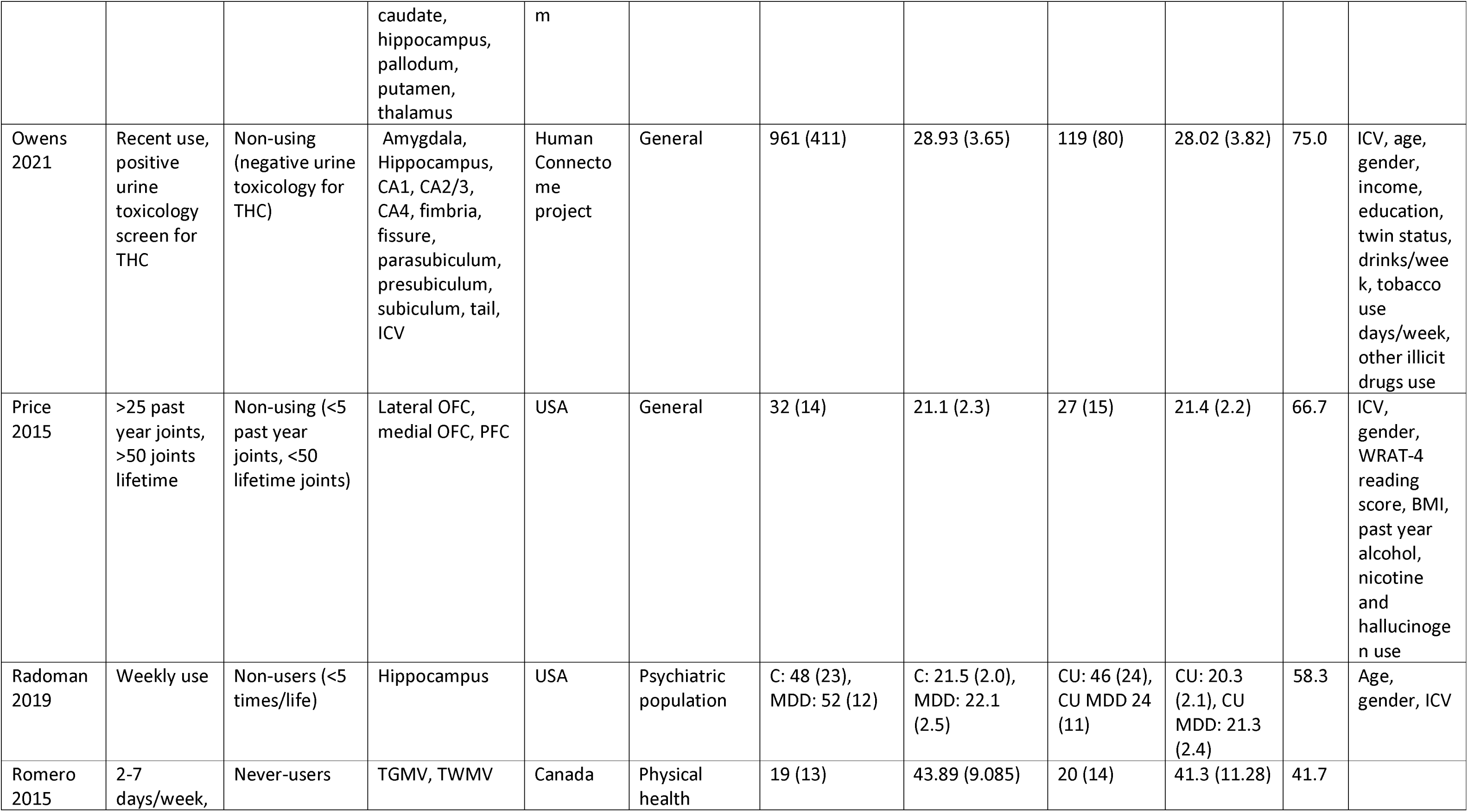
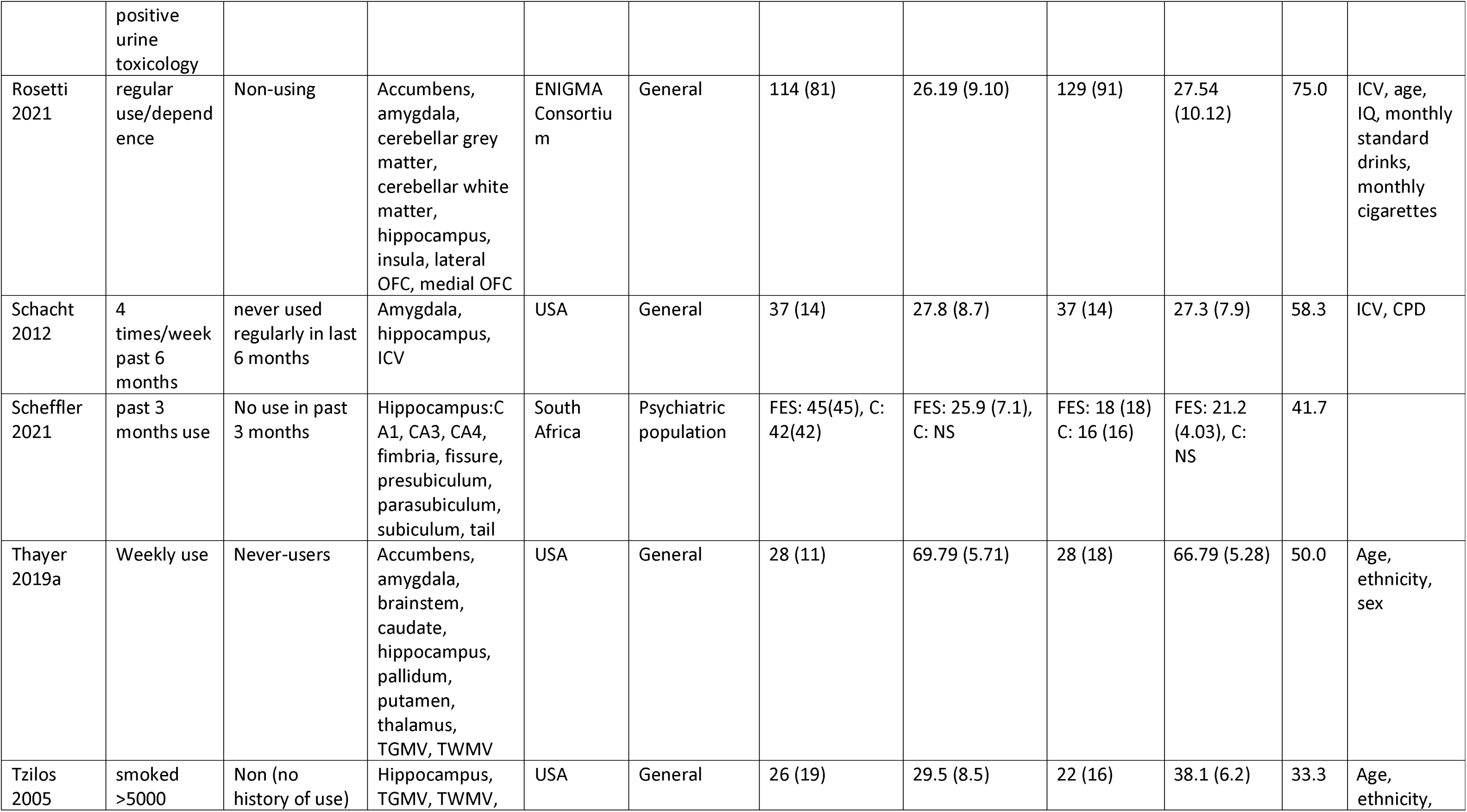
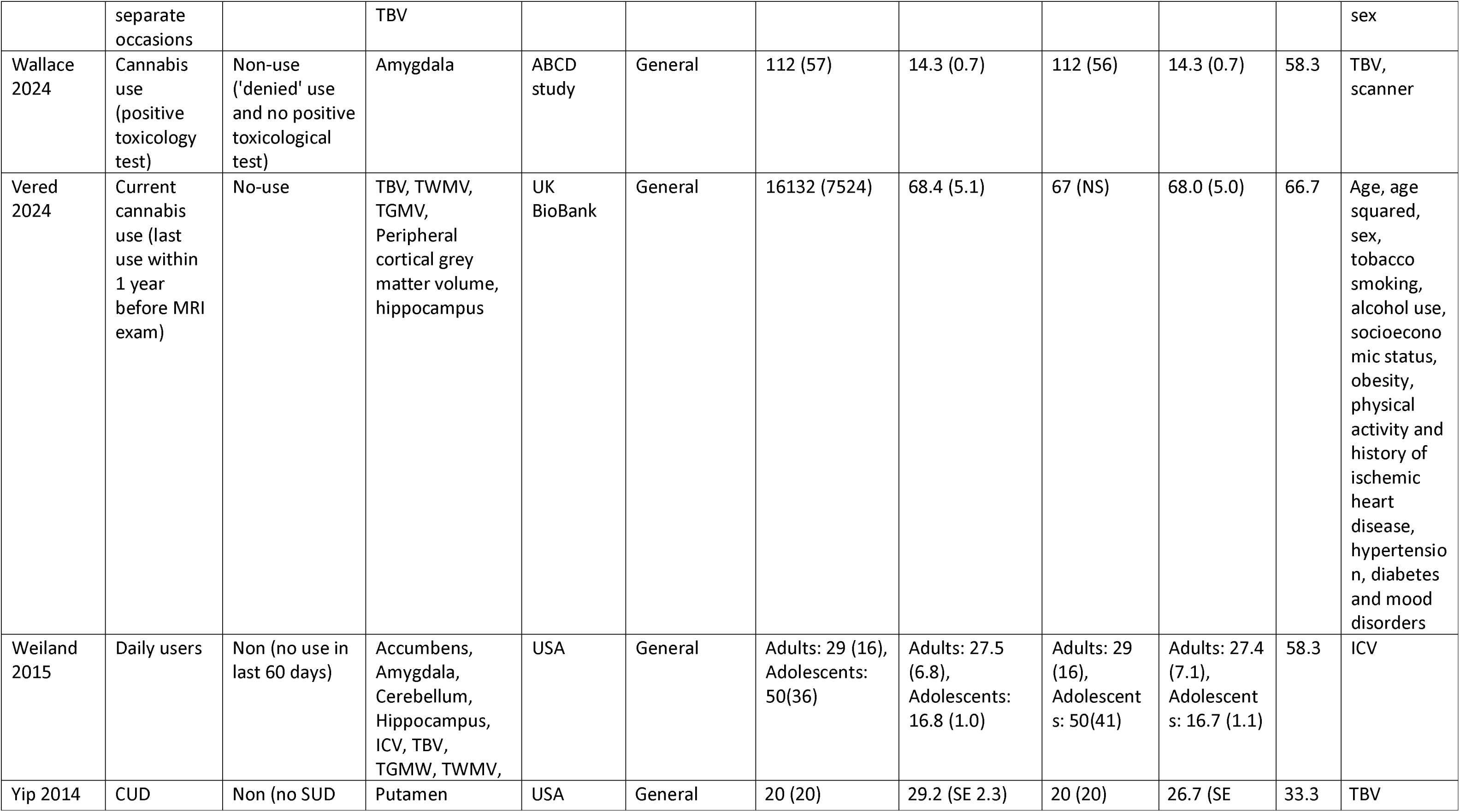
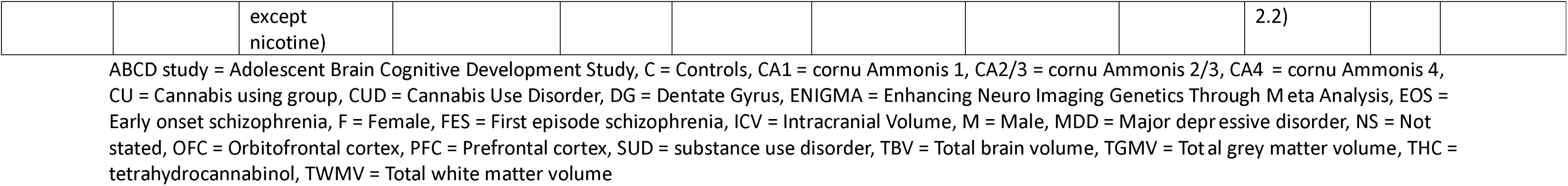
Characteristics of cross-sectional cannabis studies included in meta-analysis.

**Table 3:** Characteristics of cross-sectional tobacco studies included in meta-analysis.

| Study ID | Exposure | Comparator | Outcome regions in meta-analysis | Country | Population | Comparator N (males) | Comparator age (mean, SD) | Exposure N (males) | Exposure age (mean, SD) | Risk of bias (%) | Covariates adjusted for (if in adjusted analysis) |
| --- | --- | --- | --- | --- | --- | --- | --- | --- | --- | --- | --- |
| Austin 2022 | Current smokers | Never smokers | TBV, TGMV, TWMV | USA | Physical health | 493(NS) | 72 (8) whole sample | 60 (NS) | 72 (8) whole sample | 63.6 | Age, sex, and MESA site; total intracranial volume, BMI, smoking status, systolic and diastolic blood pressure, use of hypertension medication, high-density lipoprotein cholesterol, low-density lipoprotein cholesterol, diabetes status, and estimated glomerular filtration rate |
| Binnewies 2023 | Smoking (current smoking) | No current smoking | Hippocampus, TGMV | European Life-brain Consortium | General | 2727 (1425) | NS | 333 (160) | NS | 54.5 | Age, sex, education, scanner and ICV |
| Brody 2004 | >20 CPD and DSM nicotine dependence | <5 cig in a lifetime | Thalamus | USA | General | 17 (10) | 37.9 (12.9) | 19 (11) | 39.5 (10.3) | 72.7 | Age, gender |
| Cardenas 2020 | >10 CPD, past 5 years | never or <40 cig in lifetime | Cerebellum, ICV | USA | Psychiatric population | C: 17(14), ALC: 21(16) | C: 48(12) , ALC: 51(12) | C: 31(27), ALC: 23(21) | C: 49(9), ALC: 49(7) | 63.6 |  |
| Chen 2023 | Active smoking in last 2 weeks | Never <100 cigs/life | Amygdala, insula, hippocampus, thalamus, caudate, putamen,<br>pallidum,<br>nucleus<br>accumbens, ICV | USA, Dallas | General | 1106 (387) | F: 48.91 (10.97), M: 48.64 (10.19) | 402 (188) | F: 49.20 (9.33), M: 48.08 (9.95) | 54.5 | Age, annual alcohol consumption, ICV |
| Cho 2016 | Current smokers | Never smoker | ICV | Korea | General | 270 (270) | 66.1 (7.1) | 128 (128) | 62.7 (7.3) | 27.3 |  |
| Choi 2010 | Current smoking (yes/no) | Not currently smoking (yes/no) | Corpus callosum, ICV | Korea | General | 28 (28) | 35.49 (13.11) | 30 (30) | 32.82 (14.12) | 36.4 |  |
| Chye 2020 | Nicotine dependence | Non-smokers - do not use nicotine | ICV | ENIGMA Consortium | General | 918 (561) | 29.7(9.6) | 565(325) | 31.1(9.9) | 81.8 |  |
| Durazzo 2007a | Smoking every day or at least once/day for 6 months prior to study | No use 6 months prior to enrolment | Frontal GM, Parietal GM, Temporal GM | USA | Physical health | 17 (14.96) | 44.4 (4.9) | 27 (24.03) | 43.8 (5.4) | 36.4 | Age |
| Durazzo 2013a | Chronic smoking, at least 10CPD for 5 years or more | Never smoked, or <40 lifetimes cigarettes, no use in 10 years prior to study | Hippocampus, CA1, CA2/3, CA4-DG, subiculum | USA | General | 43 (35) | 46.5 (11) | 39 (33) | 43.3 (12.6) | 72.7 | ICV, education, average lifetime drinks/month |
| Durazzo 2013b | 'smokers' undefined | lifetime non-smoking | ICV | USA | Psychiatric population | 33 (NS) | 52.0 (9.7) | 43 (NS) | 50.1 (8.9) | 45.5 |  |
| Durazzo 2017 | 10CPD, past 5 years | Never or <40 cig in lifetime | Cerebellar cortex, thalamus | USA | General | 43 (34.83) | 43 (13) | 40 (34.80) | 47 (11) | 72.7 | BDI, average lifetime drinks/month |
| Durhan 2016 | Current smokers (undefined) | <5 cig in a lifetime | Cerebellar cortex, thalamus, ICV | Turkey | Physical health | C: 14 (4), CIS: 17 (4) | C: 29.8 (7.9), CIS: 29.7 (6.9) | C: 13 (7), CIS: 16 (8) | C: 33.7 (7.4), CIS: 34.3 (8.9) | 18.2 |  |
| Duriez 2014 - cross-sectional | Current smoker | Never smoker | Hippocampus, ICV, TBV, TGMV, TWMV | France | General | 920 (162) | M: 72.3 (4), F: 72.7 (4.06) whole sample | 83 (49) | M: 72.3 (4), F: 72.7 (4.06) whole sample | 54.5 |  |
| Elbejjani 2019 | >5 cigs/week almost every week, for at least 3 months | Never regular use | Accumbens, amygdala, caudate, frontal GM, hippocampus, parietal GM, putamen, temporal GM, thalamus, TBV, TGMV, TWMV | USA | General | 362 (173) | 50.18 (3.61) | 117 (60) | 49.63 (3.57) | 81.8 | Age, race, educational attainment, total intracranial volume, study centre, substance use and psychological composite factor |
| Gazdzinski 2005 | <1/2 times/week in the past 6 months | No use for at least one year before enrolment | Caudate, cerebellum, frontal GM, parietal GM, temporal GM, thalamus, ICV | USA | Psychiatric population | nsLD: 23 (100%); nsALC: 13 (100%) | nsLD: 47.5 (6.0); nsALC: 49.8 (9.7) | sLD: 7 (100%); sALC: 24 (100%) | sLD: 38.1 (8.7); sALC: 49.4 (8.3) | 63.6 | Age |
| Janowitz 2014 | >15 CPD | Never smokers | Hippocampus | Germany | General | 991 (NS) | NS | 235 (NS) | NS | 63.6 | Gender, age, height, education, alcohol consumption, depression, physical activity, waist circumference, HbA1c, LDL and HDL cholesterol, diastolic blood pressure, antidepressants, antidiabetics, lipid- lowering drugs, antihypertensive medication |
| Launer 2015 | Current smokers (undefined) | Never-smoker | TBV | USA | General | 412 (NS) | 50.3 (3.50) whole sample | 107 (NS) | 50.3 (3.50) whole sample | 45.5 | Age, sex, race, ICV |
| Li 2015 | DSM nicotine dependence | <5 cigs/life, none past 10 years | Accumbens, caudate, putamen | China | General | 22 (22) | 19.3 (2.6) | 27 (27) | 19.7 (2.0) | 72.7 | Age, ICV |
| Liang 2022 | Current past 6 months, >10 CPD >2 years | lifetime < 2 pack years, not smoked for past 2 years) | Caudate, hippocampus, putamen, thalamus | USA | Physical health | SN: 43 (41), PWH: 106 (89) | SN: 47.2 (13.6), PWH: 51.0 (9.5) | SN: 65 (51), PWH: 40 (37) | SN: 43.7 (12.4), PWH: 48 (10.1) | 63.6 | Age, sex, ICV, cannabis use duration |
| Lie 2022 | >85 nmol/L Serum cotinine level, self-reported smoked regularly past 10 years | serum cotinine levels ≤85 nmol/L, did not report regular smoking, only snuff use | Thalamus, TGMV, TWMV | Norway | Physical health | 37 (15) | 37.5 (6.8) | 47 (15) | 37.9 (9.7) | 63.6 | Age, sex, vascular disease, ICV, BL EDSS, time from diagnosis |
| Lin 2021 | >10 CPD, last year | <5 cig in a lifetime | Accumbens, amygdala, caudate, hippocampus, ICV, pallidum, putamen, thalamus | USA | General | 67 (39) | M: 22.54 (3.09), F: 24.44 (3.29) | 60 (35) | M: 22.54 (2.62), F: 22.50 (2.89) | 54.5 |  |
| Linli 2023 | Currently smoke on most or all days | Never smoker | ICV, TGMV, 166 cortical and subcortical regions | UK Biobank | General | 14667 (6418) | 63.12 (7.52) | 1254 (644) | 61.84 (7.29) | 63.6 | Age, sex, handedness, ethnicity, BMI, alcohol status, imaging site, and ICV |
| Luhar 2013 | Current smoker (undefined) | Non-smoker (undefined) | Pallidum | USA | Psychiatric population | NALC: 7 (4); ALC: 7 (4) | NALC: 50.4 (9.8), ALC: 52.7 (11.2) | NALC: 6 (66.7%), ALC: 7 (57.1%) | NALC: 47.0 (7.8), ALC: 51.1 (11.2) | 54.5 |  |
| Rinigin 2022 | Current smoker (currently smoke cigarettes daily) | Non-smoker (responded no to have you ever smoked cigarettes etc., regularly) | Thalamus, hippocampus, amygdala, caudal anterior cingulate, rostral anterior cingulate, posterior cingulate, pars opercularis, pars orbitalis, pars traingularis, rostral middle frontal, lateral orbitofrontal, medial orbitofrontal, superior temporal, insula | Australia | Psychiatric population | 125 (63) | C: 37.2 (14.7), S: 38.1 (9.4) | 158 (119) | C: 40.9 (12.8), S: 37.2 (10.0) | 54.5 | Age, gender, site |
| Shang 2024 | Current smoker | Never smoker | TBV, TGMV, TWMV | UK Biobank | General | 20997 (NS) | NS | 2183 (NS) | NS | 54.5 | Age, gender |
| Shen 2017 | >10 CPD in last year, DSM nicotine dependence | <20 cig in lifetime, none in past 10 years | Amygdala | China | General | 41 (41) | 38.46 (8.60) | 84 (84) | 38.23 (6.85) | 27.3 |  |
| Wang 2020 | >10 CPD in last year, DSM nicotine dependence | <20 cig in lifetime, none in past 10 years | ICV, thalamus | China | General | 41 (41) | 38.5 (8.6) | 84 (84) | 38.2 (6.8) | 81.8 |  |
| Yu 2018 | DSM-V nicotine dependence, >10 cigs/day, CO >6 ppm | <5 cigs/life, CO <4 ppm | ICV, thalamus | China | General | 36 (36) | 20.1 (1.2) | 36 (36) | 20.8 (1.8) | 72.7 | ICV |
| Yuan 2016 | Tobacco | Non-smoker | Accumbens, caudate, putamen | China | General | 60 (52) | 19.95 (1.8) | 60 (53) | 20.0 (1.7) | 45.5 | ICV |

ALC = Alcohol drinking/dependent sample, BDI = Beck's depression inventory, BL EDSS = Baseline Expanded Disability Status Scale, C = controls, Cig = cigarette, CIS = Clinically Isolated Syndrome, CPD = Cigarettes per day, DSM = Diagnostic Statistical Manual, ENGIMA = Enhancing Neuro Imaging Genetics Through Meta Analysis, F = Female, GM = Grey matter, ICV = Intracranial Volume, M = Male, NALC = Non-alcoholic participants, NS = Not stated, NsALC = Non-smoking alcohol dependent individuals, NsLD = Non-smoking light drinkers, PWH = People with human immunodeficiency virus (HIV), S = Schizophrenia spectrum disorders, SALC = Smoking alcohol dependent individuals, SLD = Smoking light drinkers, SN = Seronegative for human immunodeficiency virus (HIV), TBV = Total brain volume, TGMV = Total grey matter volume, TWMV = Total white matter volume

**Table 4:**
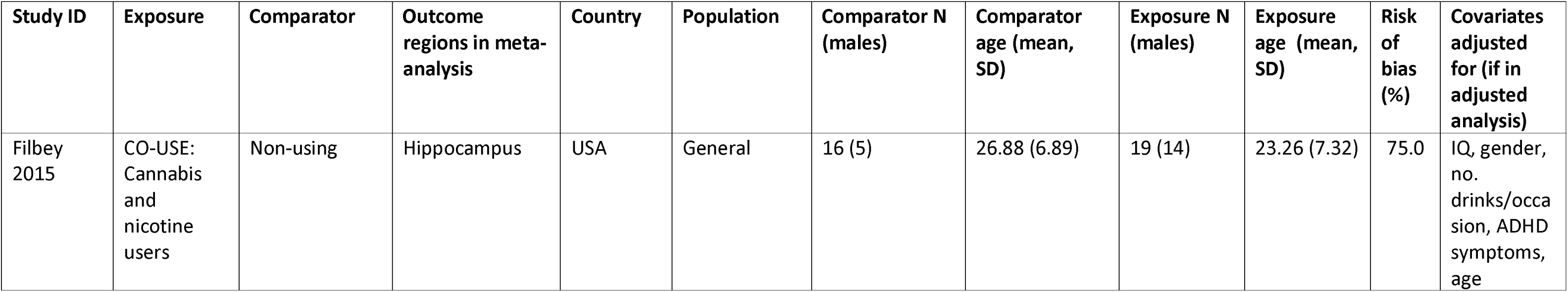
Characteristics of cross-sectional co-use study.

| Study ID | Exposure | Comparator | Outcome regions in meta-analysis | Country | Population | Comparator N (males) | Comparator age (mean, SD) | Exposure N (males) | Exposure age (mean, SD) | Risk of bias (%) | Covariates adjusted for (if in adjusted analysis) |
| --- | --- | --- | --- | --- | --- | --- | --- | --- | --- | --- | --- |
| Filbey 2015 | CO-USE: Cannabis and nicotine users | Non-using | Hippocampus | USA | General | 16 (5) | 26.88 (6.89) | 19 (14) | 23.26 (7.32) | 75.0 | IQ, gender, no. drinks/occasion, ADHD symptoms, age |

### Data extraction

We extracted the following data from studies: exposure/comparator definitions, outcome regions, study design, population group (e.g., general, psychiatric), demographics (e.g., age, sex), frequency of use, and MRI parameters. For meta-analysis the following data was extracted: effect size, standard error (or standard deviation/95% CI), p values, n per group. Studies which reported on multiple different brain measures were treated as one study with multiple outcomes. Data on all brain regions examined in each study were extracted.

Study characteristics were extracted by one reviewer (either KS, MS, CB, S Dance). Risk of bias and outcome extraction were completed independently by two reviewers for all studies (first by either KS, MS, CB, S Dance, second by either AB, MC, CC, S Daryan, SH or VL). KS checked for consistency and discrepancies were resolved by discussion or contacting a third reviewer. Where there was limited information to determine study inclusion, data extraction or synthesis, the study’s authors were contacted for further information. For more information on data extraction, see supplementary file 1.

### Risk of Bias assessment

Risk of bias (ROB) was assessed using an adapted version of the Newcastle Ottawa Scale (NOS), split by cross-sectional and longitudinal study designs (29). The NOS assesses studies on ascertainment of exposure, comparability and outcome assessment. The adapted version used in this review is outlined in supplementary file 1, tables 5 and 6. Studies were given one or two stars if presenting low risk of bias in these categories and the number of stars was then summed to give an overall score. Due to cannabis studies having one more ROB criteria, scores were standardised as percentages to aid interpretation, where higher scores indicate lower risk of bias.

**Table 5:** Characteristics of longitudinal cannabis studies.

| Study ID | Exposure | Comparator | Outcome regions in meta-analysis | Country | Population | Comparator N (males) | Comparator age (mean, SD) | Exposure N (males) | Exposure age (mean, SD) | Risk of bias (%) | Covariates adjusted for (if in adjusted analysis) |
| --- | --- | --- | --- | --- | --- | --- | --- | --- | --- | --- | --- |
| Garimella 2020 | >10 days/month at least two years, no treatment for CUD | <50x life, not used in past year | Hippocampal tail, Subiculum, CA1, Presubiculum, Parasubiculum, CA3, CA4, Fimbria, Whole hippocampus, Hippocampal fissure, molecular layer, GC-ML-DG (molecular layer, dentate gyrus), HATA (Hippocampus-amygdala transition area) | Netherlands | General | 17 (11) | 21.5 (2.67) | 17 (14) | 20.7 (2.2) | 68.8 |  |
| Koenders 2017 | >10 uses/month at least 2 years, smoked joints | <50 lifetime occasions | Hippocampus | The Netherlands | General | 23 (13) | 21.79 (2.6) | 20 (14) | 20.64 (2.23) | 68.8 |  |
| Luo 2022 | Use at baseline | Non-use at baseline | Superior temporal sulcus, caudal anterior cingulate cortex, caudal middle frontal cortex, cuneus cortex, entorhinal cortex, fusiform gyrus, inferior parietal cortex, inferior temporal gyrus, isthmus-cingulate cortex, lateral occipital cortex, lateral orbital frontal cortex, lingual gyrus, medial orbital frontal cortex, middle temporal gyrus, parahippocampal gyrus, paracentral lobule, IFG pars opercularis,<br>IFG pars<br>orbitalis, IFG<br>pars<br>triangularis,<br>pericalcarine<br>cortex,<br>postcentral<br>gyrus,<br>posterior-<br>cingualte<br>cortex,<br>precentral<br>gyrus,<br>precuneus<br>cortex, rostral<br>anterior<br>cingualte<br>cortex, rostral<br>middle frontal<br>gyrus, superior<br>frontal gyrus,<br>superior<br>parietal cortex,<br>superior<br>temporal<br>gyrus,<br>supramarginal<br>gyrus, frontal<br>pole, temporal<br>pole,<br>transverse<br>temporal cortex, insula | USA, NCANDA study | General | 510 (NS) | 16.02 (2.45) whole sample | 17 (NS) | 16.02 (2.45) whole sample | 68.8 |  |
| Rais 2008 | self-reported cannabis use at baseline | no lifetime use | ICV, TBV, cerebral grey matter, cerebral white matter | Netherlands | Psychiatric population | C: 31 (25), FES: 32 (26) | C: 24.72 (6.66), FES: 23.28 (5.10) | FES: 19 (19) | FES: 21.83 (3.91) | 43.8 |  |
| Wang 2021 | >10 days/month at least two years, no treatment for CUD | <30 lifetime uses | Hippocampus, CA1, CA2/CA3, CA4/DG, SR/SL/SM, Subiculum | The Netherlands | General | 22 (14) | 21.56 (2.45) | 20 (15) | 20.53 (2.11) | 56.3 |  |
| Welch 2011 | use in 2 year follow up period | No use in 2 year follow up | Thalamus | Edinburgh High Risk Study | Psychiatric population | 32 (15) | 21.11 (2.87) | 25 (15) | 21.76 (2.52) | 62.5 |  |
| Xu 2022 | >10 days/month at least two years, no treatment for CUD | Non (less than 30x life, not used in past year) | TGMV, TWMV, Accumbens, Amygdala, Caudate, Pallidum, Putamen, Thalamus | The Netherlands | General | 22 (14) | 21.56 (2.45) | 20 (15) | 20.53 (2.11) | 56.3 |  |

CA1 = cornu Ammonis1, CA3 = cornu Ammonis 3, CA4 = cornu Ammonis 4, CUD = Cannabis use disorder, FES = First episode schizophrenia, GC-ML-DG = Hippocampus molecular layer dentate gyrus, HATA = Hippocampus-amygdala transition area, ICV = Intracranial Volume, IFG = Inferior frontal gyrus, NS = Not stated, SR/SL/SM = Stratum Radiatum/Stratum Lacunosum/Stratum Moleculare, TBV = Total brain volume, TGMV = Total grey matter volume, TWMV = Total white matter volume

**Table 6:** Characteristics of longitudinal tobacco studies.

| Study ID | Exposure | Comparator | Outcome regions in meta-analysis | Country | Population | Comparator N (males) | Comparator age (mean, SD) | Exposure N (males) | Exposure age (mean, SD) | Risk of bias (%) | Covariates adjusted for (if in adjusted analysis) |
| --- | --- | --- | --- | --- | --- | --- | --- | --- | --- | --- | --- |
| Duriez 2014 - longitudinal | current smoker | Never smoker | TBV, TGMV | France | General | 714 (118) | M: 72.0 (3.93), F: 72.3 (3.93) whole sample | 63 (37) | M: 72.0 (3.93), F: 72.3 (3.93) whole sample | 53.3 | ICV, Age, educational level, MMSE score, BMI, CES-D score, SBP, DBP, fasting blood glucose, cholesterol emia, alcohol consumption, WML, and Apo-ε4 charge |
| Kim 2018 | Heavy smoker >23 pack years | Nonsmokers | TBV | Korea | General | 603 (101) | M: 59.8 (2.27), F: 59.2 (7.0) whole sample | 103 (95) | M: 59.8 (2.27), F: 59.2 (7.0) whole sample | 53.3 | ICV, age, sex, baseline volume, waist-to-hip ratio, hypertension n, and diabetes mellitus |
| Otsuka 2022 | Self-reported current smokers | Self-reported never smoker | TGMV | Japan | General | 1451 (674) | M: 60.24 (11.72), F: 59.21 (11.87) whole sample | 214 (175) | M: 60.24 (11.72), F: 59.21 (11.87) whole sample | 60.0 | Age, TGMV at baseline, education, family income, living alone, depressive symptoms and history of stroke, dyslipidemia, diabetes, hypertension, or heart disease. |
| Van Haren 2010 | Self-reported current smokers | Self-reported nonsmoker | Cerebellum | The Netherlands | Psychiatric | C: 78 (NS), S: 42 (NS) | NS | C: 35 (NS), S: 54 (NS) | NS | 53.3 |  |

C = Controls, CES-D = Centre for Epidemiologic Studies-Depression scale, DBP = diastolic blood pressure, F = Female, ICV = Intracranial Volume, M = Male, MMSE = mini-mental scale examination, NS = Not stated, S = Schizophrenia patients, SBP = Systolic blood pressure, TBV = Total brain volume, TGMV = Total grey matter volume, WML = White Matter lesions

Quality of MR studies was assessed using a quality assessment framework undertaken in a previous review of MR studies in addiction (30). The system gives each study a total score of either ‘–’, ‘– +’or ‘+’, based on key factors important in the validity of MR studies (phenotype measurement, instrument strength, sample size and analytical methods (30). For a study to be scored as sufficient (‘-+’) the study must have sufficient sample size and main analytical methods (e.g. instrument strength, temporality and harmonisation, same ethnic group, sample overlap reported, analyses addressing horizontal pleiotropy;(30)).

### Meta-analysis of brain volume

Specific brain regions were included in the meta-analysis if they were examined by at least three studies. Analysis was organised by study exposure and methodology (cross sectional, longitudinal) for each outcome region. For each region we calculated Hedges’ g for differences between user and non-user groups in individual studies, using the esc package in R (31,32). Hedges’ g represents the effect size of the standardised mean difference (33) and was used to allow for variation in outcome measures and small sample bias (common to neuroimaging studies) (33). Where necessary, Hedges’ g values were aggregated across left and right hemispheres, see supplementary file 1 for more information. Individual Hedges’ g estimates were then synthesised using generic inverse variance random effects meta-analysis for each exposure comparison in distinct brain regions, using the meta and metafor packages in R (31,34,35). Random effects models were used to account for heterogeneity across studies. Meta-analyses were split into adjusted and unadjusted estimates for each outcome, for more information see supplementary file 1.

Statistical heterogeneity was quantified using I. As I values between 50-75% indicate substantial heterogeneity, values in this range or above were considered problematic and associated meta-analysis results should be treated with caution (33). Small study bias was assessed using funnel plots, for evidence of asymmetry, and Egger’s test. If there was evidence for bias, then Egger’s trim and fill was conducted for that outcome (36).

All analyses were conducted using R version 4.4.2. The R script and data are available on Open Science Framework (DOI 10.17605/OSF.IO/SFPK2). Studies were summarised narratively where a study met inclusion criteria but there was not enough information for inclusion in any meta-analysis.

## RESULTS

The search identified 24,809 records. Further cross-referencing from searching relevant systematic reviews in the field (15,18) identified an additional two records, totalling 24,811. Removal of duplicates resulted in 18,294 studies for title/abstract screening. Of these studies, 17,920 were excluded due to not meeting eligibility criteria. The full text of 374 studies were screened. Of those studies 271 were excluded (see supplementary file 1, table 32, for list of studies excluded with reasons), leaving 103 independent studies included in the review, and 77 included in the meta-analysis (see table 1 for full list of included studies). See supplementary file 1, figure 1 for the PRISMA flowchart.

**Figure 1.**
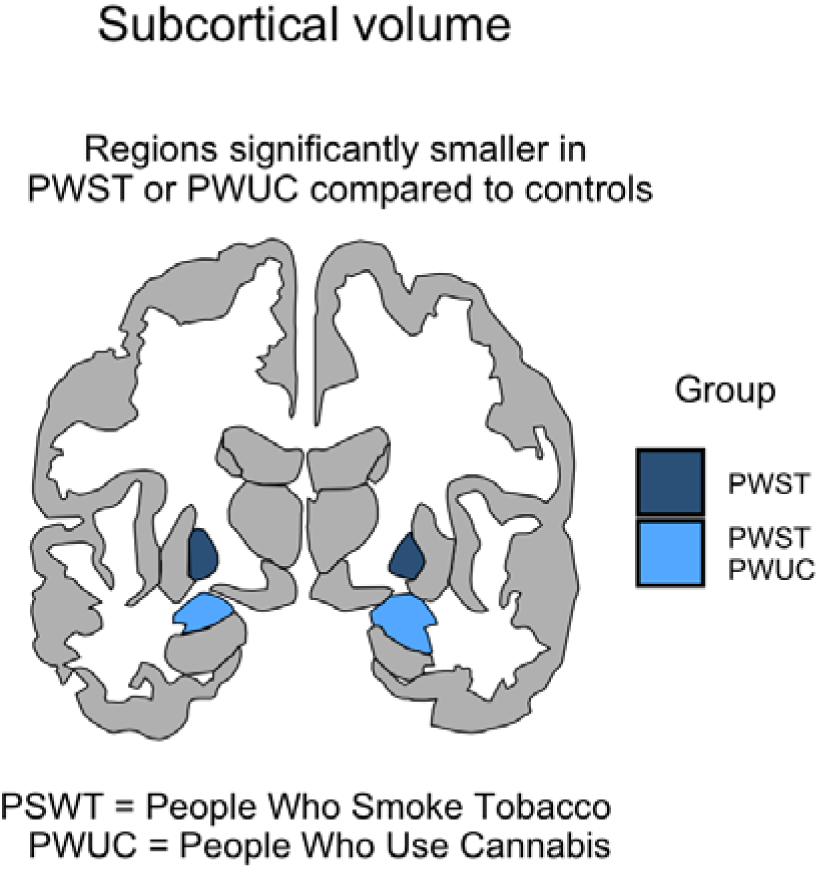
Subcortical differences from adjusted analysis between people who use cannabis, tobacco and people who do not.

**Figure 2.**
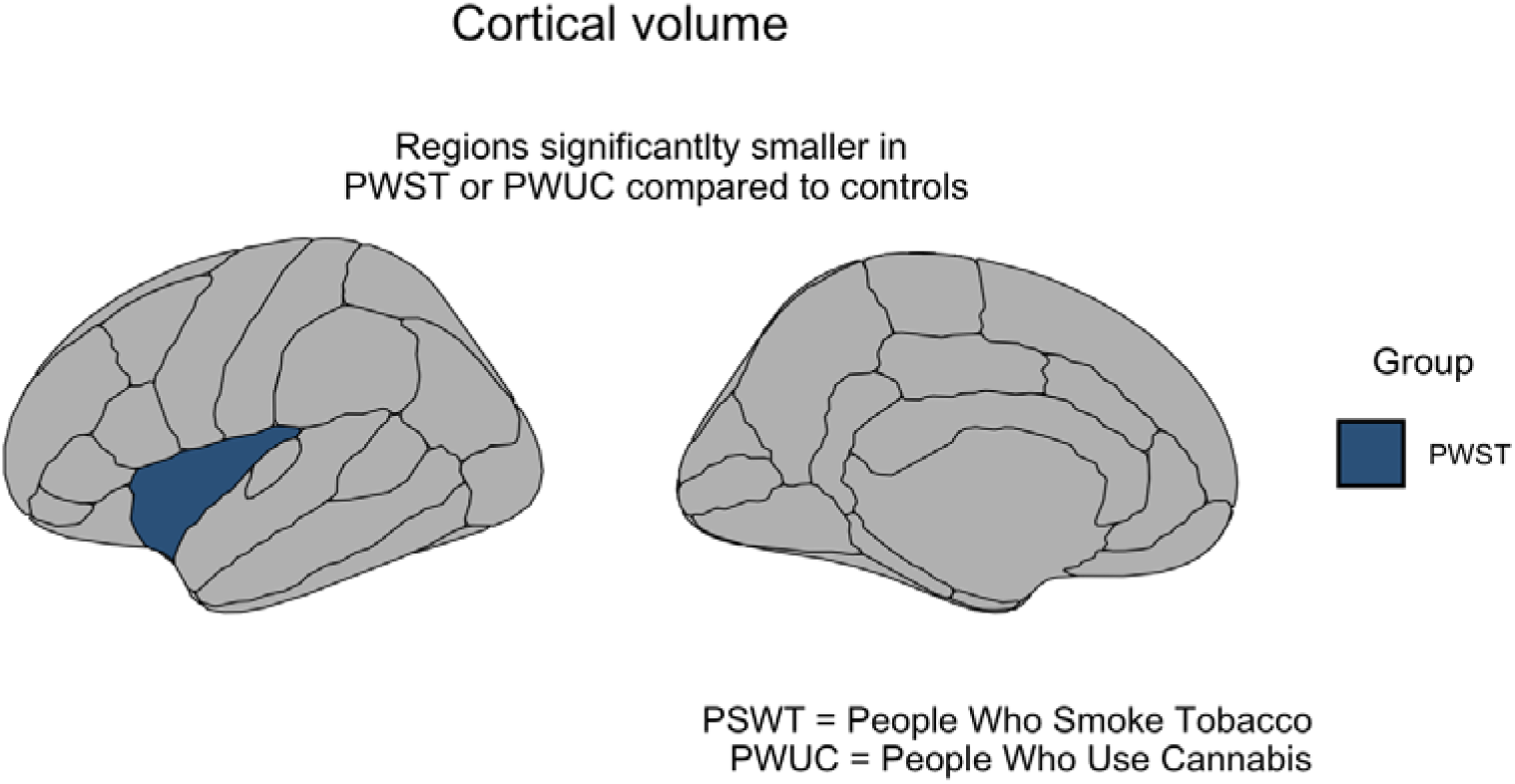
Cortical differences between people who use cannabis, tobacco and people who do not.

**Figure 3.**
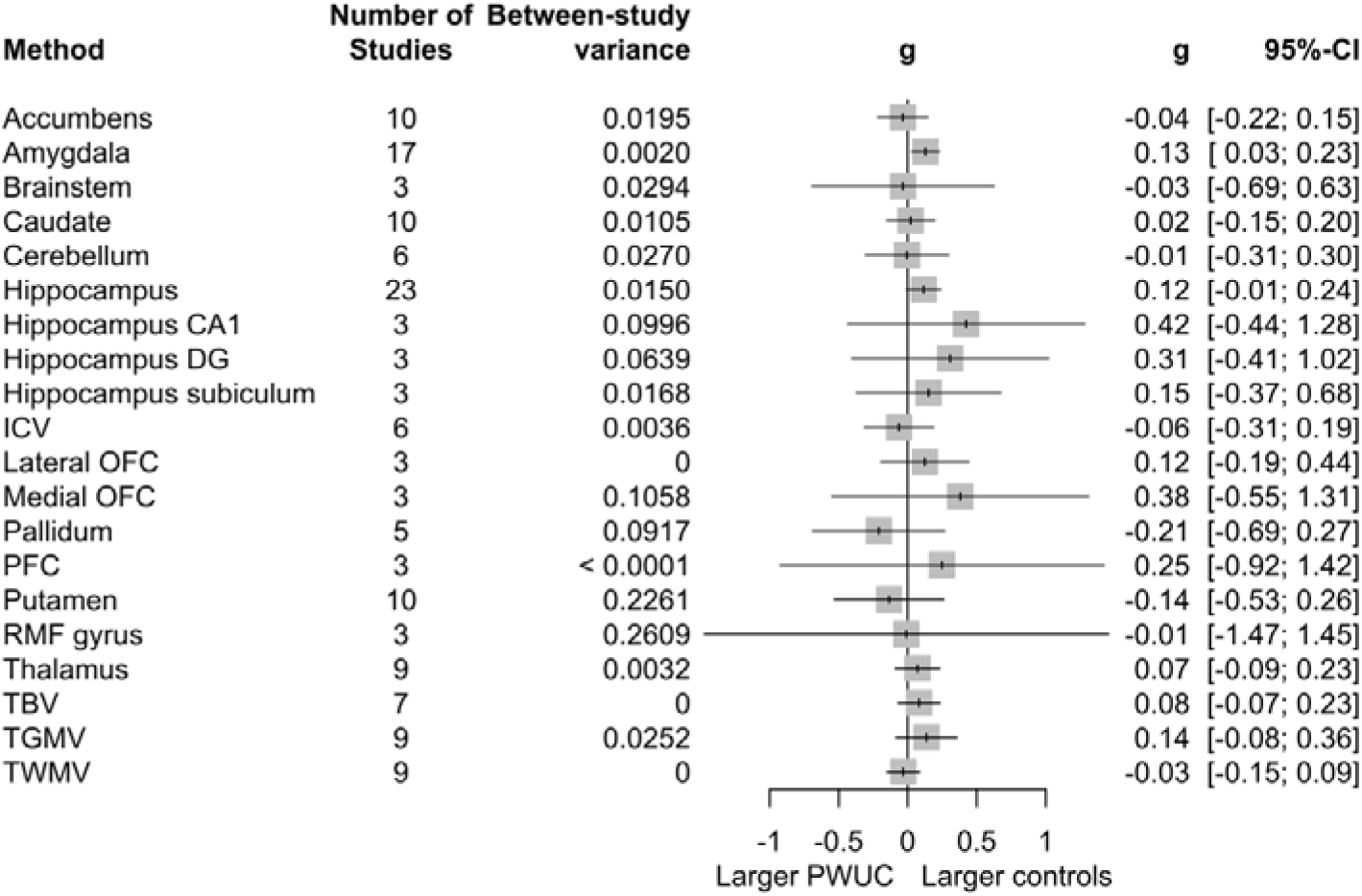
Summary forest plot of adjusted cross-sectional meta-analyses of differences in brain volume between People Who Use Cannabis (PWUC) and controls. Results presented to the nearest four decimal places; 0 refers to <0.0001.

### Overview of sample characteristics

Study characteristics are reported in tables 2-7. For studies included in the cross-sectional meta-analysis of cannabis use, the overall sample comprised 18,247 (58% male) participants, aged between 14 – 70 years, of which 1,965 (62% male) were people who use cannabis and 16,282 (57% male) were controls. In the cross-sectional tobacco analysis, the overall sample comprised 51,194 (24% male) participants, aged between 19 – 73 years, of which 6,453 (36% male) were people who smoked tobacco and 44,741 (22% male) were controls. For the longitudinal tobacco analysis, the overall sample comprised 3,357 (36% male) participants, aged between 59 – 72 years, of which 469 (66% male) were people who smoked tobacco and 2,888 (30% male) were controls. Average time of follow up was 3.78 years (range 2 – 5). MRI data acquisition and processing varied between studies (supplementary file 1, tables 7-11).

**Table 7:**
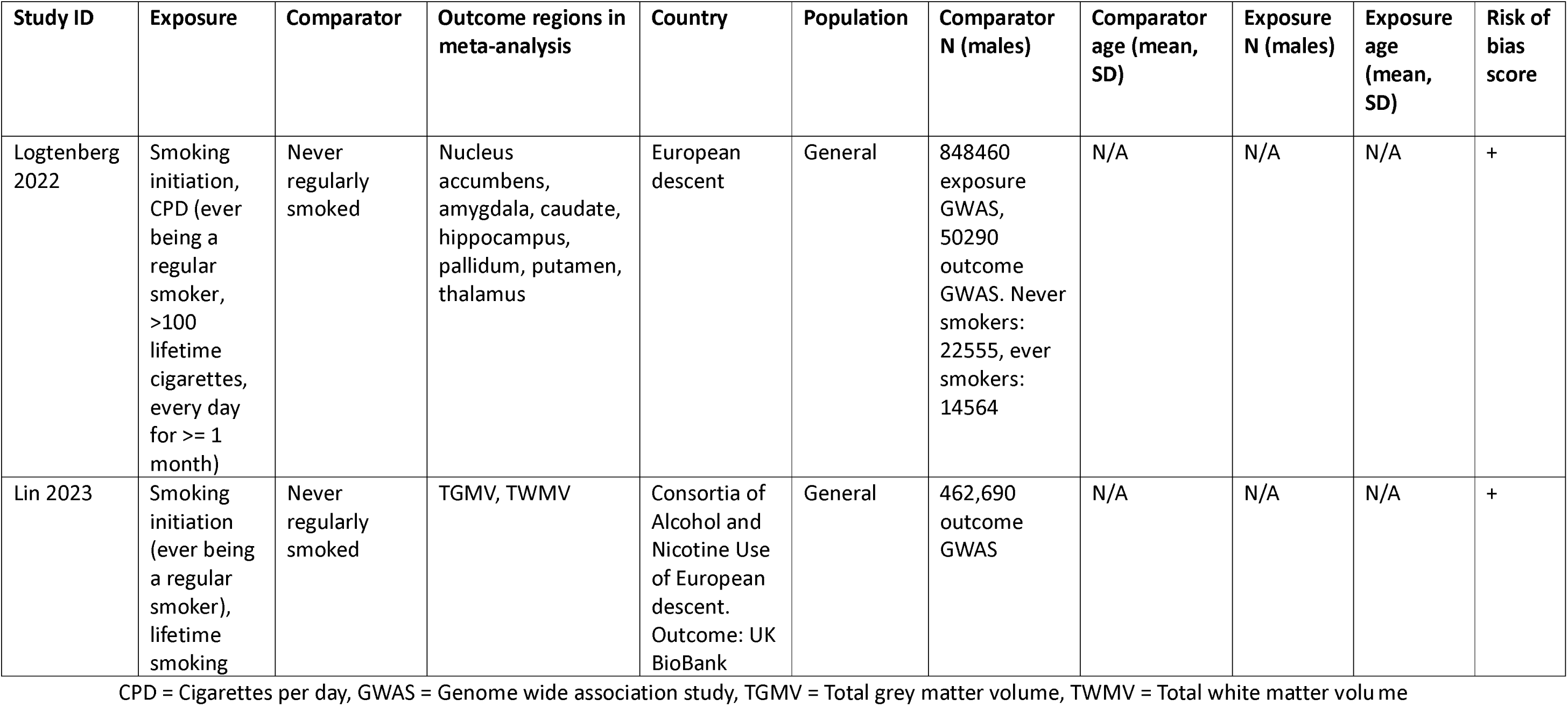
Characteristics of MR tobacco studies.

| Study ID | Exposure | Comparator | Outcome regions in meta-analysis | Country | Population | Comparator N (males) | Comparator age (mean, SD) | Exposure N (males) | Exposure age (mean, SD) | Risk of bias score |
| --- | --- | --- | --- | --- | --- | --- | --- | --- | --- | --- |
| Logtenberg 2022 | Smoking initiation, CPD (ever being a regular smoker, >100 lifetime cigarettes, every day for >= 1 month) | Never regularly smoked | Nucleus accumbens, amygdala, caudate, hippocampus, pallidum, putamen, thalamus | European descent | General | 848460 exposure GWAS, 50290 outcome GWAS. Never smokers: 22555, ever smokers: 14564 | N/A | N/A | N/A | + |
| Lin 2023 | Smoking initiation (ever being a regular smoker), lifetime smoking | Never regularly smoked | TGMV, TWMV | Consortia of Alcohol and Nicotine Use of European descent. Outcome: UK BioBank | General | 462,690 outcome GWAS | N/A | N/A | N/A | + |

CPD = Cigarettes per day, GWAS = Genome wide association study, TGMV = Total grey matter volume, TWMV = Total white matter volume

### Risk of bias assessments

Detailed risk of bias assessment results can be found in supplementary tables file 1, tables 12-14 for cross-sectional studies, and 15 and 16 for longitudinal studies. Summary scores are reported in tables 2-7. For cross-sectional studies, the average NOS risk of bias score was 53.8% (25.0%-83.3%) for cannabis and 54.3% (18.2%-81.8%) for tobacco, suggesting a moderate-substantial risk of bias. For longitudinal studies, the average NOS risk of bias score was 60.7% (43.8%-68.8%) for cannabis and 60.7% (43.8%-68.8%) for tobacco, again suggesting a moderate-substantial risk of bias.

For the two MR studies, the quality rating was ‘+’ for both (37,38). It is important to note that both studies used summary level data from the same datasets to obtain their genetic estimates, for both the exposures and outcomes. Therefore, studies scored similar on phenotype measurement, and instrument strength. Both studies used extensive additional sensitivity methods, resulting in scores of ‘+’ (see supplementary file 1, table 17).

### Cross-sectional studies: meta-analysis results

#### Cannabis vs control

Fifty cross-sectional studies were identified, of which 44 were included in the meta-analysis. A summary of meta-analysis results is displayed in figures 1-4 and supplementary file 1 tables 18-21. Forest plots of analysis for each individual region are in supplementary file 2. Narrative results are in supplementary file 1, table 28.

**Figure 4.**
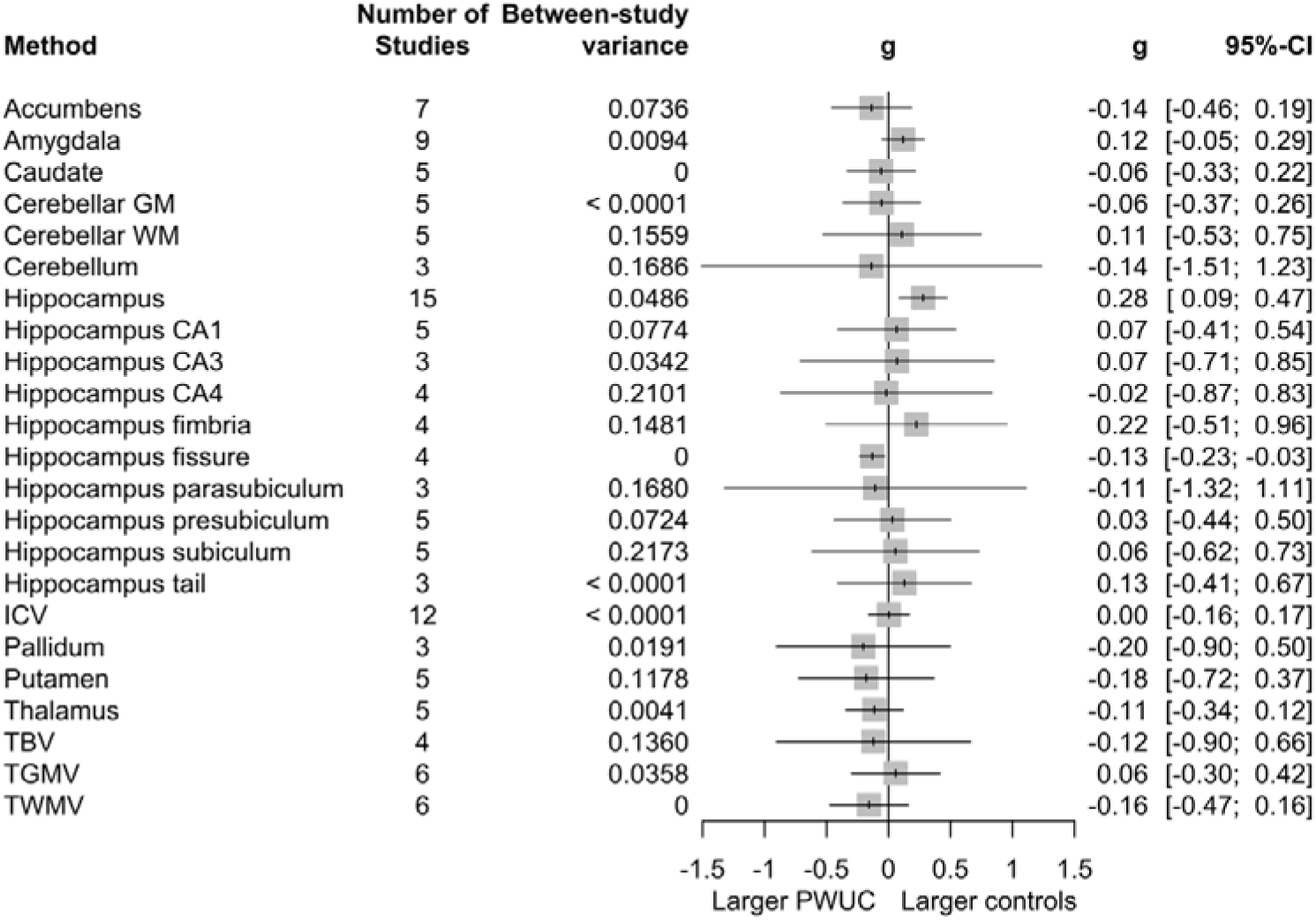
Summary forest plot of unadjusted cross-sectional meta-analyses of differences in brain volume between People Who Use Cannabis (PWUC) and controls. Results presented to the nearest four decimal places; 0 refers to <0.0001.

**Figure 5.**
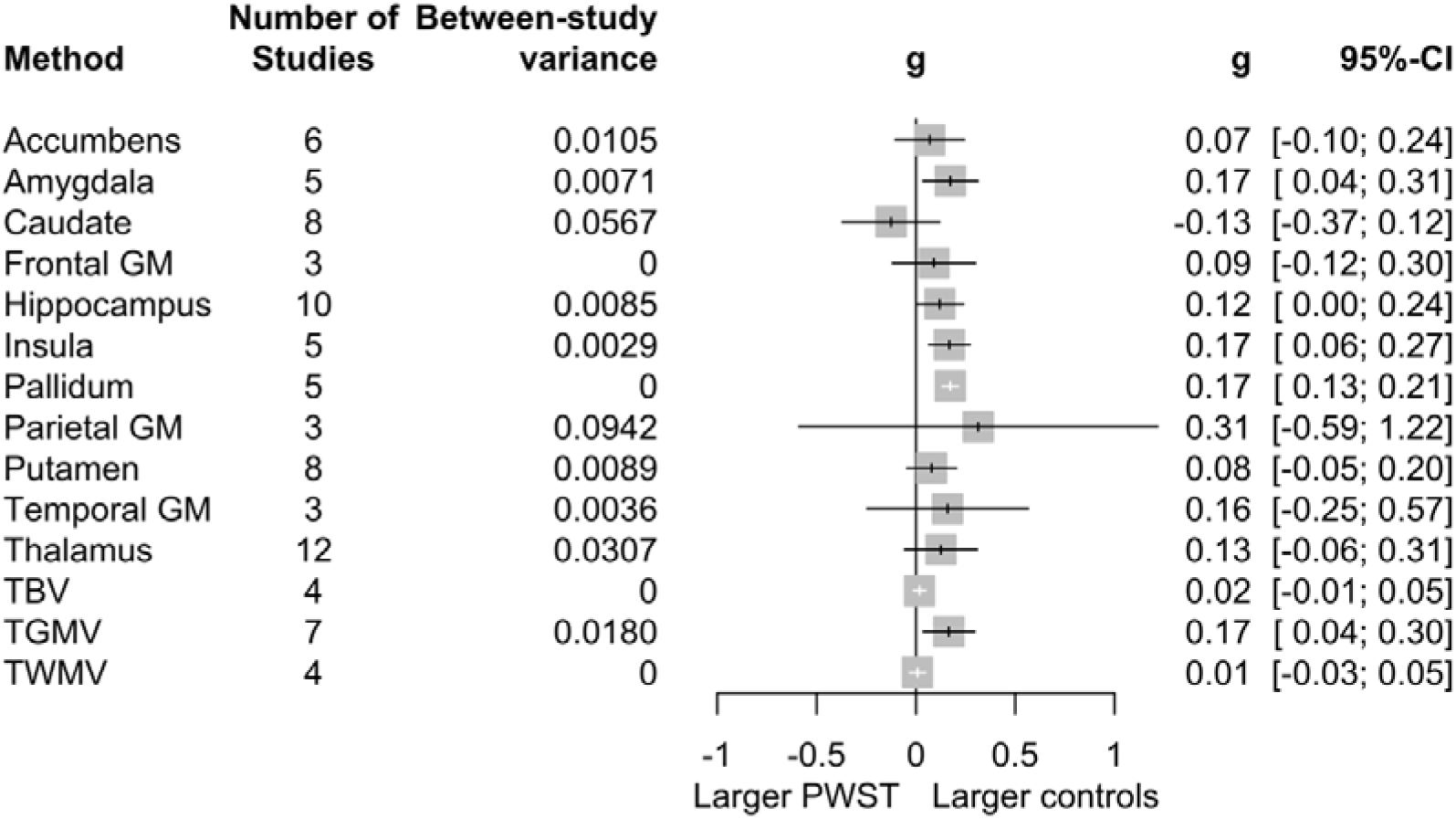
Summary forest plot of adjusted cross-sectional meta-analyses of differences in brain volume between People Who Smoke Tobacco (PWST) and controls. Results presented to the nearest four decimal places; 0 refers to <0.0001.

In the adjusted analysis, there was a difference in amygdala volume between people who use cannabis and controls, with people who use cannabis having smaller amygdala volume compared to controls with a small effect size (k = 17, Hedges’ g = 0.13, 95% CI [0.03, 0.23], p = 0.016, I = 22.90%). There was no evidence of differences for any other region investigated in the adjusted analysis. Egger’s test suggested asymmetry in the literature for ICV but implementing Egger’s trim and fill did not change these results. I heterogeneity estimates varied across regions, but was generally moderate to substantial, with highest heterogeneity for the hippocampus CA1, hippocampus dentate gyrus (DG), medial orbitofrontal cortex (OFC), pallidum, putamen and rostral middle frontal gyrus.

In the unadjusted analysis there were differences in the volumes of the hippocampus and hippocampus fissure between people who use cannabis and controls. People who use cannabis had a smaller hippocampus than controls (k = 15, g = 0.28, 95% CI [0.09, 0.47], p = 0.008, I = 53.50%), but a larger fissure than controls (k = 4, g = −0.13, 95% CI [−0.23, −0.03], p = 0.026, I = 0%). However, Egger’s test suggested asymmetry in the unadjusted hippocampus analysis and once Egger’s trim and fill was implemented, differences in hippocampus volume between users and controls were no longer present (k = 19, g = 0.13, 95% CI [−0.12, 0.38], p = 0.279, I = 68.70%). I heterogeneity estimates varied across regions, but were generally moderate to substantial, with highest heterogeneity estimates for the hippocampus, hippocampus CA4, fimbria, parasubiculum and subiculum, putamen, accumbens, and cerebellum.

#### Tobacco vs control

Forty cross-sectional studies were identified for inclusion, 30 of which were included in the meta-analysis. A summary of meta-analysis results is in figure 1,2,5 and 6. and supplementary file 1 tables 22-25. Forest plots of analysis for each individual region are in supplementary file 2. Narrative results in supplementary file 1, table 29.

In the adjusted analysis, there were volumetric differences between people who smoke tobacco and controls in the amygdala, insula, pallidum, TGMV, and the hippocampus. There were no differences for any other region investigated in the adjusted analysis. People who smoke tobacco had smaller volume in the amygdala (k = 5, g = 0.17, 95% CI [0.04, 0.31], p = 0.025, I = 55.90%), insula (k = 5, g = 0.17, 95% CI [0.06, 0.27], p = 0.011, I = 31.20%), pallidum (k = 5, g = 0.17, 95% CI [0.13, 0.21], p = <0.0001, I = 0%) and TGMV (k = 7, g = 0.17, 95% CI [0.04, 0.30], p = 0.020, I = 93.70). There was weak evidence for smaller hippocampus volume in people who smoke tobacco, compared to controls (k = 10, g = 0.12, 95% CI [0.00, 0.24], p = 0.049, I = 62.30%). Egger’s test suggested asymmetry in the caudate, insula, parietal grey matter and putamen analyses, but implementing Egger’s trim and fill did not change the conclusions for these regions. Across regions, I heterogeneity estimates varied, and were generally either low (with scores of <0.0001) or moderate to substantial, with highest heterogeneity estimates for TGMV.

In the unadjusted analysis, there were no differences in volume between people who smoke tobacco and controls for any brain region (figure 6). There was no evidence of asymmetry for any regions. I heterogeneity estimates varied across regions, but were mostly either substantial or low-moderate, with highest heterogeneity estimates for the thalamus and caudate.

**Figure 6.**
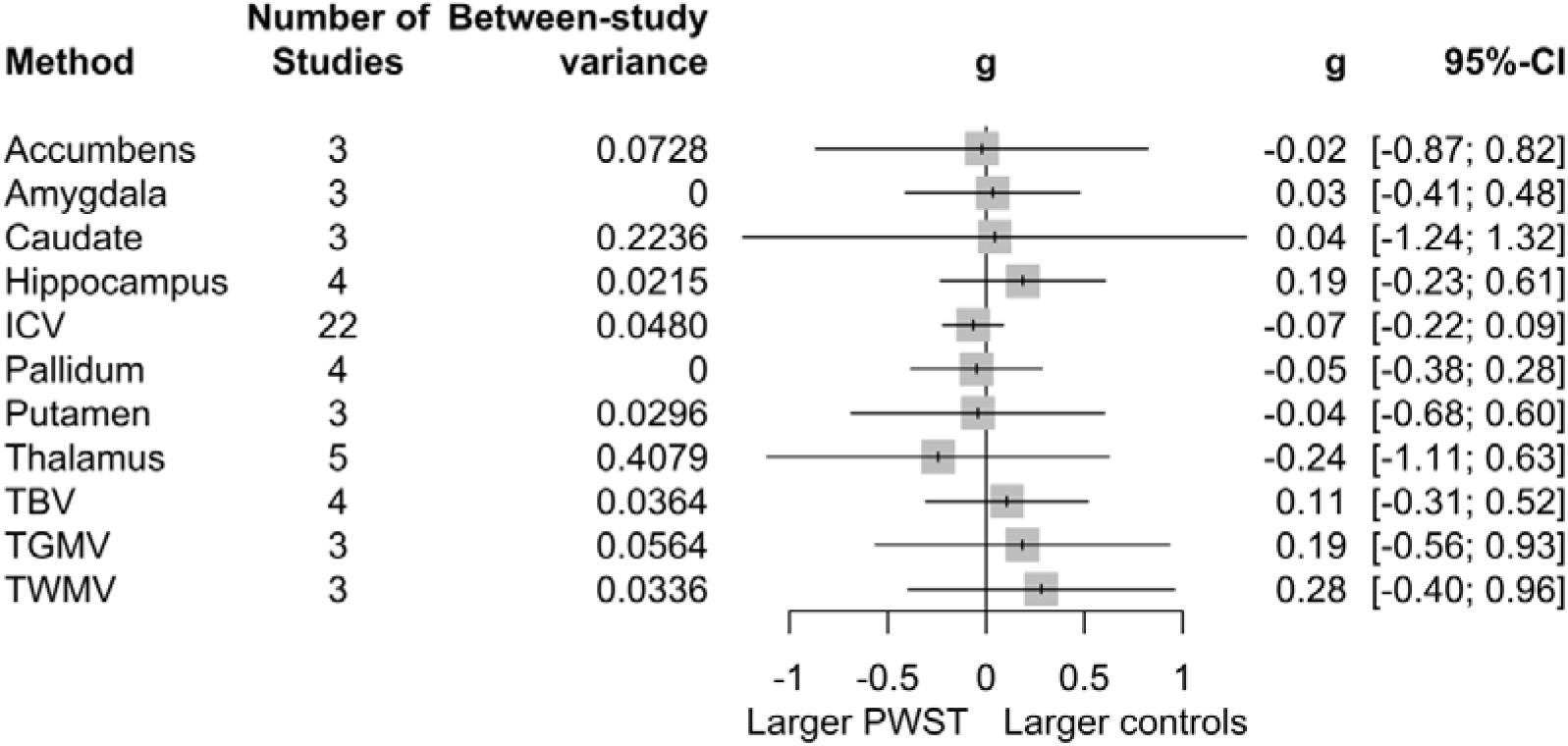
Summary forest plot of unadjusted cross-sectional meta-analyses of differences in brain volume between People Who Smoke Tobacco (PWST) and controls. Results presented to the nearest four decimal places; 0 refers to <0.0001.

#### Co-use of cannabis and tobacco vs control

There was one eligible cross-sectional study which investigated co-use (25). Filbey et al., (2015) compared bilateral hippocampal volumes in groups with combined use of cannabis and nicotine with cannabis only, nicotine only and non-using groups. The authors found a main effect of group for the volume of the right hippocampus (F (3,77) = 4.36, *p* = 0.007), but not the left hippocampus (F (3,77) = 1.576, *p* = 0.202), whilst controlling for alcoholic drinks, IQ, gender and age. Post-hoc pairwise comparisons suggest people who used only cannabis had smaller right hippocampus volumes compared to the controls (F (1, 43) = 9.23, p = 0.004) and nicotine only groups (F (1,45) = 5.79, p = 0.02). But for the group who co-used cannabis and nicotine there were no differences compared to controls (F (1, 27) = 2.96, p = 0.09) or nicotine only groups (F (1,39) = 2.75, p = 0.11).

### Longitudinal studies

#### Longitudinal changes in cannabis vs control groups

Seven longitudinal studies using independent samples were identified for inclusion, none of which were meta-analysed because there were no brain regions with more than two independent samples. A narrative summary can be found in supplementary file 1, table 30. In brief, most studies found no longitudinal volumetric differences in between people who use cannabis and those who do not, including in cortical regions (39), global measures of brain integrity and subcortical regions (40). No differences were identified in volumes of the hippocampus (41–43) and hippocampus subfields (41,43). Garimella et al., (41) found volumetric increases in the right parasubiculum, right fimbria, and the right and left CA3 in people who use cannabis compared to controls. Contrastingly, Wang et al., (43) identified a faster rate of volume decrease for the right hippocampus in people who use cannabis compared to controls. In patients with schizophrenia, Rais et al., (44) found larger TGMV loss in patients who used cannabis compared to non-using patients and non-using controls. Welch et al., (45)reported that in people with familial high risk for schizophrenia, there was greater right thalamic volume loss from baseline to follow up, in people who use cannabis compared to controls.

#### Longitudinal changes in tobacco vs control groups

Four unique longitudinal studies were identified for inclusion, three of which were included in the meta-analysis for only two outcome regions. Total brain volume (TBV) and TGMV were the only outcome regions with at least three samples to be meta-analysed (see figure 7). For both TBV and TGMV, all studies reported a decrease in brain volume between baseline to follow up. For TBV, there was no difference in TBV between people who use tobacco and controls (k = 3, g = 0.11, 95% CI [− 0.11, 0.32], p = 0.170, I = 0%). For TGMV, there was a greater decrease in TGMV in people who use tobacco compared to controls (k = 5, g = 0.05, 95% CI [0.01, 0.10], p = 0.037, I = 0%). There was no evidence of heterogeneity or asymmetry (see supplementary file 1, tables 26 and 27, and supplementary file 2). A narrative summary can be found in supplementary file 1 table 31.

**Figure 7.**
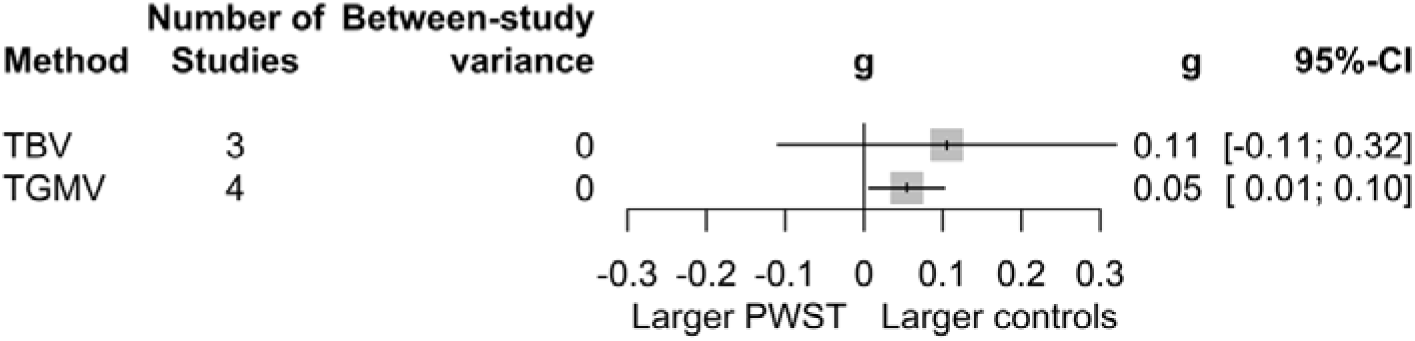
Summary forest plot of adjusted longitudinal meta-analyses of differences in brain volume between People Who Smoke Tobacco (PWST) and controls. A positive value indicates a greater reduction in PWST compared to controls. Results presented to the nearest four decimal places; 0 refers to <0.0001.

#### Longitudinal changes in co-use vs control groups

There were no longitudinal studies which investigated cannabis and tobacco co-use and brain volume.

### Mendelian randomisation studies

#### Cannabis use

There were no studies which used MR to investigate the causal effect of cannabis on brain volume.

#### Tobacco use

We identified two tobacco MR studies (37,38). Logtenberg et al., (38) investigated bidirectional associations between smoking and subcortical brain volume. They found weak evidence that smoking initiation might decrease amygdala volumes (*β* = −0.05, p = 0.046). There was strong evidence that smoking more cigarettes per day (CPD) might decrease hippocampus volumes (*β* = - 94.73, p = 1.8E-06). Lin et al., (37) investigated the links between smoking initiation and grey/white matter volume and found that smoking initiation might decrease TGMV (*β* −0.100, 95% CI [−0.156, - 0.043], p = 0.000523), but this was attenuated when adjusting for alcohol drinking using multivariable MR (*β* = −0.056, se = 0.037, p = 0.122). Lin et al., (2023) found no effect of smoking initiation on white matter volume (*β* = −0.004, 95% CI [−0.065, 0.058], p = 0.907).

#### Co-use

There were no studies which used MR to investigate the causal effect of cannabis and tobacco co-use on brain volume.

## DISCUSSION

Overall, this systematic review and meta-analyses demonstrated that cross-sectional evidence suggests people who use cannabis had smaller volumes in the amygdala. There were smaller volumes in the amygdala, insula and pallidum in people who smoke tobacco compared to non-smokers. The evidence from longitudinal studies is limited, with only seven studies investigating cannabis and four investigating tobacco. For cannabis, narrative synthesis identified limited consistency in evidence for differences in brain volume between groups. For tobacco, meta-analyses showed no differences in TBV, but did identify smaller TGMV in people who smoked tobacco compared to controls. Evidence from two MR studies suggests that smoking more cigarettes might decrease hippocampus volume and smoking initiation might reduce amygdala volume (38) and TGMV (37). Only one study examined the co-use of cannabis and tobacco, reporting that reductions in right hippocampal volumes were associated with cannabis use, but not cannabis and nicotine co-use or nicotine use alone (25).

The literature assessing cannabis and/or tobacco use and brain volume was mostly cross-sectional, which limits our ability to make causal inferences on the effects of these substances on brain structure. Longitudinal study designs should be used as well as methods which allow improved understanding of causality, such as well-designed instrumental variable studies, particularly in a triangulation framework. Furthermore, larger studies powered to detect small effects with consideration of key confounders are needed. Clearer reporting of non-significant results and sample overlap is needed to facilitate accurate evidence synthesis.

Our meta-analysis of adjusted estimates identified smaller amygdala volumes, updating earlier findings from previous meta-analyses that reported no significant differences (15,18,46). This discrepancy may be due to the inclusion of a greater number of studies in the current review, which likely increased the statistical power to detect subtle effects. Due to our conservative exclusion of samples with fully and partially overlapping participants, this review did not assess all relevant regions, as there was not enough independent studies available for meta-analysis, e.g. the lateral OFC and anterior cingulate cortex where previous reviews have found reductions in people who use cannabis (15).

In our review we found consistent differences in global brain volume between people who smoke and controls across cross-sectional, longitudinal studies and MR methods. There was consistent evidence for amygdala reductions in people who smoke tobacco across cross-sectional and MR methods, suggesting a potential causal effect (38). Although, Horizontal pleiotropy is likely present, whereby genetic variants influence brain volume through pathways other than smoking, or the causal relationship between smoking and amygdala volume may operate via multiple distinct mechanisms, such as risk-taking (38,47).

Consistent with the most recent umbrella review of voxel-based morphometry (VBM) reviews examining smoking and grey matter volume (23), we found that the insula was smaller in people who currently smoke tobacco compared to controls. We identified additional smaller volumes in the amygdala, pallidum and TGMV. Van De Weijer et al., (23) also concluded that there was consistent evidence for smaller volumes in the prefrontal cortex and cingulate cortex in people who smoke compared to controls. The prefrontal cortex and cingulate cortex were not meta-analysed in this review due to insufficient independent studies that reported extracted volumes for these regions.

The group differences identified in the reviewed literature could be attributable to the primary psychoactive components of each substance. For instance, THC could cause structural changes through reducing the number of synapses (48–50), reducing neuronal density (51), and reduction in dendritic length (50) or complexity (49). The anatomically specific effects of THC on the amygdala and hippocampus in our review could be due to the high density of CB1 receptors (52–54). Similarly, nicotine has been associated with cell loss, altered cell size and reduced dendritic length and complexity. Brain regions associated with tobacco use in this review have a high density of nicotinergic receptors (55–57).

Brain volume changes could also be attributed to harms of combustion. Combustion can cause high concentrations of reaction oxygen species (ROS), which cause oxidative stress and inflammation (increase of proinflammatory cytokines), leading to neuronal cell death (58–62). Tobacco smoke contains thousands of harmful carcinogens and combustion products, so the global effect of tobacco on total grey matter volume on the brain could be attributed to oxidative stress and inflammation from the combustion of these (59). While cannabis smoke also contains harmful byproducts, cannabis also contains CBD which has been recognised for antioxidative/anti-inflammatory effects. This could explain why limited differences were found for cannabis use, although evidence for the protective effects of CBD is inconsistent (58) and differences in exposure to CBD or other cannabinoids was not provided in individual studies.

Understanding neurobiological actions of cannabis, tobacco and co-use could inform public health interventions. For example, Vermont department of health have released posters warning young people of the harms of cannabis for brain development (63). Given that this review found evidence for differences attributed to tobacco, public health campaigns could also include messaging around potential harms of tobacco use on the brain. Interventions could also consider implications of co-use of cannabis and tobacco and whether supplementing cannabis with tobacco is a technique for harm reduction or if this increases harm. Considering the limited previous research, future studies should look at effects of co-use on the brain to better disentangle effects.

### Strengths and Limitations

To our knowledge, this is the most comprehensive synthesis of studies exploring the association between cannabis use, tobacco use and brain volume to date. The inclusion of different study designs; cross-sectional, longitudinal and Mendelian randomisation, is a strength. Another strength of this review was the separation of adjusted and unadjusted meta-analysis. Although, there were inconsistencies in the adjusted estimates, for example, some studies adjusted for ICV only, whereas others adjusted for multiple confounding variables (e.g., other drug use, alcohol use), which introduced heterogeneity into the analysis. However, by presenting both sets of analyses and considering evidence from adjusted estimates, the results of this meta-analysis may provide stronger evidence for associations with the exposures of interest and minimised confounding by other variables. Due to the vast extent of the literature reviewed herein, this review focused on binary categorisations of current use or non-use. It is, however, also important to consider the level of severity of use and - for cannabis - potency of products used, as these could theoretically affect differences in brain volume.

## CONCLUSION

This systematic review and meta-analysis, the largest to date, synthesised findings from 103 studies using rigorous methodology and comprehensive inclusion criteria. We found cross-sectional evidence that people who use cannabis had smaller volumes in the amygdala. There were smaller volumes in the amygdala, insula, and pallidum associated with tobacco use. There was consistent evidence for reductions in TGMV associated with smoking across cross-sectional, longitudinal and MR studies. Finally, this review highlights significant gaps in the literature, including a lack of studies using longitudinal and causal inference designs, as well as a lack of research on cannabis and tobacco co-use. Addressing these limitations would enhance the quality of evidence in estimating the effects of cannabis and tobacco on the brain.

## Data Availability

All data produced in the present study are available upon reasonable request to the authors

