## Supplementary file 1 for "Associations between cannabis use, tobacco use and co-use with brain volume: a systematic review and meta-analysis": Sawyer et al Supplementary File 1.docx

### Table 1. PRISMA checklist

| **Section and Topic** | **Item #** | **Checklist item** | **Location where item is reported** |
| --- | --- | --- | --- |
| **TITLE** | | |  |
| Title | 1 | Identify the report as a systematic review. | 1 |
| **ABSTRACT** | | |  |
| Abstract | 2 | See the PRISMA 2020 for Abstracts checklist. | 2 |
| **INTRODUCTION** | | |  |
| Rationale | 3 | Describe the rationale for the review in the context of existing knowledge. | 3 |
| Objectives | 4 | Provide an explicit statement of the objective(s) or question(s) the review addresses. | 4 |
| **METHODS** | | |  |
| Eligibility criteria | 5 | Specify the inclusion and exclusion criteria for the review and how studies were grouped for the syntheses. | 4 |
| Information sources | 6 | Specify all databases, registers, websites, organisations, reference lists and other sources searched or consulted to identify studies. Specify the date when each source was last searched or consulted. | 5 |
| Search strategy | 7 | Present the full search strategies for all databases, registers and websites, including any filters and limits used. | S1 |
| Selection process | 8 | Specify the methods used to decide whether a study met the inclusion criteria of the review, including how many reviewers screened each record and each report retrieved, whether they worked independently, and if applicable, details of automation tools used in the process. | 5 |
| Data collection process | 9 | Specify the methods used to collect data from reports, including how many reviewers collected data from each report, whether they worked independently, any processes for obtaining or confirming data from study investigators, and if applicable, details of automation tools used in the process. | 5 |
| Data items | 10a | List and define all outcomes for which data were sought. Specify whether all results that were compatible with each outcome domain in each study were sought (e.g. for all measures, time points, analyses), and if not, the methods used to decide which results to collect. | 5/6/S1 |
|  | 10b | List and define all other variables for which data were sought (e.g. participant and intervention characteristics, funding sources). Describe any assumptions made about any missing or unclear information. | 5 |
| Study risk of bias assessment | 11 | Specify the methods used to assess risk of bias in the included studies, including details of the tool(s) used, how many reviewers assessed each study and whether they worked independently, and if applicable, details of automation tools used in the process. | 5 |
| Effect measures | 12 | Specify for each outcome the effect measure(s) (e.g. risk ratio, mean difference) used in the synthesis or presentation of results. | 6 |
| Synthesis methods | 13a | Describe the processes used to decide which studies were eligible for each synthesis (e.g. tabulating the study intervention characteristics and comparing against the planned groups for each synthesis (item #5)). | 6 |
|  | 13b | Describe any methods required to prepare the data for presentation or synthesis, such as handling of missing summary statistics, or data conversions. | 6 |
|  | 13c | Describe any methods used to tabulate or visually display results of individual studies and syntheses. | 6 |
|  | 13d | Describe any methods used to synthesize results and provide a rationale for the choice(s). If meta-analysis was performed, describe the model(s), method(s) to identify the presence and extent of statistical heterogeneity, and software package(s) used. | 6 |
|  | 13e | Describe any methods used to explore possible causes of heterogeneity among study results (e.g. subgroup analysis, meta-regression). | 6 |
|  | 13f | Describe any sensitivity analyses conducted to assess robustness of the synthesized results. | NS |
| Reporting bias assessment | 14 | Describe any methods used to assess risk of bias due to missing results in a synthesis (arising from reporting biases). | NS |
| Certainty assessment | 15 | Describe any methods used to assess certainty (or confidence) in the body of evidence for an outcome. | 6 |
| **RESULTS** | | |  |
| Study selection | 16a | Describe the results of the search and selection process, from the number of records identified in the search to the number of studies included in the review, ideally using a flow diagram. | 6/S1 |
|  | 16b | Cite studies that might appear to meet the inclusion criteria, but which were excluded, and explain why they were excluded. | S1 |
| Study characteristics | 17 | Cite each included study and present its characteristics. | Tables 1-7 |
| Risk of bias in studies | 18 | Present assessments of risk of bias for each included study. | 7, S1 |
| Results of individual studies | 19 | For all outcomes, present, for each study: (a) summary statistics for each group (where appropriate) and (b) an effect estimate and its precision (e.g. confidence/credible interval), ideally using structured tables or plots. | S2 |
| Results of syntheses | 20a | For each synthesis, briefly summarise the characteristics and risk of bias among contributing studies. | 7 |
|  | 20b | Present results of all statistical syntheses conducted. If meta-analysis was done, present for each the summary estimate and its precision (e.g. confidence/credible interval) and measures of statistical heterogeneity. If comparing groups, describe the direction of the effect. | 7, Figures 3-7, S1 |
|  | 20c | Present results of all investigations of possible causes of heterogeneity among study results. | 7 |
|  | 20d | Present results of all sensitivity analyses conducted to assess the robustness of the synthesized results. | NA |
| Reporting biases | 21 | Present assessments of risk of bias due to missing results (arising from reporting biases) for each synthesis assessed. | 7, S2 |
| Certainty of evidence | 22 | Present assessments of certainty (or confidence) in the body of evidence for each outcome assessed. | 7 |
| **DISCUSSION** | | |  |
| Discussion | 23a | Provide a general interpretation of the results in the context of other evidence. | 9 |
|  | 23b | Discuss any limitations of the evidence included in the review. | 9/10 |
|  | 23c | Discuss any limitations of the review processes used. | 11 |
|  | 23d | Discuss implications of the results for practice, policy, and future research. | 11 |
| **OTHER INFORMATION** | | |  |
| Registration and protocol | 24a | Provide registration information for the review, including register name and registration number, or state that the review was not registered. | 4 |
|  | 24b | Indicate where the review protocol can be accessed, or state that a protocol was not prepared. | 4 |
|  | 24c | Describe and explain any amendments to information provided at registration or in the protocol. | NA |
| Support | 25 | Describe sources of financial or non-financial support for the review, and the role of the funders or sponsors in the review. | 2 |
| Competing interests | 26 | Declare any competing interests of review authors. | 2 |
| Availability of data, code and other materials | 27 | Report which of the following are publicly available and where they can be found: template data collection forms; data extracted from included studies; data used for all analyses; analytic code; any other materials used in the review. | 6 |

*From:* Page MJ, McKenzie JE, Bossuyt PM, Boutron I, Hoffmann TC, Mulrow CD, et al. The PRISMA 2020 statement: an updated guideline for reporting systematic reviews. BMJ 2021;372:n71. doi: 10.1136/bmj.n71

### Search strategy

#### Table 2: SCOPUS search strategy

| Name | Terms | Output |
| --- | --- | --- |
| Tobacco | ( TITLE-ABS-KEY ( tobacco )  OR  TITLE-ABS-KEY ( smoking )  OR  TITLE-ABS-KEY ( smok* )  OR  TITLE-ABS-KEY ( cigar* ) ) | 681,575 |
| Cannabis | ( TITLE-ABS-KEY ( cannabis )  OR  TITLE-ABS-KEY ( marijuana ) ) | 66,984 |
| Tobacco OR Cannabis | ( ( TITLE-ABS-KEY ( tobacco )  OR  TITLE-ABS-KEY ( smoking )  OR  TITLE-ABS-KEY ( smok* )  OR  TITLE-ABS-KEY ( cigar* ) ) )  OR  ( ( TITLE-ABS-KEY ( cannabis )  OR  TITLE-ABS-KEY ( marijuana ) ) ) | 728,260 |
| Brain | ( TITLE-ABS-KEY ( "brain structure" )  OR  TITLE-ABS-KEY ( "brain imaging" )  OR  TITLE-ABS-KEY ( "brain volume" )  OR  TITLE-ABS-KEY ( "magnetic resonance imaging" )  OR  TITLE-ABS-KEY ( mri )  OR  TITLE-ABS-KEY ( neuroimaging )  OR  TITLE-ABS-KEY ( "structural magnetic resonance imaging" ) ) | 1,124,113 |
| Tobacco OR Cannabis AND Brain | ( ( ( TITLE-ABS-KEY ( tobacco )  OR  TITLE-ABS-KEY ( smoking )  OR  TITLE-ABS-KEY ( smok* )  OR  TITLE-ABS-KEY ( cigar* ) ) )  OR  ( ( TITLE-ABS-KEY ( cannabis )  OR  TITLE-ABS-KEY ( marijuana ) ) ) )  AND  ( ( TITLE-ABS-KEY ( "brain structure" )  OR  TITLE-ABS-KEY ( "brain imaging" )  OR  TITLE-ABS-KEY ( "brain volume" )  OR  TITLE-ABS-KEY ( "magnetic resonance imaging" )  OR  TITLE-ABS-KEY ( mri )  OR  TITLE-ABS-KEY ( neuroimaging )  OR  TITLE-ABS-KEY ( "structural magnetic resonance imaging" ) ) ) | 13,670 – 14/09/2022 |
| Tobacco OR Cannabis AND Brain  updated search | ( ( ( TITLE-ABS-KEY ( tobacco )  OR  TITLE-ABS-KEY ( smoking )  OR  TITLE-ABS-KEY ( smok* )  OR  TITLE-ABS-KEY ( cigar* ) ) )  OR  ( ( TITLE-ABS-KEY ( cannabis )  OR  TITLE-ABS-KEY ( marijuana ) ) ) )  AND  ( ( TITLE-ABS-KEY ( "brain structure" )  OR  TITLE-ABS-KEY ( "brain imaging" )  OR  TITLE-ABS-KEY ( "brain volume" )  OR  TITLE-ABS-KEY ( "magnetic resonance imaging" )  OR  TITLE-ABS-KEY ( mri )  OR  TITLE-ABS-KEY ( neuroimaging )  OR  TITLE-ABS-KEY ( "structural magnetic resonance imaging" ) ) ) Date AFT 20220914 | 3485 05/09/2024 |

#### Table 3: PubMed search strategy

| Name | Terms | Output |
| --- | --- | --- |
| Tobacco | **(((((((tobacco[MeSH Terms]) OR (smoking[MeSH Terms])) OR (smok*[MeSH Terms])) OR (cigar*[MeSH Terms])) OR (tobacco[Title/Abstract])) OR (smoking[Title/Abstract])) OR (smok*[Title/Abstract])) OR (cigar*[Title/Abstract])** | 432,277 |
| Cannabis | **(((cannabis[MeSH Terms]) OR (marijuana[MeSH Terms])) OR (cannabis[Title/Abstract])) OR (marijuana[Title/Abstract])** | 39460 |
| Tobacco OR Cannabis | **((((((((tobacco[MeSH Terms]) OR (smoking[MeSH Terms])) OR (smok*[MeSH Terms])) OR (cigar*[MeSH Terms])) OR (tobacco[Title/Abstract])) OR (smoking[Title/Abstract])) OR (smok*[Title/Abstract])) OR (cigar*[Title/Abstract])) OR ((((cannabis[MeSH Terms]) OR (marijuana[MeSH Terms])) OR (cannabis[Title/Abstract])) OR (marijuana[Title/Abstract]))** | 458725 |
| Brain | **(((((((("magnetic resonance imaging"[MeSH Terms]) OR (neuroimaging[MeSH Terms])) OR ("brain structure"[Title/Abstract])) OR ("brain imaging"[Title/Abstract])) OR ("brain volume"[Title/Abstract])) OR ("magnetic resonance imaging"[Title/Abstract])) OR (mri[Title/Abstract])) OR (neuroimaging[Title/Abstract])) OR ("structural magnetic resonance imaging"[Title/Abstract])** | 828473 |
| Tobacco OR Cannabis AND Brain | **(((((((((tobacco[MeSH Terms]) OR (smoking[MeSH Terms])) OR (smok*[MeSH Terms])) OR (cigar*[MeSH Terms])) OR (tobacco[Title/Abstract])) OR (smoking[Title/Abstract])) OR (smok*[Title/Abstract])) OR (cigar*[Title/Abstract])) OR ((((cannabis[MeSH Terms]) OR (marijuana[MeSH Terms])) OR (cannabis[Title/Abstract])) OR (marijuana[Title/Abstract]))) AND ((((((((("magnetic resonance imaging"[MeSH Terms]) OR (neuroimaging[MeSH Terms])) OR ("brain structure"[Title/Abstract])) OR ("brain imaging"[Title/Abstract])) OR ("brain volume"[Title/Abstract])) OR ("magnetic resonance imaging"[Title/Abstract])) OR (mri[Title/Abstract])) OR (neuroimaging[Title/Abstract])) OR ("structural magnetic resonance imaging"[Title/Abstract]))** | 5555 |
| Tobacco OR Cannabis AND Brain: Humans | **(((((((((tobacco[MeSH Terms]) OR (smoking[MeSH Terms])) OR (smok*[MeSH Terms])) OR (cigar*[MeSH Terms])) OR (tobacco[Title/Abstract])) OR (smoking[Title/Abstract])) OR (smok*[Title/Abstract])) OR (cigar*[Title/Abstract])) OR ((((cannabis[MeSH Terms]) OR (marijuana[MeSH Terms])) OR (cannabis[Title/Abstract])) OR (marijuana[Title/Abstract]))) AND ((((((((("magnetic resonance imaging"[MeSH Terms]) OR (neuroimaging[MeSH Terms])) OR ("brain structure"[Title/Abstract])) OR ("brain imaging"[Title/Abstract])) OR ("brain volume"[Title/Abstract])) OR ("magnetic resonance imaging"[Title/Abstract])) OR (mri[Title/Abstract])) OR (neuroimaging[Title/Abstract])) OR ("structural magnetic resonance imaging"[Title/Abstract]))** Filters: **Humans** | 4836 14/09/2022 |
| Updated search | (("tobacco products"[MeSH Terms] OR "nicotiana"[MeSH Terms] OR "smoking"[MeSH Terms] OR "smok*"[MeSH Terms] OR "tobacco"[Title/Abstract] OR "smoking"[Title/Abstract] OR "smok*"[Title/Abstract] OR "cigar*"[Title/Abstract] OR ("cannabis"[MeSH Terms] OR "cannabis"[MeSH Terms] OR "cannabis"[Title/Abstract] OR "marijuana"[Title/Abstract])) AND ("magnetic resonance imaging"[MeSH Terms] OR "neuroimaging"[MeSH Terms] OR "brain structure"[Title/Abstract] OR "brain imaging"[Title/Abstract] OR "brain volume"[Title/Abstract] OR "magnetic resonance imaging"[Title/Abstract] OR "mri"[Title/Abstract] OR "neuroimaging"[Title/Abstract] OR "structural magnetic resonance imaging"[Title/Abstract])) AND ((humans[Filter]) AND (2022:2024[pdat])) | 751 05/09/2024 |

#### Table 4: PsycINFO search strategy

| Search | Terms | Output |
| --- | --- | --- |
| Tobacco | Index Terms: tobacco *OR* Index Terms: smoking *OR* Index Terms: smok* *OR* Index Terms: cigar* *OR* Title: tobacco *OR* Title: smoking  *OR* Title: smok* *OR* Title: cigar*  *OR* Abstract: tobacco *OR* Abstract: smoking  *OR* Abstract: smok* *OR* Abstract: cigar* | **73,567** |
| Cannabis | Index Terms: cannabis *OR* Index Terms: marijuana *OR* Title: cannabis *OR* Title: marijuana *OR* Abstract: cannabis *OR* Abstract: marijuana | **23,492** |
| Tobacco OR Cannabis | ((Index Terms: (tobacco)) *OR* (Index Terms: (smoking)) *OR* (Index Terms: (smok*)) *OR* (Index Terms: (cigar*)) *OR* (title: (tobacco)) *OR* (title: (smoking)) *OR* (title: (smok*)) *OR* (title: (cigar*)) *OR* (abstract: (tobacco)) *OR* (abstract: (smoking)) *OR* (abstract: (smok*)) *OR* (abstract: (cigar*))) *OR* ((Index Terms: (cannabis)) *OR* (Index Terms: (marijuana)) *OR* (title: (cannabis)) *OR* (title: (marijuana)) *OR* (abstract: (cannabis)) *OR* (abstract: (marijuana))) | 89,924 |
| Brain | Index Terms: "brain structure" *OR* Index Terms: "brain imaging" *OR* Index Terms: "brain volume" *OR* Index Terms: "magnetic resonance imaging" *OR* Index Terms: mri *OR* Index Terms: neuroimaging *OR* Index Terms: "structural magnetic resonance imaging" *OR* Title: "brain structure" *OR* Title: "brain imaging" *OR* Title: "brain volume" *OR* Title: "magnetic resonance imaging" *OR* Title: mri *OR* Title: neuroimaging *OR* Title: "structural magnetic resonance imaging" *OR* Abstract: "brain structure" *OR* Abstract: "brain imaging" *OR* Abstract: "brain volume" *OR* Abstract: "magnetic resonance imaging" *OR* Abstract: mri *OR* Abstract: neuroimaging *OR* Abstract: "structural magnetic resonance imaging" | 114,512 |
| Tobacco OR Cannabis AND brain | ((((Index Terms: (tobacco))) *OR* ((Index Terms: (smoking))) *OR* ((Index Terms: (smok*))) *OR* ((Index Terms: (cigar*))) *OR* ((title: (tobacco))) *OR* ((title: (smoking))) *OR* ((title: (smok*))) *OR* ((title: (cigar*))) *OR* ((abstract: (tobacco))) *OR* ((abstract: (smoking))) *OR* ((abstract: (smok*))) *OR* ((abstract: (cigar*)))) *OR* (((Index Terms: (cannabis))) *OR* ((Index Terms: (marijuana))) *OR* ((title: (cannabis))) *OR* ((title: (marijuana))) *OR* ((abstract: (cannabis))) *OR* ((abstract: (marijuana))))) *AND* ((Index Terms: ("brain structure")) *OR* (Index Terms: ("brain imaging")) *OR* (Index Terms: ("brain volume")) *OR* (Index Terms: ("magnetic resonance imaging")) *OR* (Index Terms: (mri)) *OR* (Index Terms: (neuroimaging)) *OR* (Index Terms: ("structural magnetic resonance imaging")) *OR* (title: ("brain structure")) *OR* (title: ("brain imaging")) *OR* (title: ("brain volume")) *OR* (title: ("magnetic resonance imaging")) *OR* (title: (mri)) *OR* (title: (neuroimaging)) *OR* (title: ("structural magnetic resonance imaging")) *OR* (abstract: ("brain structure")) *OR* (abstract: ("brain imaging")) *OR* (abstract: ("brain volume")) *OR* (abstract: ("magnetic resonance imaging")) *OR* (abstract: (mri)) *OR* (abstract: (neuroimaging)) *OR* (abstract: ("structural magnetic resonance imaging"))) | 1703 14/09/2022 |
| Updated search | ((((Index Terms: (tobacco))) *OR* ((Index Terms: (smoking))) *OR* ((Index Terms: (smok*))) *OR* ((Index Terms: (cigar*))) *OR* ((title: (tobacco))) *OR* ((title: (smoking))) *OR* ((title: (smok*))) *OR* ((title: (cigar*))) *OR* ((abstract: (tobacco))) *OR* ((abstract: (smoking))) *OR* ((abstract: (smok*))) *OR* ((abstract: (cigar*)))) *OR* (((Index Terms: (cannabis))) *OR* ((Index Terms: (marijuana))) *OR* ((title: (cannabis))) *OR* ((title: (marijuana))) *OR* ((abstract: (cannabis))) *OR* ((abstract: (marijuana))))) *AND* ((Index Terms: ("brain structure")) *OR* (Index Terms: ("brain imaging")) *OR* (Index Terms: ("brain volume")) *OR* (Index Terms: ("magnetic resonance imaging")) *OR* (Index Terms: (mri)) *OR* (Index Terms: (neuroimaging)) *OR* (Index Terms: ("structural magnetic resonance imaging")) *OR* (title: ("brain structure")) *OR* (title: ("brain imaging")) *OR* (title: ("brain volume")) *OR* (title: ("magnetic resonance imaging")) *OR* (title: (mri)) *OR* (title: (neuroimaging)) *OR* (title: ("structural magnetic resonance imaging")) *OR* (abstract: ("brain structure")) *OR* (abstract: ("brain imaging")) *OR* (abstract: ("brain volume")) *OR* (abstract: ("magnetic resonance imaging")) *OR* (abstract: (mri)) *OR* (abstract: (neuroimaging)) *OR* (abstract: ("structural magnetic resonance imaging"))) *AND* Year: 2021 *To* 2024 | 366 05/09/2024 |

### Methods: additional information

Data extraction:
When the same outcome was reported from multiple studies with overlapping samples, studies were included according to the following criteria, in order: 1) the study with the largest sample size, 2) the study with the lowest risk of bias; if these were the same across studies, the oldest study was included. For studies which reported outcomes for females and males separately, or stratified by age (e.g., young vs old), data were extracted separately where data for the whole sample was not available.

Where studies reported multiple exposure groups, and where it was not possible to include all user/non-user comparisons for each exposure group, the exposure group that was less likely to be confounded (e.g. no or fewer comorbid conditions or drug use), and the non-using group which most closely matched the exposure group, were selected. For example, if a study reported cannabis use in a sample with schizophrenia, with both a non-using schizophrenia group and a non-using non-schizophrenia group, the control group would be the non-using schizophrenia group as this more closely matches the exposure group. Similarly, if a study reported a cannabis and methamphetamine group and a cannabis only group compared with a drug-free control group, the cannabis only group would be selected.

Meta-analysis:
If a study reported regional volumes split by hemisphere, Hedges’ g values were aggregated across left and right hemispheres using the “aggregate” function from the metafor package in R, with a correlation of 0.8 and inverse-variance weighting (1,2). A correlation of 0.8 was used, based on the correlation of left and right estimates reported in the meta-analysis by Rocchetti et al., (3).

Meta-analyses were split into adjusted and unadjusted estimates for each outcome. Adjusted analyses included studies where the comparison of group differences was statistically adjusted for potential confounders, or means adjusted, for example by intracranial volume (ICV; either in the MRI pipeline or statistically). Unadjusted analysis included studies which only matched groups on key confounders (e.g., age and/or sex) or reported raw volumes (i.e., not adjusted by ICV or any other variable in either the MRI pipeline or statistically). If a study reported both adjusted and unadjusted data, it was included in both the adjusted and unadjusted analyses.

Methods references:

1.	Harrer M, Cuijpers P, Furukawa TA, Ebert DD. Doing meta-analysis with R: a hands-on guide. First edition. Boca Raton London New York: CRC Press; 2022. 474 p.

2.	Viechtbauer W. metafor: Meta-Analysis Package for R [Internet]. 2009 [cited 2024 Aug 7]. p. 4.6-0. Available from: https://CRAN.R-project.org/package=metafor

3.	Rocchetti M, Crescini A, Borgwardt S, Caverzasi E, Politi P, Atakan Z, et al. Is cannabis neurotoxic for the healthy brain? A meta‐analytical review of structural brain alterations in non‐psychotic users. Psychiatry Clin Neurosci. 2013 Nov;67(7):483–92.

### Table 5: Newcastle Ottawa Scale cross-sectional risk of bias template

| **ROB component** | **Options** | **Justification for adaptation** |
| --- | --- | --- |
| 1) Representativeness of the sample | a) *Truly representative of the average in the target population. (all subjects or random sampling)  b) *Somewhat representative of the average in the target group. (non-random sampling)  c) Selected group of users/convenience sample.  d) No description of the derivation of the included subjects. | N/A |
| 2) Sample size | a) *Justified and satisfactory (including sample size calculation).  b) Not justified.  c) No information provided | N/A |
| 3) Non-respondents | a) *Proportion of target sample recruited attains pre-specified target or basic summary of non-respondent characteristics in sampling frame recorded.  b) Unsatisfactory recruitment rate, no summary data on non-respondents.  c) No information provided | N/A |
| 4) Ascertainment of exposure | a) **secure record (Laboratory sample test, e.g. CO/saliva samples for tobacco, urine sample for THC)  b) *self-report using detailed descriptions (e.g. smoke every day for last month) or self-report from participant living in a place where cannabis/tobacco is legal  c) self-report only  d) no description | This has been edited to distinguish between studies which assess detail substance use using self-report questions, and including frequency/amount in their descriptives, versus studies which use limited questioning to assess use (e.g. yes/no). The legalisation criteria was added to account for differences where people may be more accurate in describing use in legal countries, as there may be more available information about the substance and/or participants may be more inclined to be truthful about use. |
| Comparability |  |  |
| 1) Did the study account for other drug use? (e.g. in screening criteria, matched groups, adjustment, or report differences and there are no difference) | a) **alcohol and tobacco/cannabis AND any other drug use  b) *alcohol and tobacco/cannabis  c) No or Unclear | This criterion assessed whether studies were investigating the specific effects of cannabis/tobacco and considering potential co-occurrence of other drug use. Alcohol, and either tobacco/cannabis are commonly used alongside each other, so studies are given a star if they account for this. Additional stars are given to account for any other drug use as the additional effects of these substance on the brain are often not considered, hence studies are recognised for accounting for this. |
| 2) FOR CANNABIS/CO-USE ONLY: Study described route of administration, e.g., co-administration with tobacco in a joint. Studies must include information on co-administration with tobacco to achieve the highest mark. | a) *Yes  b) No or unclear | Cannabis is often co-used with tobacco, which can vary by region, and specific mixing of cannabis with tobacco is often not reported or assessed. Studies which attempted to describe the specific administration of cannabis used are recognised for this detail, and such consideration of confounding effects of tobacco use. |
| 3) Study matches groups or controls for Age | a) *Yes  b) No or unclear | A key confounder in investigations of brain volume. |
| 4) Study matches groups or controls for sex or gender | a) *Yes  b) No or unclear | A key confounder in investigations of brain volume. |
| Outcome |  |  |
| 1) Assessment of outcome | a) *Automated brain tissue volume computations (e.g. SPM, FreeSurfer)  b) *Manual tracing blinded  c) Manual tracing unblinded  d) No description | Whether automatic or manual segmentations are more valid is a contested issue, therefore this review gives the same scores for automatic and blinded manual tracings. Unblinded manual tracings are not given a star due to the potential for biased assessments. |
| 2) Statistical test | a) *Statistical test used to analyse the data clearly described, appropriate and measures of association presented, and unit of ROI measure reported  b) Statistical test not appropriate, not described or incomplete. | Minimal edits, more description is provided for reviewers. |

### Table 6: Newcastle Ottawa Scale longitudinal risk of bias template

| **ROB component** | **Options** | **Justification for adaptation** |
| --- | --- | --- |
| 1) Representativeness of the sample | a) *Truly representative of the average in the target population. (all subjects or random sampling)  b) *Somewhat representative of the average in the target group. (non-random sampling)  c) Selected group of users/convenience sample.  d) No description of the derivation of the included subjects. | N/A |
| 2) Sample size | a) *Justified and satisfactory (including sample size calculation).  b) Not justified.  c) No information provided | N/A |
| 3) Selection of the non-exposed cohort | a) *drawn from the same community as the exposed cohort  b) drawn from a different source  c) no description of the derivation of the non-exposed cohort | N/A |
| 4) Ascertainment of exposure | a) **secure record (Laboratory sample test, e.g. CO/saliva samples for tobacco, urine sample for THC)  b) *self-report using detailed descriptions (e.g. smoke every day for last month) or self-report from participant living in a place where cannabis/tobacco is legal  c) self-report only  d) no description | This has been edited to distinguish between studies which assess detail substance use using self-report questions, and including frequency/amount in their descriptives, versus studies which use limited questioning to assess use (e.g. yes/no). The legalisation criteria were added to account for differences where people may be more accurate in describing use in legal countries, as there may be more available information about the substance and/or participants may be more inclined to be truthful about use. |
| 5) Demonstration that outcome of interest (brain differences) was not present at start of study | a) **At baseline the whole sample non-users - brain differences not present at the start of the study  b) *Baseline measure and follow up measure of brain volume, change calculated or baseline values adjusted for  c) unclear or No - only took outcome measure at follow up, or only measured exposure at follow up | N/A |
| Comparability |  |  |
| 1) Did the study account for other drug use? (e.g. in screening criteria, matched groups, adjustment, or report differences and there are no difference) | a) **alcohol and tobacco/cannabis AND any other drug use  b) *alcohol and tobacco/cannabis  c) No or Unclear | This criterion assessed whether studies were investigating the specific effects of cannabis/tobacco and considering potential co-occurrence of other drug use. Alcohol, and either tobacco/cannabis are commonly used alongside each other, so studies are given a star if they account for this. Additional stars are given to account for any other drug use as the additional effects of these substance on the brain are often not considered, hence studies are recognised for accounting for this. |
| 2) FOR CANNABIS/CO-USE ONLY: Study described route of administration, e.g., co-administration with tobacco in a joint. Studies must include information on co-administration with tobacco to achieve the highest mark. | a) *Yes  b) No or unclear | Cannabis is often co-used with tobacco, which can vary by region, and specific mixing of cannabis with tobacco is often not reported or assessed. Studies which attempted to describe the specific administration of cannabis used are recognised for this detail, and such consideration of confounding effects of tobacco use. |
| 3) Study matches groups or controls for Age | a) *Yes  b) No or unclear | A key confounder in investigations of brain volume. |
| 4) Study matches groups or controls for sex or gender | a) *Yes  b) No or unclear | A key confounder in investigations of brain volume. |
| Outcome |  |  |
| 1) Assessment of outcome | a) *Automated brain tissue volume computations (e.g. SPM, FreeSurfer)  b) *Manual tracing blinded  c) Manual tracing unblinded  d) no description | Whether automatic or manual segmentations are more valid is a contested issue, therefore this review gives the same scores for automatic and blinded manual tracings. Unblinded manual tracings are not given a star due to the potential for biased assessments. |
| 2) Is the temporal relationship between exposure (current use) /outcome (changes in brain volume) feasible? | a) *Yes  b) No or unclear | N/A |
| 3) Adequacy of follow up of cohorts | a) *complete follow up - all subjects accounted for  b) *subjects lost to follow up unlikely to introduce bias - small number lost (<20%)  c) follow up rate < 80% and no description of those lost  d) no statement | N/A |
| 4) Statistical test | a) *Statistical test used to analyse the data clearly described, appropriate and measures of association presented, and unit of ROI measure reported  b) Statistical test not appropriate, not described or incomplete. | N/A |

Figure 1: PRISMA flow diagram

**Records identified on 14/09/22 from:**

SCOPUS (n = 17153)

PubMed (n = 5587)

PsycINFO (n = 2069)

Additional sources (n = 2)

Total (n = 24811)

Records removed *before screening*:

Duplicate records removed (n = 6517)

Records marked as ineligible by automation tools (n = 0)

Records removed for other reasons (n = 0)

**Records screened**

**(n = 18294)**

Records excluded**

(n = 17920)

**Reports sought for retrieval**

**(n = 374)**

Reports not retrieved

(n = 0)

**Reports assessed for eligibility**

**(n = 374)**

**Reports excluded: (n = 271)**

Wrong exposure (n = 99)

Wrong outcomes (n = 116)

Wrong comparator (n = 15)

Wrong study design (n = 10)

Unable to obtain full text (n = 5)

Wrong method (n = 8)

Overlapping samples (n = 18)

**Independent studies included in review**

**(n = 103)**

**Co-use: (n = 1)**

**Cannabis: (n = 57)**

Cross-sectional (n = 50)

Longitudinal (n = 7)

MR (n = 0)

**Tobacco: (n = 45)**

Cross-sectional (n = 40)

Longitudinal (n = 4)^+^

MR (n = 2)

*^+^ 1 tobacco study included cross-sectional and longitudinal analysis so is counted twice here*

*MR = Mendelian Randomisation studies*

**Identification of studies via databases and registers**

**Identification**

**Screening**

**Included**

**Studies included in meta-analysis across type and region (77):**

**Cannabis: (n = 44)**

Cross-sectional (n = 44)

Longitudinal (n = 0)

**Tobacco: (n = 33)**

Cross-sectional (n = 30)

Longitudinal (n = 3)

**Meta-analysis**

### Table 7: MRI parameters for cross-sectional cannabis studies included in meta-analysis

| **Study ID** | **MRI Magnet: Tesla** | **MRI Magnet: Brand** | **Acquisition Sequence** | **Acquisition Duration** | **Voxel size, mm/slice thickness** | **Repetition time TR, ms** | **Echo time TE, ms** | **Flip angle °** | **Field of View FOV mm** | **Matrix size** | **Technique for segmentation (automated, semi automated or manual)** | **Programme used for analysis (SPM/freesurfer)** |
| --- | --- | --- | --- | --- | --- | --- | --- | --- | --- | --- | --- | --- |
| Ashtari 2011 | 1.5 | General Electric | 3D SPGR | 7:44 min | NS | 10.1 | 4.2 | NS | 242x22 cm2 | 256x192 | Semi-automated | ITK-SNAP, ANTS |
| Batalla 2018 | 1.5 | General Electric | IR-FSPGR | NS | 1 mm isotropic | 11.8 | 4.2 | 15 | 300 | 256 x 256 | Semi-automated | Linux |
| Block 2000 | 1.5 | GE Signa | SGRASS | NS | NS | 24 | 5 | 40 | 260 | 256x192 | Semi-automated | BRAINS |
| Buchy 2016 | 3 / 1.5 | Siemens/GE HDx/GE Discovery | MPRAGE/IS-SPGR | NS | 1x1, 1.2mm slice thickness | 2300/7.0 | 2.91 | 9/8 | 256x240x176/26cm | NS | Automated | FreeSurfer |
| Cahn 2004 | 1.5 | Philips | NS | NS | 1.2 (slice thickness) | 30 | 4.6 | 30 | 256 | NS | Semi-automated | NS |
| Churchwell 2010 | 3 | Siemens | 3D MPRAGE | NS | NS | 20 | 3.38 | 8 | 256 | NS | Semi-automated | FreeSurfer |
| Churchwell 2012 | 3 | Siemens | MPRAGE | NS | 1.0 (slice thickness) | 3 | 3.38 | 8 | 256 | NS | Semi-automated | FreeSurfer |
| Chye 2017b | 3 | Siemens | High resolution MPRAGE | NS | 0.5x0.5x1 | 1900 | 2.15 | NS | 256 | NS | Automated | FreeSurfer |
| Cohen 2012 | 1.5 | Siemens | MPRAGE | NS | 1 x 1 x 1 mm | 9.7 | 4 | 12 | NS | NS | Combination of manual, semi-automated, and automated steps | NS |
| D'Souza 2021 | 3 | Siemens | MPRAGE | NS | 0.98 x 0.98 x 1 | 2500 | 3.34 | 7 | NS | NS | Automated | FreeSurfer |
| Gilman 2014 | 3 | Siemens | 3D MPRAGE | 6′03′′ | 1x1x1 | 2.5 | 1.64, 3.5,  5.36, 7.22 | NS | 256 | NS | Automated | FSL-VBM |
| James 2011 | 1.5 | Siemens | 3D T1-weighted FLASH | NS | 1mm slice thickness | 12 | 5.6 | 19 | NS | 256 x 256 | Automated | FSL FAST |
| Knodt 2022 | 3 | Siemens | MPRAGE | total scan time: 6 minutes and 52 seconds | slice thickness = 0.9 mm with no gap (voxel size 0.9×0.875×0.875 mm) | 2400 | 1.98 | 9 | 224mm | 256x256 | Automated | FreeSurfer |
| Koenders 2015 | 3 | Philips | NS | NS | 1.0 or 1.2 (slice thickness) | 9.6 (mean) | 4.4 (mean) | 8 | NS | NS | Automated | Freesurfer |
| Koenis 2021 | 3 | Siemens | GRE | 7:02 min | 0.8mm isotropic | 2400 | 2.09 | 8 | NS | NS | Automated | FreeSurfer |
| Kumra 2012 | 3 | Siemens | MPRAGE | NS | NS | 2530 | 3.65 | 7 | 256x176 | NS | Semi-automated | FreeSurfer |
| Levar 2018 | 3 | Siemens | MPRAGE | NS | 1 mm isotropic | 2530 | 1.64/3.5/ 5.26/7.22 | NS | 256 | NS | Automated | Freesurfer |
| Lopez-Larson 2011 | 3 | Siemens | 3D MPRAGE | NS | 1×1×1* | 20 | 3.38 | 8 | 256 | 256×256 | Automated | FreeSurfer |
| Lorenzetti 2020 | 3 | Siemens | MPRAGE | NS | 1 mm isotropic | 1900 | 2.15 | NS | 256 | NS | Semi-automated | Manual or FreeSurfer |
| Lorenzetti 2024 site 1 | 3 | NS | TFE | NS | 1x1x1.2 | 9.6 | 4.6 | 8 | NS | 256x256x182 | Automated | FreeSurfer |
| Lorenzetti 2024 site 2 | 1.5 | NS | FSGIR | NS | 1.17x1.17x1.2 | 11.8 | 4.2 | 15 | NS | 256x256x124 | Automated | FreeSurfer |
| Lorenzetti 2024 site 3 | 3 | NS | MPRAGE | NS | 1x1x1.1 | 1900 | 2.15 | 12 | NS | 256x256x176 | Automated | FreeSurfer |
| Mashhoon 2015 | 3 | Siemens | MPRAGE | NS | 1 x 1 x 1.3 mm3 | 2100 | 2.7 | 12 | NS | 256 x 256 | Automated | Freesurfer |
| Mata 2010 | 1.5 | GE Signa | SPGR | 14:48 min | NS | 24 | 5 | rotation: 45 | 260x190 | 256x192 | Automated | BRAINS2 |
| McQueeny 2011 | 3 | GE | 3D SPGR | 7′19′′ | NS | 8 | 3 | 12 | 240 | NS | Semi-automated | AFNI, manual tracings blinded |
| Medina 2007a | 1.5 | GE Signa | 3D FSE | 8′36′′ | 0.94×0.94×1.3 | 20 | 16 | NS | 240 | NS | Semi-automated | AFNI, manual tracings blinded |
| Medina 2007b | 1.5 | GE Signa | 3D FSE | 8′36′′ | 0.94×0.94×1.3 | 20 | 16 | NS | 240 | NS | Semi-automated | FAST, AFNI, manual blinded |
| Medina 2009 | 1.5 | GE Signa | 3D FSE | 08:36 | 0.9375 × 0.9375 × 1.328 | 2000 | 16 | NS | 240 | NS | semi-automated | FAST, AFNI |
| Medina 2010 | 1.5 | GE Signa | 3D FSE | 8′36′′ | 0.94×0.94×1.3 | 20 | 16 | NS | 240 | NS | Semi-automated | AFNI |
| Meier 2019 | 3 | Siemens | SPGR | NS | 0.8 mm (slice thickness) | 1630 | 2.48 | 8 | 204 | 256 x 256 | Automated | Freesurfer |
| Meier 2022 | 3 | Siemens | MPRAGE | total scan time: 6 minutes and 52 seconds | slice thickness = 0.9 mm with no gap (voxel size 0.9×0.875×0.875 mm) | 2400 | 1.98 | 9 | 224mm | 256x256 | Automated | FreeSurfer |
| Moreno-Alcazar 2018 | 1.5 | GE Signa | NS | NS | 0.47x0.47x1mm3 | 2000 | 3.93 | 15 | NS | 512x512 | Automated | FIRST |
| Navarri 2022 | NS | NS | NS | NS | NS | NS | NS | NS | NS | NS | Automated | FreeSurfer |
| Owens 2021 | 3 | Siemens | NS | NS | 0.7mm3 isotropic | 2400 | 2.14 | NS | 224x240 | 320x320 | Automated | FreeSurfer |
| Price 2015 | 4 | Varian | 3D MDEFT | 15′ | 1×1×1* | 13 | 6 | 20 | 256 | 192×96 | Semi-automated | FreeSurfer |
| Radoman 2019 | 3 | Siemens | MPRAGE | NS | 1 mm isotropic | 2.53 | 1.64/3.5/ 5.36/7.22 | NS | 256 | NS | Automated | Freesurfer |
| Romero 2015 | 3 | Siemens | NS | NS | 1 mm (slice thickness) | 8.1 | 3.2 | 8 | 220 | NS | Automated | SPM8, VBM8 |
| Rosetti 2021 | NS | NS | NS | NS | NS | NS | NS | NS | NS | NS | Automated | FreeSurfer |
| Schacht 2012 | 3 | Siemens | MPRAGE | 6 | 1x1x1 | 2300 | 2.74 | 81 | 256x256 | 256x256x176 | Automated | FreeSurfer |
| Scheffler 2021 | 3 | Siemens | MPRAGE | NS | 0.9x0.9x1 | 2080 | 4.88 | NS | 230 | NS | Automatic | FreeSurfer |
| Scheffler 2021 | 3 | Siemens | ME-MPRAGE | NS | 1x1x1 | 2530 | 1.53/3.21/ 4.89/6.57 | 7 | 256 | NS | Automatic | FreeSurfer |
| Scheffler 2021 | 3 | Siemens | ME-MPRAGE | NS | 1x1x1 | 2530 | 1.63/3.47/ 5.31/7.15 | 7 | 280 | NS | Automatic | FreeSurfer |
| Thayer 2019a | 3 | Siemens | MPRAGE | NS | 0.8 x 0.8 x 0.8 mm | 2400 | 2.07 | 8 | 256 x 256 | NS | Automated | FSL, Freesurfer |
| Tzilos 2005 | 1.5 | GE Signa | SPGR | NS | 0.976x0.976 | 35 | 5 | 45 | NS | 256x256 | Semi-automated | MRX |
| Vered 2024 | 3 | Siemens | MPRAGE | NS | NS | NS | NS | NS | NS | NS | NS | NS |
| Wallace 2024 | NS | GE, Siemens or Phillips |  | 5:38-7:12 min | 1x1x1 | 6.31-2500 | 2-2.9 | 8 | 256x240-256 | 256x256 | Automated | FreeSurfer |
| Weiland 2015 | 3 | Siemens | 3D MPRAGE | NS | 1×1×1 | 2.5 | 1.64, 3.5, 5.36, 7.22, 9.08 | 7 | 256 | 256×256 | Automated | FreeSurfer |
| Yip 2014 | 3 | Siemens | MPRAGE | NS | NS | 2530 | 3.34 | 7 | 256x256 | 256x256 | Semi-automated | FSL |

### Table 8: MRI parameters for cross-sectional tobacco studies included in meta-analysis

| **Study ID** | **MRI Magnet: Tesla** | **MRI Magnet: Brand** | **Acquisition Sequence** | **Acquisition: Duration** | **Voxel size, mm/slice thickness** | **Repetition time TR, ms** | **Echo time TE, ms** | **Flip angle °** | **Field of View FOV mm** | **Matrix size** | **Technique for segmentation (automated, semi automated or manual)** | **Programme used for analysis (SPM/freesurfer)** |
| --- | --- | --- | --- | --- | --- | --- | --- | --- | --- | --- | --- | --- |
| Austin 2022 | 3 | Siemens | 3D (isotropic sagittal ??) | NS | 1 (slice thickness) | 1900 | 2.93 | 9 | 250 | 176 slices | Automated | NS |
| Binnewies 2023 - Base II | 3 | Siemens | NS | NS | 1.0mm slice thickness | 2500 | 4.77 | 7 | 256x256 | NS | Automated | FreeSurfer |
| Binnewies 2023 - Betula | 3 | Discovery GE | NS | NS | 1.0mm slice thickness | 8.19 | 3.2 | 12 | 250x250 | NS | Automated | FreeSurfer |
| Binnewies 2023 - Cam-CAN | 3 | Siemens | NS | NS | 1.0mm slice thickness | 2250 | 2.98 | 9 | 256x240 | NS | Automated | FreeSurfer |
| Binnewies 2023 - LCBC | 1.5 | Siemens | NS | NS | 1.2mm slice thickness | 2500 | 3.61 | 8 | 240x240 | NS | Automated | FreeSurfer |
| Binnewies 2023 - LCBC | 1.5 | Siemens | NS | NS | 1.2mm slice thickness | 2400 | 3.79 | 8 | 240x240 | NS | Automated | FreeSurfer |
| Binnewies 2023 - LCBC | 3 | Siemens | NS | NS | 1.0mm slice thickness | 2300 | 2.98 | 8 | 256x256 | NS | Automated | FreeSurfer |
| Binnewies 2023 - LCBC | 3 | Siemens | NS | NS | 0.8mm slice thickness | 2400 | 2.22 | 8 | 240x256 | NS | Automated | FreeSurfer |
| Binnewies 2023 - LISA | 3 | Phillips | NS | NS | 0.85mm3 | 9.3 | 2.7 | 8 | NS | 288x288 | Automated | FreeSurfer |
| Binnewies 2023 - MOTAR | 3 | Phillips | NS | NS | 1.0mm slice thickness | 9 | 3.5 | NS | 256x256 | NS | Automated | FreeSurfer |
| Binnewies 2023 - NESDA | 3 | Phillips | NS | NS | 1.0mm slice thickness | 9 | 3.5 | NS | NS | NS | Automated | FreeSurfer |
| Binnewies 2023 - Whitehall-II | 3 | Siemens | NS | NS | 1.0mm slice thickness | 2530 | 1.79/3.65/ 5.51/7.37 | 7 | 256x256 | NS | Automated | FreeSurfer |
| Binnewies 2023 - Whitehall-II | 3 | Siemens | NS | NS | 1.0mm slice thickness | 1900 | 3.97 | 8 | 192x192 | NS | Automated | FreeSurfer |
| Brody 2004 | 1.5 | Siemens | 3DFT SPGR | NS | 1.5mm slices | 30 | 7 | 30 | 230 | 256x192 | Manual | Manual tracing |
| Cardenas 2020 | 4 | Siemens | 3D MPRAGE | NS | 1.0 isotropic resolution | 2300 | 3 | 7 | NS | NS | Automated | FreeSurfer |
| Chen 2023 | 3T | NS | NS | NS | NS | NS | NS | NS | NS | NS | NS | FreeSurfer |
| Cho 2016 | NS | NS | NS | NS | NS | NS | NS | NS | NS | NS | Automated | Montreal Neurological Institute image processing software (CIVET) |
| Choi 2010 | 3 | FORTE | MPRAGE | NS | 1.5mm slices | 10 | 4 |  | 220x192x192 | s56x224x128 | Manual | Brain Voyager |
| Chye 2020 - Tang 2020 | NS | NS | MPRAGE | NS | NS | 2300 | 2.98 | 9 | NS | NS | Automated | FreeSurfer |
| Chye 2020 - Claus 2013 | NS | NS | MPRAGE | NS | NS | 2530 | 1.64, 3.5, 5.36, 7.22, 9.08 | 7 | NS | NS | Automated | FreeSurfer |
| Chye 2020 - Wilcox 2017 | NS | NS | MPRAGE | NS | NS | 2300 | 2.74 | 8 | NS | NS | Automated | FreeSurfer |
| Chye 2020 - Morales 2014 | NS | NS | MPRAGE | NS | NS | 1900-2530 | 2.26-3.31 | 7-9 | NS | NS | Automated | FreeSurfer |
| Chye 2020 - Zhang 2011 | NS | NS | MPRAGE | NS | NS | 2500 | 4.38 | 8 | NS | NS | Automated | FreeSurfer |
| Chye 2020 - Luijten 2012 | NS | NS | FSPGR | NS | NS | 10.6 | 2.2 | NS | NS | NS | Automated | FreeSurfer |
| Durazzo 2007a | 1.5 | Siemens | MPRAGE | NS | 1x1mm2 1.5mm slice thickness | 9.7 | 4 | NS | NS | NS | Automated: but reviewed by trained operator | NS |
| Durazzo 2013a | 4 | Siemens | MPRAGE | NS | 1x1x1 | 2300 | 3 | 7 |  | NS | Automated | FreeSurfer |
| Durazzo 2013b | 1.5 | Siemens | MPRAGE | NS | 1.5mm (coronal partitions) | 9.7 | 4 | 15 | NS | NS | Automated | FreeSurfer |
| Durazzo 2017 | 4 | Siemens | MPRAGE | NS | 1.0mm3 | 2300 | 3 | 7 | NS | NS | Automated | FreeSurfer |
| Durhan 2016 | 3 | Siemens | MPRAGE | TA: 4.06? | 0.9mm3 | 2600 | 3.1 | NS | NS | 224x256 | Automated | FreeSurfer |
| Duriez 2014 - cross-sectional | 1.5 | Siemens | 3D SPGR | TI = 600ms | 1.0x0.98 x0.98 | 9.7 | 4 | NS | NS | 256x192x256 | Automated | SPM99 |
| Elbejjani 2019 | 3 | Siemens or Philips | MPRAGE | NS | 1 mm (slice thickness) | 1900 | 2.89 | 9 | 250 | 256 x 256 | Automated | NS |
| Gazdzinksi 2005 | 1.5 | Siemens | MPRAGE | NS | 1x1 1.5mm (slabs) | 9.7 | 4 | NS | NS | NS | Semi-automated | NS |
| Janowitz 2014 | 1.5 | Siemens | Multiplanar reconstruction | NS | 1.0x1.0x1.0mm | 1900 | 3.4 | 15 | NS | NS | Automated | FreeSurfer |
| Launer 2015 | 3 | Siemens or Philips | MPRAGE | NS | 1 mm (slice thickness) | 1900 | 2.89 | 9 | 250 | 256 x 256 | Automated | NS |
| Li 2015 | 3 | General Electric | NS | NS | 1mm isotropic | 1900 | 2.26 | 9 | 256 x 256 | 256 x 256 | Automated | Freesurfer |
| Liang 2022 | 3 | Siemens | MPRAGE | NS | 1mm slice thickness | 2200 | 4.47 | 12 | 256 | 256x256 | Automated | FSL, FreeSurfer |
| Lie 2022 | 3/1.5 | NS | MPRAGE | NS | 1x1x1 | 1800 | 2.28 | 8 | NS | NS | Automated | FreeSurfer |
| Lin 2021 | 3 | GE Signa | FSPGR | NS | 1mm slice thickness | 6.1 | 2.8 | 15 | 256x256 | 256x256 | Automated | FreeSurfer |
| Linli 2023 | 3 | Siemens | NS | NS | 1x1x1 | NS | NS | NS | 208x256x256 | NS | Automated | CAT12 |
| Luhar 2013 | 3 | Siemens | MPRAGE | NS | 1mm slice thickness | 2530 | 1.79 - 7.55 (RMS average used) | 7 | 256 | 256 x 256 | Automated | FreeSurfer |
| Rinigin 2022 | 1.5 | Siemens | MPRAGE | NS | 0.98x0.98x1 | 1980 | 4.3 | NS | 250x250 | 256x256 | Automated | FreeSurfer |
| Shang 2024 | 3 | Siemens | NS | NS | NS | NS | NS | NS | NS | NS | Semi-automated | Functional MRI of the Brain Software Library |
| Shen 2017 | 3.0T | GE Signa | NS | NS | NS | 2000 | 30 ms | 80 | 240 × 240 mm2 | 64 × 64 | Automatic | Freesurfer |
| Wang 2020 | 3T | GE Signa | FSGPR | NS | 1.2mm slice thickness | 5.056 | 1.116 | 15 | 240x240 | 256x256 | Automated | FreeSurfer |
| Yu 2018 | 3 | GE Signa | NS | NS | 1 mm isotropic | 8.5 | 3.4 | 12 | 240 x 240 | 240 x 240 | Automated | Freesurfer |
| Yuan 2016 | 3T | GE Signa | NS | NS | NS | 8.5 | 3.4 | 12 | 240 × 240 | 240 × 240 | Automatic | Freesurfer |

### Table 9: MRI parameters for cross-sectional co-use studies

| **Study ID** | **MRI Magnet: Tesla** | **MRI Magnet: Brand** | **Acquisition: Sequence** | **Acquisition: Duration** | **Voxel size, mm/slice thickness** | **Repetition time TR, ms** | **Echo time TE, ms** | **Flip angle °** | **Field of View FOV mm** | **Matrix size** | **Technique for segmentation (automated, semi automated or manual)** | **Programme used for analysis (SPM/freesurfer)** |
| --- | --- | --- | --- | --- | --- | --- | --- | --- | --- | --- | --- | --- |
| Filbey 2015 | 3 | Siemens Tim Trio | MPRAGE | 6 min | 1x1x1 | 2300 | 2.74 | 8 | 256x256 | NS | Automated | FreeSurfer |

### Table 10: MRI parameters for longitudinal cannabis studies

| **Study ID** | **MRI Magnet: Tesla** | **MRI Magnet: Brand** | **Acquisition: Sequence** | **Acquisition: Duration** | **Voxel size, mm/slice thickness** | **Repetition time TR, ms** | **Echo time TE, ms** | **Flip angle °** | **Field of View FOV mm** | **Matrix size** | **Technique for segmentation (automated, semi automated or manual)** | **Programme used for analysis (SPM/freesurfer)** |
| --- | --- | --- | --- | --- | --- | --- | --- | --- | --- | --- | --- | --- |
| Garimella 2020 | 3 | Intera, Philips | NS | NS | 1.2mm (slice thickness) | 9600 | 4600 | 8 | 256x256 | 256x256 | Automated | FreeSurfer |
| Koenders 2017 | 3 | Philips | TFE | NS | 1.2mm (slice thickness) | 9.6 | 4.6 | 8 | 256x256 | 256x256 | Manual tracings | SPM/FLIRT |
| Luo 2022 | 3 | GE | IR-SPGR | NS | 1.2x0.9x0.9 slice thickness | 5.904 | 1.932 | 11 | 240 | 256x256 | Automated | NS |
| Luo 2022 | 3 | Siemens | MPRAGE | NS | 1.2x0.9x0.9 slice thickness | 1900 | 2.92 | 9 | 240 | 256x256 | Automated | NS |
| Rais 2008 | 1.5 | Philips | FFE | NS | 1.2mm slice thickness | 30 | 4.6 | 30 | 256 x 256 | NS | Manual | N/A |
| Wang 2021 | 3 | Philips | NS | NS | 1.2mm (slice thickness) | 9600.00 | 4600.00 | 8 | 256X256 | 256X256 | Automated | volBrain |
| Welch 2011 | 1 | Siemens | MPRAGE | TI = 200ms | 1.88mm slice thickness | 10 | 4 | 12 | 250x250mm | NS | Semi-automated | Analyze for UNIX, manual tracing |
| Xu 2022 | 3 | Philips | NS | NS | 1x1x1.22 | 9600 | 4160 | 8 | 256x256 | 256x256 | Automated | FSL, SINEAX |

### Table 11: MRI parameters for longitudinal tobacco studies

| **Study ID** | **MRI Magnet: Tesla** | **MRI Magnet: Brand** | **Acquisition: Sequence** | **Acquisition: Duration** | **Voxel size, mm/slice thickness** | **Repetition time TR, ms** | **Echo time TE, ms** | **Flip angle °** | **Field of View FOV mm** | **Matrix size** | **Technique for segmentation (automated, semi automated or manual)** | **Programme used for analysis (SPM/freesurfer)** |
| --- | --- | --- | --- | --- | --- | --- | --- | --- | --- | --- | --- | --- |
| Duriez 2014 | 1.5 | Siemens | 3D SPGR | NS | 1.0x0.98x0.98 | 9.7 | 4 | NS | NS | 256x192x256 | Automated | SPM |
| Kim 2018 | 1.5 | GE signa | FSPGR | NS | 1.6mm slice thickness | 7.32 | 3.2 | 12 | 220x220 | 512x512 | Automated | BRAINS |
| Kim 2018 | 1.5 | GE signa | FSPGR | NS | 1.6mm slice thickness | 7.7 | 3.37 | 12 | 240x240 | 512x512 | Automated | BRAINS |
| Kim 2018 | 1.5 | GE signa | FSPGR | NS | 1.2mm slice thickness | 10.9 | 5.01 | 13 | 240x240 | 512x512 | Automated | BRAINS |
| Otsuka 2022 | 3 | Siemens | MPRAGE | NS | 0.98x0.98x1.1 | 1800 | 1.98 | 9 | NS | 256x256 | Automated | FreeSurfer |
| Van Haren 2010 | 1.5 | Phillips | 3D-FFE | NS | 1.2mm slice thickness | 30 | 4.6 | 30 | 256x256 | NS | Automated | NS |

### Table 12: Cannabis cross sectional risk of bias results

|  | **Exposure** | | | | **Comparability** | | | | **Outcome** | |  |
| --- | --- | --- | --- | --- | --- | --- | --- | --- | --- | --- | --- |
| **Study ID** | **1) Representativeness of the sample** | **2) Sample size** | **3) Non-respondents** | **4) Ascertainment of exposure** | **1) Did the study account for other drug use?** | **2) FOR CANNABIS/CO-USE ONLY: Study described route of administration?** | **3) Study matches groups or controls for Age** | **4) Study matches groups or controls for sex or gender** | **1) Assessment of outcome** | **2) Statistical test** | **%** |
| Ashtari 2011 | c) Selected group of users/convenience sample. | b) Not justified. | c) No information provided | b) *self-report using detailed descriptions (e.g. smoke every day for last month) or self-report from participant living in a place where cannabis/tobacco is legal | c) No or Unclear | a) *Yes | b) No or unclear | a) *Yes | c) Manual tracing unblinded | a) *Statistical test used to analyse the data clearly described, appropriate and measures of association presented, and unit of ROI measure reported | 33.3 |
| Batalla 2018 | c) Selected group of users/convenience sample. | b) Not justified. | c) No information provided | a) **secure record (Laboratory sample test, e.g. CO/saliva samples for tobacco, urine sample for THC) | a) **alcohol and tobacco/cannabis AND any other drug use | b) No or unclear | a) *Yes | a) *Yes | b) *Manual tracing blinded | a) *Statistical test used to analyse the data clearly described, appropriate and measures of association presented, and unit of ROI measure reported | 66.7 |
| Block 2000 | c) Selected group of users/convenience sample. | b) Not justified. | c) No information provided | c) self-report only | c) No or Unclear | b) No or unclear | a) *Yes | a) *Yes | c) Manual tracing unblinded | a) *Statistical test used to analyse the data clearly described, appropriate and measures of association presented, and unit of ROI measure reported | 25 |
| Buchy 2016 | b) *Somewhat representative of the average in the target group. (non-random sampling) | c) No information provided | c) No information provided | c) self-report only | c) No or Unclear | b) No or unclear | b) No or unclear | b) No or unclear | a) *Automated brain tissue volume computations (e.g. SPM, FreeSurfer) | a) *Statistical test used to analyse the data clearly described, appropriate and measures of association presented, and unit of ROI measure reported | 25 |
| Cahn 2004 | c) Selected group of users/convenience sample. | b) Not justified. | c) No information provided | b) *self-report using detailed descriptions (e.g. smoke every day for last month) or self-report from participant living in a place where cannabis/tobacco is legal | b) *alcohol and tobacco/cannabis | b) No or unclear | a) *Yes | a) *Yes | b) *Manual tracing blinded | a) *Statistical test used to analyse the data clearly described, appropriate and measures of association presented, and unit of ROI measure reported | 50 |
| Churchwell 2010 | d) No description of the derivation of the included subjects. | b) Not justified. | c) No information provided | a) **secure record (Laboratory sample test, e.g. CO/saliva samples for tobacco, urine sample for THC) | c) No or Unclear | b) No or unclear | b) No or unclear | b) No or unclear | c) Manual tracing unblinded | a) *Statistical test used to analyse the data clearly described, appropriate and measures of association presented, and unit of ROI measure reported | 25 |
| Churchwell 2012 | d) No description of the derivation of the included subjects. | b) Not justified. | c) No information provided | c) self-report only | b) *alcohol and tobacco/cannabis | b) No or unclear | a) *Yes | b) No or unclear | a) *Automated brain tissue volume computations (e.g. SPM, FreeSurfer) | a) *Statistical test used to analyse the data clearly described, appropriate and measures of association presented, and unit of ROI measure reported | 41.7 |
| Chye 2017b | b) *Somewhat representative of the average in the target group. (non-random sampling) | b) Not justified. | c) No information provided | a) **secure record (Laboratory sample test, e.g. CO/saliva samples for tobacco, urine sample for THC) | a) **alcohol and tobacco/cannabis AND any other drug use | b) No or unclear | a) *Yes | a) *Yes | a) *Automated brain tissue volume computations (e.g. SPM, FreeSurfer) | a) *Statistical test used to analyse the data clearly described, appropriate and measures of association presented, and unit of ROI measure reported | 75 |
| Cohen 2012 | d) No description of the derivation of the included subjects. | b) Not justified. | c) No information provided | c) self-report only | b) *alcohol and tobacco/cannabis | b) No or unclear | a) *Yes | a) *Yes | b) *Manual tracing blinded | a) *Statistical test used to analyse the data clearly described, appropriate and measures of association presented, and unit of ROI measure reported | 41.7 |
| D'Souza 2021 | b) *Somewhat representative of the average in the target group. (non-random sampling) | b) Not justified. | c) No information provided | a) **secure record (Laboratory sample test, e.g. CO/saliva samples for tobacco, urine sample for THC) | a) **alcohol and tobacco/cannabis AND any other drug use | a) *Yes | a) *Yes | a) *Yes | a) *Automated brain tissue volume computations (e.g. SPM, FreeSurfer) | a) *Statistical test used to analyse the data clearly described, appropriate and measures of association presented, and unit of ROI measure reported | 83.3 |
| Gilman 2014 | c) Selected group of users/convenience sample. | b) Not justified. | c) No information provided | a) **secure record (Laboratory sample test, e.g. CO/saliva samples for tobacco, urine sample for THC) | c) No or Unclear | a) *Yes | a) *Yes | a) *Yes | a) *Automated brain tissue volume computations (e.g. SPM, FreeSurfer) | a) *Statistical test used to analyse the data clearly described, appropriate and measures of association presented, and unit of ROI measure reported | 58.3 |
| James 2011 | c) Selected group of users/convenience sample. | b) Not justified. | c) No information provided | a) **secure record (Laboratory sample test, e.g. CO/saliva samples for tobacco, urine sample for THC) | a) **alcohol and tobacco/cannabis AND any other drug use | b) No or unclear | a) *Yes | a) *Yes | a) *Automated brain tissue volume computations (e.g. SPM, FreeSurfer) | a) *Statistical test used to analyse the data clearly described, appropriate and measures of association presented, and unit of ROI measure reported | 66.7 |
| Knodt 2022 | a) *Truly representative of the average in the target population. (all subjects or random sampling) | a) *Justified and satisfactory (including sample size calculation). | b) Unsatisfactory recruitment rate, no summary data on non-respondents. | b) *self-report using detailed descriptions (e.g. smoke every day for last month) or self-report from participant living in a place where cannabis/tobacco is legal | c) No or Unclear | b) No or unclear | a) *Yes | a) *Yes | a) *Automated brain tissue volume computations (e.g. SPM, FreeSurfer) | a) *Statistical test used to analyse the data clearly described, appropriate and measures of association presented, and unit of ROI measure reported | 58.3 |
| Koenders 2015 | c) Selected group of users/convenience sample. | b) Not justified. | c) No information provided | b) *self-report using detailed descriptions (e.g. smoke every day for last month) or self-report from participant living in a place where cannabis/tobacco is legal | c) No or Unclear | b) No or unclear | a) *Yes | a) *Yes | a) *Automated brain tissue volume computations (e.g. SPM, FreeSurfer) | a) *Statistical test used to analyse the data clearly described, appropriate and measures of association presented, and unit of ROI measure reported | 41.7 |
| Koenis 2021 | b) *Somewhat representative of the average in the target group. (non-random sampling) | b) Not justified. | b) Unsatisfactory recruitment rate, no summary data on non-respondents. | a) **secure record (Laboratory sample test, e.g. CO/saliva samples for tobacco, urine sample for THC) | c) No or Unclear | b) No or unclear | a) *Yes | a) *Yes | a) *Automated brain tissue volume computations (e.g. SPM, FreeSurfer) | a) *Statistical test used to analyse the data clearly described, appropriate and measures of association presented, and unit of ROI measure reported | 58.3 |
| Kumra 2012 | c) Selected group of users/convenience sample. | b) Not justified. | b) Unsatisfactory recruitment rate, no summary data on non-respondents. | a) **secure record (Laboratory sample test, e.g. CO/saliva samples for tobacco, urine sample for THC) | c) No or Unclear | b) No or unclear | a) *Yes | a) *Yes | b) *Manual tracing blinded | a) *Statistical test used to analyse the data clearly described, appropriate and measures of association presented, and unit of ROI measure reported | 50 |
| Levar 2018 | d) No description of the derivation of the included subjects. | b) Not justified. | c) No information provided | a) **secure record (Laboratory sample test, e.g. CO/saliva samples for tobacco, urine sample for THC) | a) **alcohol and tobacco/cannabis AND any other drug use | b) No or unclear | a) *Yes | a) *Yes | a) *Automated brain tissue volume computations (e.g. SPM, FreeSurfer) | b) Statistical test not appropriate, not described or incomplete. | 58.3 |
| Lisdahl 2016 | b) *Somewhat representative of the average in the target group. (non-random sampling) | a) *Justified and satisfactory (including sample size calculation). | c) No information provided | b) *self-report using detailed descriptions (e.g. smoke every day for last month) or self-report from participant living in a place where cannabis/tobacco is legal | a) **alcohol and tobacco/cannabis AND any other drug use | b) No or unclear | a) *Yes | a) *Yes | a) *Automated brain tissue volume computations (e.g. SPM, FreeSurfer) | a) *Statistical test used to analyse the data clearly described, appropriate and measures of association presented, and unit of ROI measure reported | 75 |
| Lopez-Larson 2011 | c) Selected group of users/convenience sample. | b) Not justified. | c) No information provided | c) self-report only | c) No or Unclear | b) No or unclear | a) *Yes | b) No or unclear | a) *Automated brain tissue volume computations (e.g. SPM, FreeSurfer) | a) *Statistical test used to analyse the data clearly described, appropriate and measures of association presented, and unit of ROI measure reported | 25 |
| Lorenzetti 2020 | d) No description of the derivation of the included subjects. | b) Not justified. | c) No information provided | b) *self-report using detailed descriptions (e.g. smoke every day for last month) or self-report from participant living in a place where cannabis/tobacco is legal | c) No or Unclear | a) *Yes | a) *Yes | a) *Yes | b) *Manual tracing blinded | a) *Statistical test used to analyse the data clearly described, appropriate and measures of association presented, and unit of ROI measure reported | 50 |
| Lorenzetti 2024 | b) *Somewhat representative of the average in the target group. (non-random sampling) | a) *Justified and satisfactory (including sample size calculation). | c) No information provided | b) *self-report using detailed descriptions (e.g. smoke every day for last month) or self-report from participant living in a place where cannabis/tobacco is legal | a) **alcohol and tobacco/cannabis AND any other drug use | b) No or unclear | a) *Yes | a) *Yes | a) *Automated brain tissue volume computations (e.g. SPM, FreeSurfer) | a) *Statistical test used to analyse the data clearly described, appropriate and measures of association presented, and unit of ROI measure reported | 75 |
| Maple 2019 | c) Selected group of users/convenience sample. | b) Not justified. | c) No information provided | b) *self-report using detailed descriptions (e.g. smoke every day for last month) or self-report from participant living in a place where cannabis/tobacco is legal | a) **alcohol and tobacco/cannabis AND any other drug use | a) *Yes | b) No or unclear | a) *Yes | a) *Automated brain tissue volume computations (e.g. SPM, FreeSurfer) | a) *Statistical test used to analyse the data clearly described, appropriate and measures of association presented, and unit of ROI measure reported | 58.3 |
| Mashhoon 2015 | b) *Somewhat representative of the average in the target group. (non-random sampling) | b) Not justified. | c) No information provided | a) **secure record (Laboratory sample test, e.g. CO/saliva samples for tobacco, urine sample for THC) | a) **alcohol and tobacco/cannabis AND any other drug use | b) No or unclear | a) *Yes | a) *Yes | a) *Automated brain tissue volume computations (e.g. SPM, FreeSurfer) | b) Statistical test not appropriate, not described or incomplete. | 66.7 |
| Mata 2010 | c) Selected group of users/convenience sample. | b) Not justified. | c) No information provided | c) self-report only | c) No or Unclear | b) No or unclear | a) *Yes | a) *Yes | a) *Automated brain tissue volume computations (e.g. SPM, FreeSurfer) | a) *Statistical test used to analyse the data clearly described, appropriate and measures of association presented, and unit of ROI measure reported | 33.3 |
| McQueeny 2011 | c) Selected group of users/convenience sample. | b) Not justified. | c) No information provided | b) *self-report using detailed descriptions (e.g. smoke every day for last month) or self-report from participant living in a place where cannabis/tobacco is legal | c) No or Unclear | b) No or unclear | a) *Yes | a) *Yes | b) *Manual tracing blinded | a) *Statistical test used to analyse the data clearly described, appropriate and measures of association presented, and unit of ROI measure reported | 41.7 |
| Medina 2007a | b) *Somewhat representative of the average in the target group. (non-random sampling) | b) Not justified. | c) No information provided | b) *self-report using detailed descriptions (e.g. smoke every day for last month) or self-report from participant living in a place where cannabis/tobacco is legal | a) **alcohol and tobacco/cannabis AND any other drug use | b) No or unclear | a) *Yes | a) *Yes | b) *Manual tracing blinded | a) *Statistical test used to analyse the data clearly described, appropriate and measures of association presented, and unit of ROI measure reported | 66.7 |
| Medina 2007b | b) *Somewhat representative of the average in the target group. (non-random sampling) | b) Not justified. | c) No information provided | b) *self-report using detailed descriptions (e.g. smoke every day for last month) or self-report from participant living in a place where cannabis/tobacco is legal | c) No or Unclear | b) No or unclear | a) *Yes | a) *Yes | b) *Manual tracing blinded | a) *Statistical test used to analyse the data clearly described, appropriate and measures of association presented, and unit of ROI measure reported | 50 |
| Medina 2009 | b) *Somewhat representative of the average in the target group. (non-random sampling) | b) Not justified. | c) No information provided | b) *self-report using detailed descriptions (e.g. smoke every day for last month) or self-report from participant living in a place where cannabis/tobacco is legal | a) **alcohol and tobacco/cannabis AND any other drug use | b) No or unclear | b) No or unclear | a) *Yes | b) *Manual tracing blinded | a) *Statistical test used to analyse the data clearly described, appropriate and measures of association presented, and unit of ROI measure reported | 50 |
| Medina 2010 | b) *Somewhat representative of the average in the target group. (non-random sampling) | b) Not justified. | c) No information provided | b) *self-report using detailed descriptions (e.g. smoke every day for last month) or self-report from participant living in a place where cannabis/tobacco is legal | c) No or Unclear | b) No or unclear | a) *Yes | a) *Yes | b) *Manual tracing blinded | a) *Statistical test used to analyse the data clearly described, appropriate and measures of association presented, and unit of ROI measure reported | 50 |
| Meier 2019 | a) *Truly representative of the average in the target population. (all subjects or random sampling) | a) *Justified and satisfactory (including sample size calculation). | a) *Proportion of target sample recruited attains pre-specified target or basic summary of non-respondent characteristics in sampling frame recorded. | b) *self-report using detailed descriptions (e.g. smoke every day for last month) or self-report from participant living in a place where cannabis/tobacco is legal | b) *alcohol and tobacco/cannabis | b) No or unclear | a) *Yes | a) *Yes | a) *Automated brain tissue volume computations (e.g. SPM, FreeSurfer) | a) *Statistical test used to analyse the data clearly described, appropriate and measures of association presented, and unit of ROI measure reported | 75 |
| Meier 2022 | a) *Truly representative of the average in the target population. (all subjects or random sampling) | a) *Justified and satisfactory (including sample size calculation). | a) *Proportion of target sample recruited attains pre-specified target or basic summary of non-respondent characteristics in sampling frame recorded. | b) *self-report using detailed descriptions (e.g. smoke every day for last month) or self-report from participant living in a place where cannabis/tobacco is legal | c) No or Unclear | b) No or unclear | a) *Yes | a) *Yes | a) *Automated brain tissue volume computations (e.g. SPM, FreeSurfer) | a) *Statistical test used to analyse the data clearly described, appropriate and measures of association presented, and unit of ROI measure reported | 66.7 |
| Moreno-Alcazar 2018 | b) *Somewhat representative of the average in the target group. (non-random sampling) | b) Not justified. | c) No information provided | b) *self-report using detailed descriptions (e.g. smoke every day for last month) or self-report from participant living in a place where cannabis/tobacco is legal | c) No or Unclear | a) *Yes | a) *Yes | a) *Yes | a) *Automated brain tissue volume computations (e.g. SPM, FreeSurfer) | a) *Statistical test used to analyse the data clearly described, appropriate and measures of association presented, and unit of ROI measure reported | 58.3 |
| Navarri 2022 | a) *Truly representative of the average in the target population. (all subjects or random sampling) | a) *Justified and satisfactory (including sample size calculation). | c) No information provided | b) *self-report using detailed descriptions (e.g. smoke every day for last month) or self-report from participant living in a place where cannabis/tobacco is legal | c) No or Unclear | b) No or unclear | a) *Yes | a) *Yes | a) *Automated brain tissue volume computations (e.g. SPM, FreeSurfer) | a) *Statistical test used to analyse the data clearly described, appropriate and measures of association presented, and unit of ROI measure reported | 58.3 |
| Owens 2021 | b) *Somewhat representative of the average in the target group. (non-random sampling) | b) Not justified. | b) Unsatisfactory recruitment rate, no summary data on non-respondents. | a) **secure record (Laboratory sample test, e.g. CO/saliva samples for tobacco, urine sample for THC) | a) **alcohol and tobacco/cannabis AND any other drug use | b) No or unclear | a) *Yes | a) *Yes | a) *Automated brain tissue volume computations (e.g. SPM, FreeSurfer) | a) *Statistical test used to analyse the data clearly described, appropriate and measures of association presented, and unit of ROI measure reported | 75 |
| Price 2015 | c) Selected group of users/convenience sample. | b) Not justified. | c) No information provided | b) *self-report using detailed descriptions (e.g. smoke every day for last month) or self-report from participant living in a place where cannabis/tobacco is legal | a) **alcohol and tobacco/cannabis AND any other drug use | a) *Yes | a) *Yes | a) *Yes | a) *Automated brain tissue volume computations (e.g. SPM, FreeSurfer) | a) *Statistical test used to analyse the data clearly described, appropriate and measures of association presented, and unit of ROI measure reported | 66.7 |
| Radoman 2019 | b) *Somewhat representative of the average in the target group. (non-random sampling) | b) Not justified. | c) No information provided | a) **secure record (Laboratory sample test, e.g. CO/saliva samples for tobacco, urine sample for THC) | c) No or Unclear | b) No or unclear | a) *Yes | a) *Yes | a) *Automated brain tissue volume computations (e.g. SPM, FreeSurfer) | a) *Statistical test used to analyse the data clearly described, appropriate and measures of association presented, and unit of ROI measure reported | 58.3 |
| Rapp 2013 | c) Selected group of users/convenience sample. | b) Not justified. | c) No information provided | a) **secure record (Laboratory sample test, e.g. CO/saliva samples for tobacco, urine sample for THC) | b) *alcohol and tobacco/cannabis | b) No or unclear | a) *Yes | a) *Yes | b) *Manual tracing blinded | a) *Statistical test used to analyse the data clearly described, appropriate and measures of association presented, and unit of ROI measure reported | 58.3 |
| Romero 2015 | c) Selected group of users/convenience sample. | b) Not justified. | c) No information provided | a) **secure record (Laboratory sample test, e.g. CO/saliva samples for tobacco, urine sample for THC) | c) No or Unclear | b) No or unclear | a) *Yes | a) *Yes | a) *Automated brain tissue volume computations (e.g. SPM, FreeSurfer) | b) Statistical test not appropriate, not described or incomplete. | 41.7 |
| Rosetti 2021 | a) *Truly representative of the average in the target population. (all subjects or random sampling) | a) *Justified and satisfactory (including sample size calculation). | c) No information provided | b) *self-report using detailed descriptions (e.g. smoke every day for last month) or self-report from participant living in a place where cannabis/tobacco is legal | a) **alcohol and tobacco/cannabis AND any other drug use | b) No or unclear | a) *Yes | a) *Yes | a) *Automated brain tissue volume computations (e.g. SPM, FreeSurfer) | a) *Statistical test used to analyse the data clearly described, appropriate and measures of association presented, and unit of ROI measure reported | 75 |
| Schacht 2012 | c) Selected group of users/convenience sample. | b) Not justified. | c) No information provided | a) **secure record (Laboratory sample test, e.g. CO/saliva samples for tobacco, urine sample for THC) | b) *alcohol and tobacco/cannabis | b) No or unclear | a) *Yes | a) *Yes | a) *Automated brain tissue volume computations (e.g. SPM, FreeSurfer) | a) *Statistical test used to analyse the data clearly described, appropriate and measures of association presented, and unit of ROI measure reported | 58.3 |
| Scheffler 2021 | c) Selected group of users/convenience sample. | b) Not justified. | b) Unsatisfactory recruitment rate, no summary data on non-respondents. | a) **secure record (Laboratory sample test, e.g. CO/saliva samples for tobacco, urine sample for THC) | a) **alcohol and tobacco/cannabis AND any other drug use | b) No or unclear | b) No or unclear | b) No or unclear | a) *Automated brain tissue volume computations (e.g. SPM, FreeSurfer) | b) Statistical test not appropriate, not described or incomplete. | 41.7 |
| Scott 2019 | a) *Truly representative of the average in the target population. (all subjects or random sampling) | b) Not justified. | c) No information provided | c) self-report only | c) No or Unclear | b) No or unclear | a) *Yes | a) *Yes | a) *Automated brain tissue volume computations (e.g. SPM, FreeSurfer) | b) Statistical test not appropriate, not described or incomplete. | 33.3 |
| Szeszko 2007 | c) Selected group of users/convenience sample. | b) Not justified. | c) No information provided | b) *self-report using detailed descriptions (e.g. smoke every day for last month) or self-report from participant living in a place where cannabis/tobacco is legal | c) No or Unclear | b) No or unclear | a) *Yes | b) No or unclear | b) *Manual tracing blinded | a) *Statistical test used to analyse the data clearly described, appropriate and measures of association presented, and unit of ROI measure reported | 33.3 |
| Thayer 2019a | b) *Somewhat representative of the average in the target group. (non-random sampling) | b) Not justified. | c) No information provided | b) *self-report using detailed descriptions (e.g. smoke every day for last month) or self-report from participant living in a place where cannabis/tobacco is legal | a) **alcohol and tobacco/cannabis AND any other drug use | b) No or unclear | a) *Yes | b) No or unclear | a) *Automated brain tissue volume computations (e.g. SPM, FreeSurfer) | b) Statistical test not appropriate, not described or incomplete. | 50 |
| Thayer 2019b | b) *Somewhat representative of the average in the target group. (non-random sampling) | a) *Justified and satisfactory (including sample size calculation). | a) *Proportion of target sample recruited attains pre-specified target or basic summary of non-respondent characteristics in sampling frame recorded. | b) *self-report using detailed descriptions (e.g. smoke every day for last month) or self-report from participant living in a place where cannabis/tobacco is legal | a) **alcohol and tobacco/cannabis AND any other drug use | a) *Yes | b) No or unclear | b) No or unclear | a) *Automated brain tissue volume computations (e.g. SPM, FreeSurfer) | a) *Statistical test used to analyse the data clearly described, appropriate and measures of association presented, and unit of ROI measure reported | 75 |
| Tzilos 2005 | c) Selected group of users/convenience sample. | b) Not justified. | c) No information provided | c) self-report only | c) No or Unclear | b) No or unclear | a) *Yes | a) *Yes | a) *Automated brain tissue volume computations (e.g. SPM, FreeSurfer) | a) *Statistical test used to analyse the data clearly described, appropriate and measures of association presented, and unit of ROI measure reported | 33.3 |
| Vered 2024 | b) *Somewhat representative of the average in the target group. (non-random sampling) | a) *Justified and satisfactory (including sample size calculation). | a) *Proportion of target sample recruited attains pre-specified target or basic summary of non-respondent characteristics in sampling frame recorded. | c) self-report only | b) *alcohol and tobacco/cannabis | b) No or unclear | a) *Yes | a) *Yes | d) no description | a) *Statistical test used to analyse the data clearly described, appropriate and measures of association presented, and unit of ROI measure reported | 58.3 |
| Wallace 2024 | b) *Somewhat representative of the average in the target group. (non-random sampling) | a) *Justified and satisfactory (including sample size calculation). | c) No information provided | a) **secure record (Laboratory sample test, e.g. CO/saliva samples for tobacco, urine sample for THC) | c) No or Unclear | b) No or unclear | a) *Yes | a) *Yes | a) *Automated brain tissue volume computations (e.g. SPM, FreeSurfer) | a) *Statistical test used to analyse the data clearly described, appropriate and measures of association presented, and unit of ROI measure reported | 66.7 |
| Weiland 2015 | b) *Somewhat representative of the average in the target group. (non-random sampling) | b) Not justified. | c) No information provided | b) *self-report using detailed descriptions (e.g. smoke every day for last month) or self-report from participant living in a place where cannabis/tobacco is legal | b) *alcohol and tobacco/cannabis | b) No or unclear | a) *Yes | a) *Yes | a) *Automated brain tissue volume computations (e.g. SPM, FreeSurfer) | a) *Statistical test used to analyse the data clearly described, appropriate and measures of association presented, and unit of ROI measure reported | 58.3 |
| Yip 2014 | b) *Somewhat representative of the average in the target group. (non-random sampling) | b) Not justified. | b) Unsatisfactory recruitment rate, no summary data on non-respondents. | c) self-report only | c) No or Unclear | b) No or unclear | a) *Yes | a) *Yes | a) *Automated brain tissue volume computations (e.g. SPM, FreeSurfer) | a) *Statistical test used to analyse the data clearly described, appropriate and measures of association presented, and unit of ROI measure reported | 33.3 |

### Table 13: Tobacco cross-sectional risk of bias results

|  | **Exposure** | | | | **Comparability** | | | **Outcome** | |  |
| --- | --- | --- | --- | --- | --- | --- | --- | --- | --- | --- |
| **Study ID** | **1) Representativeness of the sample** | **2) Sample size** | **3) Non-respondents** | **4) Ascertainment of exposure** | **1) Did the study account for other drug use?** | **2) Study matches groups or controls for Age** | **3) Study matches groups or controls for sex or gender** | **1) Assessment of outcome** | **2) Statistical test** | **%** |
| Austin 2022 | b) *Somewhat representative of the average in the target group. (non-random sampling) | a) *Justified and satisfactory (including sample size calculation). | a) *Proportion of target sample recruited attains pre-specified target or basic summary of non-respondent characteristics in sampling frame recorded. | c) self-report only | c) No or Unclear | a) *Yes | a) *Yes | a) *Automated brain tissue volume computations (e.g. SPM, FreeSurfer) | a) *Statistical test used to analyse the data clearly described, appropriate and measures of association presented, and unit of ROI measure reported | 63.6 |
| Binnewies 2023 | b) *Somewhat representative of the average in the target group. (non-random sampling) | a) *Justified and satisfactory (including sample size calculation). | c) No information provided | c) self-report only | c) No or Unclear | a) *Yes | a) *Yes | a) *Automated brain tissue volume computations (e.g. SPM, FreeSurfer) | a) *Statistical test used to analyse the data clearly described, appropriate and measures of association presented, and unit of ROI measure reported | 54.5 |
| Brody 2004 | b) *Somewhat representative of the average in the target group. (non-random sampling) | b) Not justified. | c) No information provided | b) *self-report using detailed descriptions (e.g. smoke every day for last month) or self-report from participant living in a place where cannabis/tobacco is legal | a) **alcohol and tobacco/cannabis AND any other drug use | a) *Yes | a) *Yes | b) *Manual tracing blinded | a) *Statistical test used to analyse the data clearly described, appropriate and measures of association presented, and unit of ROI measure reported | 72.7 |
| Cardenas 2020 | c) Selected group of users/convenience sample. | b) Not justified. | b) Unsatisfactory recruitment rate, no summary data on non-respondents. | b) *self-report using detailed descriptions (e.g. smoke every day for last month) or self-report from participant living in a place where cannabis/tobacco is legal | a) **alcohol and tobacco/cannabis AND any other drug use | a) *Yes | a) *Yes | a) *Automated brain tissue volume computations (e.g. SPM, FreeSurfer) | a) *Statistical test used to analyse the data clearly described, appropriate and measures of association presented, and unit of ROI measure reported | 63.6 |
| Chen 2006 | c) Selected group of users/convenience sample. | b) Not justified. | c) No information provided | c) self-report only | c) No or Unclear | a) *Yes | b) No or unclear | d) no description | a) *Statistical test used to analyse the data clearly described, appropriate and measures of association presented, and unit of ROI measure reported | 18.2 |
| Chen 2023 | b) *Somewhat representative of the average in the target group. (non-random sampling) | a) *Justified and satisfactory (including sample size calculation). | c) No information provided | c) self-report only | c) No or Unclear | a) *Yes | a) *Yes | a) *Automated brain tissue volume computations (e.g. SPM, FreeSurfer) | a) *Statistical test used to analyse the data clearly described, appropriate and measures of association presented, and unit of ROI measure reported | 54.5 |
| Cho 2016 | c) Selected group of users/convenience sample. | c) No information provided | c) No information provided | b) *self-report using detailed descriptions (e.g. smoke every day for last month) or self-report from participant living in a place where cannabis/tobacco is legal | c) No or Unclear | b) No or unclear | b) No or unclear | a) *Automated brain tissue volume computations (e.g. SPM, FreeSurfer) | a) *Statistical test used to analyse the data clearly described, appropriate and measures of association presented, and unit of ROI measure reported | 27.3 |
| Choi 2010 | c) Selected group of users/convenience sample. | b) Not justified. | c) No information provided | c) self-report only | c) No or Unclear | a) *Yes | a) *Yes | a) *Automated brain tissue volume computations (e.g. SPM, FreeSurfer) | a) *Statistical test used to analyse the data clearly described, appropriate and measures of association presented, and unit of ROI measure reported | 36.4 |
| Chye 2020 | b) *Somewhat representative of the average in the target group. (non-random sampling) | a) *Justified and satisfactory (including sample size calculation). | c) No information provided | b) *self-report using detailed descriptions (e.g. smoke every day for last month) or self-report from participant living in a place where cannabis/tobacco is legal | a) **alcohol and tobacco/cannabis AND any other drug use | a) *Yes | a) *Yes | a) *Automated brain tissue volume computations (e.g. SPM, FreeSurfer) | a) *Statistical test used to analyse the data clearly described, appropriate and measures of association presented, and unit of ROI measure reported | 81.8 |
| Durazzo 2007a | b) *Somewhat representative of the average in the target group. (non-random sampling) | b) Not justified. | a) *Proportion of target sample recruited attains pre-specified target or basic summary of non-respondent characteristics in sampling frame recorded. | c) self-report only | c) No or Unclear | a) *Yes | b) No or unclear | a) *Automated brain tissue volume computations (e.g. SPM, FreeSurfer) | b) Statistical test not appropriate, not described or incomplete. | 36.4 |
| Durazzo 2013a | b) *Somewhat representative of the average in the target group. (non-random sampling) | b) Not justified. | c) No information provided | b) *self-report using detailed descriptions (e.g. smoke every day for last month) or self-report from participant living in a place where cannabis/tobacco is legal | a) **alcohol and tobacco/cannabis AND any other drug use | a) *Yes | a) *Yes | a) *Automated brain tissue volume computations (e.g. SPM, FreeSurfer) | a) *Statistical test used to analyse the data clearly described, appropriate and measures of association presented, and unit of ROI measure reported | 72.7 |
| Durazzo 2013b | c) Selected group of users/convenience sample. | b) Not justified. | c) No information provided | c) self-report only | a) **alcohol and tobacco/cannabis AND any other drug use | a) *Yes | b) No or unclear | a) *Automated brain tissue volume computations (e.g. SPM, FreeSurfer) | a) *Statistical test used to analyse the data clearly described, appropriate and measures of association presented, and unit of ROI measure reported | 45.5 |
| Durazzo 2017 | b) *Somewhat representative of the average in the target group. (non-random sampling) | b) Not justified. | b) Unsatisfactory recruitment rate, no summary data on non-respondents. | b) *self-report using detailed descriptions (e.g. smoke every day for last month) or self-report from participant living in a place where cannabis/tobacco is legal | a) **alcohol and tobacco/cannabis AND any other drug use | a) *Yes | a) *Yes | a) *Automated brain tissue volume computations (e.g. SPM, FreeSurfer) | a) *Statistical test used to analyse the data clearly described, appropriate and measures of association presented, and unit of ROI measure reported | 72.7 |
| Durhan 2016 | c) Selected group of users/convenience sample. | b) Not justified. | b) Unsatisfactory recruitment rate, no summary data on non-respondents. | c) self-report only | c) No or Unclear | a) *Yes | b) No or unclear | a) *Automated brain tissue volume computations (e.g. SPM, FreeSurfer) | b) Statistical test not appropriate, not described or incomplete. | 18.2 |
| Duriez 2014 cross-sectional | b) *Somewhat representative of the average in the target group. (non-random sampling) | b) Not justified. | a) *Proportion of target sample recruited attains pre-specified target or basic summary of non-respondent characteristics in sampling frame recorded. | c) self-report only | c) No or Unclear | a) *Yes | a) *Yes | a) *Automated brain tissue volume computations (e.g. SPM, FreeSurfer) | a) *Statistical test used to analyse the data clearly described, appropriate and measures of association presented, and unit of ROI measure reported | 54.5 |
| Elbejjani 2019 | b) *Somewhat representative of the average in the target group. (non-random sampling) | b) Not justified. | a) *Proportion of target sample recruited attains pre-specified target or basic summary of non-respondent characteristics in sampling frame recorded. | b) *self-report using detailed descriptions (e.g. smoke every day for last month) or self-report from participant living in a place where cannabis/tobacco is legal | a) **alcohol and tobacco/cannabis AND any other drug use | a) *Yes | a) *Yes | a) *Automated brain tissue volume computations (e.g. SPM, FreeSurfer) | a) *Statistical test used to analyse the data clearly described, appropriate and measures of association presented, and unit of ROI measure reported | 81.8 |
| Gazdzinksi 2005 | c) Selected group of users/convenience sample. | b) Not justified. | c) No information provided | b) *self-report using detailed descriptions (e.g. smoke every day for last month) or self-report from participant living in a place where cannabis/tobacco is legal | a) **alcohol and tobacco/cannabis AND any other drug use | a) *Yes | a) *Yes | a) *Automated brain tissue volume computations (e.g. SPM, FreeSurfer) | a) *Statistical test used to analyse the data clearly described, appropriate and measures of association presented, and unit of ROI measure reported | 63.6 |
| Hoogendam 2012 | b) *Somewhat representative of the average in the target group. (non-random sampling) | b) Not justified. | a) *Proportion of target sample recruited attains pre-specified target or basic summary of non-respondent characteristics in sampling frame recorded. | c) self-report only | c) No or Unclear | a) *Yes | a) *Yes | a) *Automated brain tissue volume computations (e.g. SPM, FreeSurfer) | a) *Statistical test used to analyse the data clearly described, appropriate and measures of association presented, and unit of ROI measure reported | 54.5 |
| Janowitz 2014 | a) *Truly representative of the average in the target population. (all subjects or random sampling) | a) *Justified and satisfactory (including sample size calculation). | b) Unsatisfactory recruitment rate, no summary data on non-respondents. | b) *self-report using detailed descriptions (e.g. smoke every day for last month) or self-report from participant living in a place where cannabis/tobacco is legal | c) No or Unclear | a) *Yes | a) *Yes | a) *Automated brain tissue volume computations (e.g. SPM, FreeSurfer) | a) *Statistical test used to analyse the data clearly described, appropriate and measures of association presented, and unit of ROI measure reported | 63.6 |
| Jha 2021 | b) *Somewhat representative of the average in the target group. (non-random sampling) | a) *Justified and satisfactory (including sample size calculation). | a) *Proportion of target sample recruited attains pre-specified target or basic summary of non-respondent characteristics in sampling frame recorded. | c) self-report only | c) No or Unclear | a) *Yes | a) *Yes | a) *Automated brain tissue volume computations (e.g. SPM, FreeSurfer) | a) *Statistical test used to analyse the data clearly described, appropriate and measures of association presented, and unit of ROI measure reported | 63.6 |
| Launer 2015 | b) *Somewhat representative of the average in the target group. (non-random sampling) | b) Not justified. | c) No information provided | b) *self-report using detailed descriptions (e.g. smoke every day for last month) or self-report from participant living in a place where cannabis/tobacco is legal | c) No or Unclear | a) *Yes | a) *Yes | a) *Automated brain tissue volume computations (e.g. SPM, FreeSurfer) | b) Statistical test not appropriate, not described or incomplete. | 45.5 |
| Li 2015 | c) Selected group of users/convenience sample. | b) Not justified. | c) No information provided | a) **secure record (Laboratory sample test, .eg CO/saliva samples for tobacco, urine sample for thc) | a) **alcohol and tobacco/cannabis AND any other drug use | a) *Yes | a) *Yes | a) *Automated brain tissue volume computations (e.g. SPM, FreeSurfer) | a) *Statistical test used to analyse the data clearly described, appropriate and measures of association presented, and unit of ROI measure reported | 72.7 |
| Liang 2022 | b) *Somewhat representative of the average in the target group. (non-random sampling) | a) *Justified and satisfactory (including sample size calculation). | b) Unsatisfactory recruitment rate, no summary data on non-respondents. | b) *self-report using detailed descriptions (e.g. smoke every day for last month) or self-report from participant living in a place where cannabis/tobacco is legal | c) No or Unclear | a) *Yes | a) *Yes | a) *Automated brain tissue volume computations (e.g. SPM, FreeSurfer) | a) *Statistical test used to analyse the data clearly described, appropriate and measures of association presented, and unit of ROI measure reported | 63.6 |
| Lie 2022 | b) *Somewhat representative of the average in the target group. (non-random sampling) | b) Not justified. | c) No information provided | a) **secure record (Laboratory sample test, .eg CO/saliva samples for tobacco, urine sample for thc) | c) No or Unclear | a) *Yes | a) *Yes | a) *Automated brain tissue volume computations (e.g. SPM, FreeSurfer) | a) *Statistical test used to analyse the data clearly described, appropriate and measures of association presented, and unit of ROI measure reported | 63.6 |
| Lin 2019 | d) No description of the derivation of the included subjects. | b) Not justified. | c) No information provided | b) *self-report using detailed descriptions (e.g. smoke every day for last month) or self-report from participant living in a place where cannabis/tobacco is legal | a) **alcohol and tobacco/cannabis AND any other drug use | a) *Yes | a) *Yes | a) *Automated brain tissue volume computations (e.g. SPM, FreeSurfer) | b) Statistical test not appropriate, not described or incomplete. | 54.5 |
| Lin 2021 | b) *Somewhat representative of the average in the target group. (non-random sampling) | b) Not justified. | c) No information provided | b) *self-report using detailed descriptions (e.g. smoke every day for last month) or self-report from participant living in a place where cannabis/tobacco is legal | c) No or Unclear | a) *Yes | a) *Yes | a) *Automated brain tissue volume computations (e.g. SPM, FreeSurfer) | a) *Statistical test used to analyse the data clearly described, appropriate and measures of association presented, and unit of ROI measure reported | 54.5 |
| Linli 2023 | b) *Somewhat representative of the average in the target group. (non-random sampling) | a) *Justified and satisfactory (including sample size calculation). | c) No information provided | b) *self-report using detailed descriptions (e.g. smoke every day for last month) or self-report from participant living in a place where cannabis/tobacco is legal | c) No or Unclear | a) *Yes | a) *Yes | a) *Automated brain tissue volume computations (e.g. SPM, FreeSurfer) | a) *Statistical test used to analyse the data clearly described, appropriate and measures of association presented, and unit of ROI measure reported | 63.6 |
| Luhar 2013 | c) Selected group of users/convenience sample. | b) Not justified. | c) No information provided | c) self-report only | a) **alcohol and tobacco/cannabis AND any other drug use | a) *Yes | a) *Yes | a) *Automated brain tissue volume computations (e.g. SPM, FreeSurfer) | a) *Statistical test used to analyse the data clearly described, appropriate and measures of association presented, and unit of ROI measure reported | 54.5 |
| Paul 2008 | b) *Somewhat representative of the average in the target group. (non-random sampling) | b) Not justified. | c) No information provided | c) self-report only | a) **alcohol and tobacco/cannabis AND any other drug use | a) *Yes | a) *Yes | a) *Automated brain tissue volume computations (e.g. SPM, FreeSurfer) | a) *Statistical test used to analyse the data clearly described, appropriate and measures of association presented, and unit of ROI measure reported | 63.6 |
| Pennington 2015 | c) Selected group of users/convenience sample. | b) Not justified. | c) No information provided | b) *self-report using detailed descriptions (e.g. smoke every day for last month) or self-report from participant living in a place where cannabis/tobacco is legal | a) **alcohol and tobacco/cannabis AND any other drug use | a) *Yes | a) *Yes | a) *Automated brain tissue volume computations (e.g. SPM, FreeSurfer) | b) Statistical test not appropriate, not described or incomplete. | 54.5 |
| Ringin 2022 | b) *Somewhat representative of the average in the target group. (non-random sampling) | a) *Justified and satisfactory (including sample size calculation). | c) No information provided | c) self-report only | c) No or Unclear | a) *Yes | a) *Yes | a) *Automated brain tissue volume computations (e.g. SPM, FreeSurfer) | a) *Statistical test used to analyse the data clearly described, appropriate and measures of association presented, and unit of ROI measure reported | 54.5 |
| Shang 2024 | b) *Somewhat representative of the average in the target group. (non-random sampling) | a) *Justified and satisfactory (including sample size calculation). | c) No information provided | c) self-report only | c) No or Unclear | a) *Yes | a) *Yes | a) *Automated brain tissue volume computations (e.g. SPM, FreeSurfer) | a) *Statistical test used to analyse the data clearly described, appropriate and measures of association presented, and unit of ROI measure reported | 54.5 |
| Shen 2017 | c) Selected group of users/convenience sample. | c) No information provided | c) No information provided | a) **secure record (Laboratory sample test, .eg CO/saliva samples for tobacco, urine sample for thc) | c) No or Unclear | b) No or unclear | b) No or unclear | a) *Automated brain tissue volume computations (e.g. SPM, FreeSurfer) | b) Statistical test not appropriate, not described or incomplete. | 27.3 |
| Valsdóttir 2022 | a) *Truly representative of the average in the target population. (all subjects or random sampling) | a) *Justified and satisfactory (including sample size calculation). | b) Unsatisfactory recruitment rate, no summary data on non-respondents. | c) self-report only | c) No or Unclear | a) *Yes | a) *Yes | a) *Automated brain tissue volume computations (e.g. SPM, FreeSurfer) | a) *Statistical test used to analyse the data clearly described, appropriate and measures of association presented, and unit of ROI measure reported | 54.5 |
| Verde 2015 | b) *Somewhat representative of the average in the target group. (non-random sampling) | b) Not justified. | c) No information provided | a) **secure record (Laboratory sample test, .eg CO/saliva samples for tobacco, urine sample for thc) | c) No or Unclear | a) *Yes | b) No or unclear | a) *Automated brain tissue volume computations (e.g. SPM, FreeSurfer) | b) Statistical test not appropriate, not described or incomplete. | 45.5 |
| Wang 2019 | c) Selected group of users/convenience sample. | b) Not justified. | c) No information provided | a) **secure record (Laboratory sample test, .eg CO/saliva samples for tobacco, urine sample for thc) | a) **alcohol and tobacco/cannabis AND Any other drug use | a) *Yes | a) *Yes | a) *Automated brain tissue volume computations (e.g. SPM, freesurfer) | a) *Statistical test used to analyse the data clearly described, appropriate and measures of association presented, and unit of ROI measure reported | 72.7 |
| Wang 2020 | b) *Somewhat representative of the average in the target group. (non-random sampling) | b) Not justified. | c) No information provided | a) **secure record (Laboratory sample test, .eg CO/saliva samples for tobacco, urine sample for thc) | a) **alcohol and tobacco/cannabis AND any other drug use | a) *Yes | a) *Yes | a) *Automated brain tissue volume computations (e.g. SPM, FreeSurfer) | a) *Statistical test used to analyse the data clearly described, appropriate and measures of association presented, and unit of ROI measure reported | 81.8 |
| Yu 2018 | c) Selected group of users/convenience sample. | b) Not justified. | c) No information provided | a) **secure record (Laboratory sample test, .eg CO/saliva samples for tobacco, urine sample for thc) | a) **alcohol and tobacco/cannabis AND any other drug use | a) *Yes | a) *Yes | a) *Automated brain tissue volume computations (e.g. SPM, FreeSurfer) | a) *Statistical test used to analyse the data clearly described, appropriate and measures of association presented, and unit of ROI measure reported | 72.7 |
| Yuan 2016 | c) Selected group of users/convenience sample. | c) No information provided | c) No information provided | a) **secure record (Laboratory sample test, .eg CO/saliva samples for tobacco, urine sample for thc) | b) *alcohol and tobacco/cannabis | b) No or unclear | b) No or unclear | a) *Automated brain tissue volume computations (e.g. SPM, FreeSurfer) | a) *Statistical test used to analyse the data clearly described, appropriate and measures of association presented, and unit of ROI measure reported | 45.5 |
| Zhao 2019 | b) *Somewhat representative of the average in the target group. (non-random sampling) | a) *Justified and satisfactory (including sample size calculation). | c) No information provided | b) *self-report using detailed descriptions (e.g. smoke every day for last month) or self-report from participant living in a place where cannabis/tobacco is legal | c) No or Unclear | a) *Yes | b) No or unclear | a) *Automated brain tissue volume computations (e.g. SPM, FreeSurfer) | b) Statistical test not appropriate, not described or incomplete. | 45.5 |

### Table 14: Co-use cross-sectional risk of bias results

|  | **Exposure** | | | | **Comparability** | | | | **Outcome** | |  |
| --- | --- | --- | --- | --- | --- | --- | --- | --- | --- | --- | --- |
| **Study ID** | **1) Representativeness of the sample** | **2) Sample size** | **3) Non-respondents** | **4) Ascertainment of exposure** | **1) Did the study account for other drug use?** | **2) FOR CANNABIS/CO-USE ONLY: Study described route of administration?** | **3) Study matches groups or controls for Age** | **4) Study matches groups or controls for sex or gender** | **1) Assessment of outcome** | **2) Statistical test** | **%** |
| Filbey 2015 | b) *Somewhat representative of the average in the target group. (non-random sampling) | b) Not justified. | c) No information provided | a) **secure record (Laboratory sample test, .eg CO/saliva samples for tobacco, urine sample for thc) | a) **alcohol and tobacco/cannabis AND any other drug use | b) No or unclear | a) *Yes | a) *Yes | a) *Automated brain tissue volume computations (e.g. SPM, FreeSurfer) | a) *Statistical test used to analyse the data clearly described, appropriate and measures of association presented, and unit of ROI measure reported | 75 |

### Table 15: Cannabis longitudinal risk of bias results

|  | **Exposure** | | | | | **Comparability** | | | | **Outcome** | | | |  |
| --- | --- | --- | --- | --- | --- | --- | --- | --- | --- | --- | --- | --- | --- | --- |
| **Study ID** | **1) Representativeness of the sample** | **2) Sample size** | **3) Selection of the non-exposed cohort** | **4) Ascertainment of exposure** | **5) Demonstration that outcome of interest was not present at start of study** | **1) Did the study account for other drug use?** | **2) Study described route of administration** | **3) Study matches groups or controls for Age** | **4) Study matches groups or controls for sex or gender** | **1) Assessment of outcome** | **2) Is the temporal relationship between exposure/outcome feasible?** | **3) Adequacy of follow up of cohorts** | **4) Statistical test** | **%** |
| Garimella 2020 | b) *Somewhat representative of the average in the target group. (non-random sampling) | b) Not justified. | a) *drawn from the same community as the exposed cohort | a) **secure record (Laboratory sample test, .eg CO/saliva samples for tobacco, urine sample for thc) | b) *Baseline measure and follow up measure of brain volume, change calculated or baseline values adjusted for | c) No or Unclear | a) *Yes | a) *Yes | a) *Yes | a) *Automated brain tissue volume computations (e.g. SPM, FreeSurfer) | a) *Yes | c) follow up rate < 80% and no description of those lost | a) *Statistical test used to analyse the data clearly described, appropriate and measures of association presented, and unit of ROI measure reported | 68.8 |
| Koenders 2017 | b) *Somewhat representative of the average in the target group. (non-random sampling) | b) Not justified. | a) *drawn from the same community as the exposed cohort | a) **secure record (Laboratory sample test, .eg CO/saliva samples for tobacco, urine sample for thc) | b) *Baseline measure and follow up measure of brain volume, change calculated or baseline values adjusted for | c) No or Unclear | a) *Yes | a) *Yes | a) *Yes | b) *Manual tracing blinded | a) *Yes | c) follow up rate < 80% and no description of those lost | a) *Statistical test used to analyse the data clearly described, appropriate and measures of association presented, and unit of ROI measure reported | 68.8 |
| Luo 2022 | b) *Somewhat representative of the average in the target group. (non-random sampling) | b) Not justified. | a) *drawn from the same community as the exposed cohort | b) *self-report using detailed descriptions (e.g. smoke every day for last month) or self-report from participant living in a place where cannabis/tobacco is legal | b) *Baseline measure and follow up measure of brain volume, change calculated or baseline values adjusted for | b) *alcohol and tobacco/cannabis | b) No or unclear | a) *Yes | a) *Yes | a) *Automated brain tissue volume computations (e.g. SPM, FreeSurfer) | a) *Yes | b) *subjects lost to follow up unlikely to introduce bias - small number lost (<20%) | a) *Statistical test used to analyse the data clearly described, appropriate and measures of association presented, and unit of ROI measure reported | 68.8 |
| Rais 2008 | c) Selected group of users/convenience sample. | b) Not justified. | a) *drawn from the same community as the exposed cohort | c) self-report only | b) *Baseline measure and follow up measure of brain volume, change calculated or baseline values adjusted for | c) No or Unclear | b) No or unclear | a) *Yes | a) *Yes | b) *Manual tracing blinded | a) *Yes | d) no statement | a) *Statistical test used to analyse the data clearly described, appropriate and measures of association presented, and unit of ROI measure reported | 43.8 |
| Wang 2021 | c) Selected group of users/convenience sample. | b) Not justified. | a) *drawn from the same community as the exposed cohort | b) *self-report using detailed descriptions (e.g. smoke every day for last month) or self-report from participant living in a place where cannabis/tobacco is legal | b) *Baseline measure and follow up measure of brain volume, change calculated or baseline values adjusted for | c) No or Unclear | b) No or unclear | a) *Yes | a) *Yes | a) *Automated brain tissue volume computations (e.g. SPM, FreeSurfer) | a) *Yes | b) *subjects lost to follow up unlikely to introduce bias - small number lost (<20%) | a) *Statistical test used to analyse the data clearly described, appropriate and measures of association presented, and unit of ROI measure reported | 56.3 |
| Welch 2011 | b) *Somewhat representative of the average in the target group. (non-random sampling) | b) Not justified. | a) *drawn from the same community as the exposed cohort | c) self-report only | c) unclear or No - only took outcome measure at follow up, or only measured exposure at follow up | a) **alcohol and tobacco/cannabis AND Any other drug use | b) No or unclear | a) *Yes | a) *Yes | a) *Automated brain tissue volume computations (e.g. SPM, FreeSurfer) | a) *Yes | b) *subjects lost to follow up unlikely to introduce bias - small number lost (<20%) | a) *Statistical test used to analyse the data clearly described, appropriate and measures of association presented, and unit of ROI measure reported | 62.5 |
| Xu 2022 | c) Selected group of users/convenience sample. | b) Not justified. | a) *drawn from the same community as the exposed cohort | b) *self-report using detailed descriptions (e.g. smoke every day for last month) or self-report from participant living in a place where cannabis/tobacco is legal | b) *Baseline measure and follow up measure of brain volume, change calculated or baseline values adjusted for | c) No or Unclear | b) No or unclear | a) *Yes | a) *Yes | a) *Automated brain tissue volume computations (e.g. SPM, FreeSurfer) | a) *Yes | b) *subjects lost to follow up unlikely to introduce bias - small number lost (<20%) | a) *Statistical test used to analyse the data clearly described, appropriate and measures of association presented, and unit of ROI measure reported | 56.3 |

### Table 16: Tobacco longitudinal risk of bias results

|  | **Exposure** | | | | | **Comparability** | | | **Outcome** | | | |  |
| --- | --- | --- | --- | --- | --- | --- | --- | --- | --- | --- | --- | --- | --- |
| **Study ID** | **1) Representativeness of the sample** | **2) Sample size** | **3) Selection of the non-exposed cohort** | **4) Ascertainment of exposure** | **5) Demonstration that outcome of interest was not present at start of study** | **1) Did the study account for other drug use?** | **2) Study matches groups or controls for Age** | **3) Study matches groups or controls for sex or gender** | **1) Assessment of outcome** | **2) Is the temporal relationship between exposure /outcome feasible?** | **3) Adequacy of follow up of cohorts** | **4) Statistical test** | **%** |
| Duriez 2014 longitudinal | b) *Somewhat representative of the average in the target group. (non-random sampling) | b) Not justified. | a) *drawn from the same community as the exposed cohort | c) self-report only | b) *Baseline measure and follow up measure of brain volume, change calculated or baseline values adjusted for | c) No or Unclear | a) *Yes | a) *Yes | a) *Automated brain tissue volume computations (e.g. SPM, FreeSurfer) | a) *Yes | c) follow up rate < 80% and no description of those lost | a) *Statistical test used to analyse the data clearly described, appropriate and measures of association presented, and unit of ROI measure reported | 53.3 |
| Kim 2018 | a) *Truly representative of the average in the target population. (all subjects or random sampling) | b) Not justified. | a) *drawn from the same community as the exposed cohort | c) self-report only | b) *Baseline measure and follow up measure of brain volume, change calculated or baseline values adjusted for | c) No or Unclear | a) *Yes | a) *Yes | a) *Automated brain tissue volume computations (e.g. SPM, FreeSurfer) | a) *Yes | d) no statement | a) *Statistical test used to analyse the data clearly described, appropriate and measures of association presented, and unit of ROI measure reported | 53.3 |
| Otsuka 2022 | a) *Truly representative of the average in the target population. (all subjects or random sampling) | b) Not justified. | a) *drawn from the same community as the exposed cohort | c) self-report only | b) *Baseline measure and follow up measure of brain volume, change calculated or baseline values adjusted for | c) No or Unclear | a) *Yes | a) *Yes | a) *Automated brain tissue volume computations (e.g. SPM, FreeSurfer) | a) *Yes | b) *subjects lost to follow up unlikely to introduce bias - small number lost (<20%) | a) *Statistical test used to analyse the data clearly described, appropriate and measures of association presented, and unit of ROI measure reported | 60 |
| Van Haren 2010 | b) *Somewhat representative of the average in the target group. (non-random sampling) | b) Not justified. | c) no description of the derivation of the non-exposed cohort | b) *self-report using detailed descriptions (e.g. smoke every day for last month) or self-report from participant living in a place where cannabis/tobacco is legal | b) *Baseline measure and follow up measure of brain volume, change calculated or baseline values adjusted for | c) No or Unclear | a) *Yes | a) *Yes | a) *Automated brain tissue volume computations (e.g. SPM, FreeSurfer) | a) *Yes | d) no statement | a) *Statistical test used to analyse the data clearly described, appropriate and measures of association presented, and unit of ROI measure reported | 53.3 |

### Table 17: Quality assessment of Mendelian randomization (MR) studies using summary level data

Using quality assessment from Treur et al., (2021) doi:10.1017/S003329172100180X

| **Study ID** |  | Logtenberg 2022 | Lin 2023 |
| --- | --- | --- | --- |
| **Phenotype measurement** | **Sample size exposure** | + | + |
|  | **Sample size outcome** | -+ | -+ |
|  | **Measure exposure** | - | - |
|  | **Measure outcome** | + | + |
| **Instrument strength** | **P value threshold** | -+ | -+ |
|  | **# SNPs** | + | + |
|  | **Biological knowledge** | - | - |
|  | **F stat reported** | -+ | -+ |
|  | **F stat sufficient** | -+ | -+ |
|  | **% var explained reported** | -+ | -+ |
|  | **% var explained sufficient** | -+ | -+ |
| **Bi-directional effects** |  | -+ | - |
| **Temporality** |  | -+ | -+ |
|  | **Harmonization** | - | - |
|  | **Ethnic group** | -+ | -+ |
| **Sample overlap** | **Reported** | - | -+ |
|  | **Present** | -+ | - |
| **Sensitivity analyses** | **Horizontal pleiotropy** | -+ | -+ |
|  | **Leave-one-out / forest plot** | - | - |
|  | **Additional methods** | + | + |
| **Total** |  | + | + |

### Cannabis cross-sectional adjusted meta-analysis summary

#### Table 18: Summary of results from adjusted cross-sectional meta-analysis of differences in brain volume between people who use cannabis (PWUC) and controls.

| **Region of interest** | **Meta-analysis results** | | | | | | |
| --- | --- | --- | --- | --- | --- | --- | --- |
|  | **k** | **Hedge’s g** | **L95%CI** | **U95%CI** | **t** | **p** | **I^2^ (%)** |
| Accumbens | 10 | -0.04 | -0.22 | 0.15 | -0.44 | 0.673 | 31.40 |
| Amygdala | 17 | 0.13 | 0.03 | 0.23 | 2.70 | 0.016 | 22.90 |
| Brainstem | 3 | -0.03 | -0.69 | 0.63 | -0.22 | 0.843 | 0.36 |
| Caudate | 10 | 0.02 | -0.15 | 0.20 | 0.28 | 0.787 | 24.10 |
| Cerebellum | 6 | -0.01 | -0.31 | 0.30 | -0.04 | 0.967 | 32.60 |
| Hippocampus | 23 | 0.12 | -0.01 | 0.24 | 1.94 | 0.066 | 42.10 |
| Hippocampus CA1 | 3 | 0.42 | -0.44 | 1.28 | 2.12 | 0.168 | 79.20 |
| Hippocampus DG | 3 | 0.31 | -0.41 | 1.02 | 1.86 | 0.205 | 69.80 |
| Hippocampus subiculum | 3 | 0.15 | -0.37 | 0.68 | 1.25 | 0.339 | 34.50 |
| Intracranial volume | 6 | -0.06 | -0.31 | 0.19 | -0.65 | 0.545 | 0.00 |
| Intracranial volume (TF) | 9 | -0.19 | -0.43 | 0.04 | -1.88 | 0.097 | 10.10 |
| Lateral orbitofrontal cortex | 3 | 0.12 | -0.19 | 0.44 | 1.67 | 0.236 | 0.00 |
| Medial orbitofrontal cortex | 3 | 0.38 | -0.55 | 1.31 | 1.76 | 0.220 | 68.70 |
| Pallidum | 5 | -0.21 | -0.69 | 0.27 | -1.22 | 0.290 | 68.10 |
| Prefrontal cortex | 3 | 0.25 | -0.92 | 1.42 | 0.91 | 0.458 | 38.50 |
| Putamen | 10 | -0.14 | -0.53 | 0.26 | -0.77 | 0.460 | 77.10 |
| Rostral middle frontal gyrus | 3 | -0.01 | -1.47 | 1.45 | -0.03 | 0.980 | 76.30 |
| Thalamus | 9 | 0.07 | -0.09 | 0.23 | 1.03 | 0.334 | 29.60 |
| Total brain volume | 7 | 0.08 | -0.07 | 0.23 | 1.34 | 0.229 | 0.00 |
| Total grey matter volume | 9 | 0.14 | -0.08 | 0.36 | 1.43 | 0.191 | 31.20 |
| Total white matter volume | 9 | -0.03 | -0.15 | 0.09 | -0.63 | 0.544 | 0.00 |

TF = Trim and Fill result

#### Table 19: Summary of results from Egger’s test for adjusted cross-sectional meta-analysis of differences in brain volume between people who use cannabis (PWUC) and controls

| **Region of interest** | **k** | **Intercept** | **L95%CI** | **U95%CI** | **t** | **p** |
| --- | --- | --- | --- | --- | --- | --- |
| Accumbens | 10 | -1.68 | -3.20 | -0.16 | -2.17 | 0.062 |
| Amygdala | 17 | -0.86 | -2.08 | 0.36 | -1.38 | 0.187 |
| Brainstem | 3 | -2.66 | -5.10 | -0.21 | -2.13 | 0.280 |
| Caudate | 10 | -1.25 | -2.80 | 0.31 | -1.57 | 0.154 |
| Cerebellum | 6 | -1.07 | -4.02 | 1.89 | -0.71 | 0.519 |
| Hippocampus | 23 | -0.50 | -1.72 | 0.71 | -0.81 | 0.427 |
| Hippocampus CA1 | 3 | 3.75 | -0.32 | 7.81 | 1.81 | 0.322 |
| Hippocampus DG | 3 | 2.98 | -0.95 | 6.90 | 1.49 | 0.377 |
| Hippocampus subiculum | 3 | 0.68 | -3.94 | 5.31 | 0.29 | 0.821 |
| Intracranial volume | 6 | 3.45 | 1.16 | 5.74 | 2.95 | 0.042 |
| Lateral orbitofrontal cortex | 3 | 0.62 | -1.81 | 3.05 | 0.50 | 0.705 |
| Medial orbitofrontal cortex | 3 | 3.13 | 0.31 | 5.94 | 2.18 | 0.274 |
| Pallidum | 5 | -2.77 | -6.28 | 0.74 | -1.55 | 0.220 |
| Prefrontal cortex | 3 | -1.19 | -6.39 | 4.01 | -0.45 | 0.731 |
| Putamen | 10 | -1.29 | -4.31 | 1.74 | -0.83 | 0.428 |
| Rostral middle frontal gyrus | 3 | -1.55 | -19.27 | 16.19 | -0.17 | 0.892 |
| Thalamus | 9 | 0.95 | -0.76 | 2.65 | 1.09 | 0.312 |
| Total brain volume | 7 | -0.67 | -2.51 | 1.18 | -0.71 | 0.509 |
| Total grey matter volume | 9 | 0.81 | -1.73 | 3.35 | 0.62 | 0.554 |
| Total white matter volume | 9 | -0.23 | -1.54 | 1.08 | -0.35 | 0.740 |

### Cannabis cross-sectional unadjusted meta-analysis summary

#### Table 20: Summary of results from unadjusted cross-sectional meta-analysis of differences in brain volume between people who use cannabis (PWUC) and controls

| **Region of interest** | **Meta-analysis results** | | | | | | |
| --- | --- | --- | --- | --- | --- | --- | --- |
|  | **k** | **Hedge’s g** | **L95%CI** | **U95%CI** | **t** | **p** | **I2 (%)** |
| Accumbens | 7 | -0.14 | -0.46 | 0.19 | -1.03 | 0.341 | 62.10 |
| Amygdala | 9 | 0.12 | -0.05 | 0.29 | 1.60 | 0.148 | 18.20 |
| Caudate | 5 | -0.06 | -0.33 | 0.22 | -0.60 | 0.583 | 0.00 |
| Cerebellar grey matter | 5 | -0.06 | -0.37 | 0.26 | -0.49 | 0.647 | 18.10 |
| Cerebellar white matter | 5 | 0.11 | -0.53 | 0.75 | 0.47 | 0.662 | 63.70 |
| Cerebellum | 3 | -0.14 | -1.51 | 1.23 | -0.43 | 0.708 | 60.60 |
| Hippocampus | 15 | 0.28 | 0.09 | 0.47 | 3.12 | 0.008 | 53.30 |
| Hippocampus (TF) | 19 | 0.13 | -0.12 | 0.38 | 1.12 | 0.279 | 68.70 |
| Hippocampus CA1 | 5 | 0.07 | -0.41 | 0.54 | 0.38 | 0.720 | 57.70 |
| Hippocampus CA3 | 3 | 0.07 | -0.71 | 0.85 | 0.38 | 0.740 | 45.30 |
| Hippocampus CA4 | 4 | -0.02 | -0.87 | 0.83 | -0.07 | 0.952 | 74.90 |
| Hippocampus fimbria | 4 | 0.23 | -0.51 | 0.96 | 0.98 | 0.400 | 72.40 |
| Hippocampus fissure | 4 | -0.13 | -0.23 | -0.03 | -4.11 | 0.026 | 0.00 |
| Hippocampus parasubiculum | 3 | -0.11 | -1.32 | 1.21 | -0.38 | 0.741 | 72.20 |
| Hippocampus presubiculum | 5 | 0.03 | -0.44 | 0.50 | 0.19 | 0.862 | 56.60 |
| Hippocampus subiculum | 5 | 0.06 | -0.62 | 0.73 | 0.23 | 0.828 | 75.70 |
| Hippocampus tail | 3 | 0.13 | -0.41 | 0.67 | 1.02 | 0.414 | 23.00 |
| Intracranial volume | 12 | 0.00 | -0.16 | 0.17 | 0.05 | 0.960 | 24.20 |
| Pallidum | 3 | -0.20 | -0.90 | 0.50 | -1.25 | 0.339 | 35.80 |
| Putamen | 5 | -0.18 | -0.73 | 0.37 | -0.91 | 0.413 | 63.00 |
| Thalamus | 5 | -0.11 | -0.34 | 0.12 | -1.36 | 0.245 | 5.00 |
| Total brain volume | 4 | -0.12 | -0.90 | 0.66 | -0.49 | 0.656 | 56.00 |
| Total grey matter volume | 6 | 0.06 | -0.30 | 0.42 | 0.42 | 0.692 | 21.30 |
| Total white matter volume | 6 | -0.16 | -0.47 | 0.16 | -1.27 | 0.259 | 0.00 |

TF = Trim and Fill result

#### Table 21: Summary of results from Egger’s test for unadjusted cross-sectional meta-analysis of differences in brain volume between people who use cannabis (PWUC) and controls

| **Region of interest** | **k** | **Intercept** | **L95%CI** | **U95%CI** | **t** | **p** |
| --- | --- | --- | --- | --- | --- | --- |
| Accumbens | 7 | -2.28 | -6.81 | 2.24 | -0.99 | 0.368 |
| Amygdala | 9 | -0.11 | -2.05 | 1.83 | -0.11 | 0.916 |
| Caudate | 5 | 0.72 | -3.41 | 4.84 | 0.34 | 0.756 |
| Cerebellar grey matter | 5 | -0.04 | -3.00 | 2.93 | -0.02 | 0.983 |
| Cerebellar white matter | 5 | -0.54 | -4.91 | 3.82 | -0.24 | 0.823 |
| Cerebellum | 3 | -0.45 | -6.42 | 5.52 | -0.15 | 0.907 |
| Hippocampus | 15 | 1.93 | 0.38 | 3.48 | 2.43 | 0.030 |
| Hippocampus CA1 | 5 | -0.10 | -5.22 | 5.01 | -0.04 | 0.971 |
| Hippocampus CA3 | 3 | 1.23 | -4.57 | 7.04 | 0.42 | 0.749 |
| Hippocampus CA4 | 4 | 0.11 | -7.00 | 7.21 | 0.03 | 0.979 |
| Hippocampus fimbria | 4 | 3.11 | -2.13 | 8.34 | 1.16 | 0.365 |
| Hippocampus fissure | 4 | 0.76 | 0.33 | 1.20 | 3.46 | 0.074 |
| Hippocampus parasubiculum | 3 | -0.95 | -9.51 | 7.60 | -0.22 | 0.863 |
| Hippocampus presubiculum | 5 | 0.31 | -4.73 | 5.35 | 0.12 | 0.912 |
| Hippocampus subiculum | 5 | 0.72 | -5.90 | 7.34 | 0.21 | 0.846 |
| Hippocampus tail | 3 | -0.23 | -5.54 | 5.07 | -0.09 | 0.945 |
| Intracranial volume | 12 | -0.31 | -1.77 | 1.15 | -0.42 | 0.684 |
| Pallidum | 3 | -1.19 | -11.07 | 8.68 | -0.24 | 0.852 |
| Putamen | 5 | 0.11 | -7.12 | 7.33 | 0.03 | 0.979 |
| Thalamus | 5 | 1.40 | -0.45 | 3.25 | 1.48 | 0.236 |
| Total brain volume | 4 | -14.29 | -25.89 | -2.68 | -2.41 | 0.137 |
| Total grey matter volume | 6 | 6.43 | -4.02 | 16.89 | 1.21 | 0.294 |
| Total white matter volume | 6 | -0.48 | -10.41 | 9.45 | -0.10 | 0.929 |

### Tobacco cross-sectional adjusted meta-analysis summary

#### Table 22: Summary of results from adjusted cross-sectional meta-analysis of differences in brain volume between people who smoke tobacco (PWST) and controls

| **Region of interest** | **Meta-analysis results** | | | | | | |
| --- | --- | --- | --- | --- | --- | --- | --- |
|  | **k** | **Hedge’s g** | **L95%CI** | **U95%CI** | **t** | **p** | **I2 (%)** |
| Accumbens | 6 | 0.07 | -0.10 | 0.24 | 1.03 | 0.350 | 60.90 |
| Accumbens (TF) | 9 | 0.16 | -0.07 | 0.38 | 1.62 | 0.143 | 68.20 |
| Amygdala | 5 | 0.17 | 0.04 | 0.31 | 3.48 | 0.025 | 55.90 |
| Caudate | 8 | -0.13 | -0.37 | 0.12 | -1.21 | 0.267 | 72.80 |
| Caudate (TF) | 12 | 0.07 | -0.22 | 0.35 | 0.51 | 0.621 | 78.00 |
| Frontal grey matter | 3 | 0.09 | -0.12 | 0.30 | 1.87 | 0.203 | 0.00 |
| Hippocampus | 10 | 0.12 | 0.00 | 0.24 | 2.27 | 0.049 | 62.30 |
| Insula | 5 | 0.17 | 0.06 | 0.27 | 4.49 | 0.011 | 31.20 |
| Insula (TF) | 7 | 0.21 | 0.09 | 0.32 | 4.37 | 0.005 | 47.10 |
| Pallidum | 5 | 0.17 | 0.13 | 0.21 | 11.88 | 0.000 | 0.00 |
| Parietal grey matter | 3 | 0.31 | -0.59 | 1.22 | 1.50 | 0.273 | 61.60 |
| Parietal grey matter (TF) | 5 | 0.02 | -0.69 | 0.72 | 0.06 | 0.953 | 68.10 |
| Putamen | 8 | 0.08 | -0.05 | 0.20 | 1.49 | 0.179 | 48.30 |
| Putamen (TF) | 11 | 0.15 | -0.02 | 0.31 | 1.98 | 0.076 | 59.60 |
| Temporal grey matter | 3 | 0.16 | -0.25 | 0.57 | 1.70 | 0.232 | 0.00 |
| Thalamus | 12 | 0.13 | -0.06 | 0.31 | 1.52 | 0.158 | 55.50 |
| Total brain volume | 4 | 0.02 | -0.01 | 0.05 | 1.86 | 0.160 | 0.00 |
| Total grey matter volume | 7 | 0.17 | 0.04 | 0.30 | 3.13 | 0.020 | 93.70 |
| Total white matter volume | 4 | 0.01 | -0.03 | 0.05 | 0.62 | 0.577 | 0.00 |

TF = Trim and Fill result

#### Table 23: Summary of results from Egger’s test for adjusted cross-sectional meta-analysis of differences in brain volume between people who smoke tobacco (PWST) and controls

| **Region of interest** | **k** | **Intercept** | **L95%CI** | **U95%CI** | **t** | **p** |
| --- | --- | --- | --- | --- | --- | --- |
| Accumbens | 6 | -2.15 | -3.05 | -1.24 | -4.67 | 0.010 |
| Amygdala | 5 | -1.93 | -3.66 | -0.21 | -2.20 | 0.115 |
| Caudate | 8 | -3.42 | -5.27 | -1.58 | -3.63 | 0.011 |
| Frontal grey matter | 3 | 0.49 | -0.80 | 1.78 | 0.75 | 0.592 |
| Hippocampus | 10 | 0.59 | -1.23 | 2.41 | 0.64 | 0.540 |
| Insula | 5 | -1.77 | -2.75 | -0.80 | -3.55 | 0.038 |
| Pallidum | 5 | -0.12 | -1.07 | 0.83 | -0.24 | 0.824 |
| Parietal grey matter | 3 | 2.60 | 2.57 | 2.63 | 167.22 | 0.004 |
| Putamen | 8 | -1.57 | -2.54 | -0.60 | -3.18 | 0.019 |
| Temporal grey matter | 3 | 1.44 | 0.92 | 1.97 | 5.39 | 0.117 |
| Thalamus | 12 | 0.09 | -1.79 | 1.96 | 0.09 | 0.930 |
| Total brain volume | 4 | 0.55 | 0.23 | 0.86 | 3.42 | 0.076 |
| Total grey matter volume | 7 | 0.76 | -4.63 | 6.14 | 0.28 | 0.794 |
| Total white matter volume | 4 | 0.38 | -0.47 | 1.23 | 0.87 | 0.475 |

### Tobacco cross-sectional unadjusted meta-analysis summary

#### Table 24: Summary of results from unadjusted cross-sectional meta-analysis of differences in brain volume between people who smoke tobacco (PWST) and controls

| **Region of interest** | **Meta-analysis results** | | | | | | |
| --- | --- | --- | --- | --- | --- | --- | --- |
|  | **k** | **Hedge’s g** | **L95%CI** | **U95%CI** | **t** | **p** | **I2 (%)** |
| Accumbens | 3 | -0.02 | -0.87 | 0.82 | -0.11 | 0.922 | 61.60 |
| Amygdala | 3 | 0.03 | -0.41 | 0.48 | 0.33 | 0.775 | 0.00 |
| Caudate | 3 | 0.04 | -1.24 | 1.32 | 0.15 | 0.896 | 84.00 |
| Hippocampus | 4 | 0.19 | -0.23 | 0.61 | 1.43 | 0.249 | 38.90 |
| Intracranial volume | 22 | -0.07 | -0.22 | 0.09 | -0.89 | 0.383 | 71.30 |
| Pallidum | 4 | -0.05 | -0.38 | 0.28 | -0.47 | 0.673 | 0.00 |
| Putamen | 3 | -0.04 | -0.69 | 0.60 | -0.28 | 0.806 | 35.90 |
| Thalamus | 5 | -0.24 | -1.11 | 0.63 | -0.78 | 0.479 | 87.30 |
| Total brain volume | 4 | 0.11 | -0.31 | 0.52 | 0.82 | 0.473 | 18.60 |
| Total grey matter volume | 3 | 0.19 | -0.56 | 0.93 | 1.07 | 0.398 | 62.30 |
| Total white matter volume | 3 | 0.28 | -0.40 | 0.96 | 1.79 | 0.216 | 49.50 |

#### Table 25: Summary of results from Egger’s test for unadjusted cross-sectional meta-analysis of differences in brain volume between people who smoke tobacco (PWST)) and controls

| **Region of interest** | **k** | **Intercept** | **L95%CI** | **U95%CI** | **t** | **p** |
| --- | --- | --- | --- | --- | --- | --- |
| Accumbens | 3 | 3.64 | -9.98 | 17.26 | 0.52 | 0.693 |
| Amygdala | 3 | 4.29 | 2.30 | 6.28 | 4.22 | 0.148 |
| Caudate | 3 | 7.96 | -10.35 | 26.28 | 0.85 | 0.551 |
| Hippocampus | 4 | 2.71 | -4.74 | 10.17 | 0.71 | 0.550 |
| Intracranial volume | 22 | 0.06 | -1.08 | 1.21 | 0.11 | 0.916 |
| Pallidum | 4 | 0.36 | -1.86 | 2.59 | 0.32 | 0.779 |
| Putamen | 3 | 4.39 | -3.99 | 12.78 | 1.03 | 0.491 |
| Thalamus | 5 | -1.63 | -14.30 | 11.04 | -0.25 | 0.817 |
| Total brain volume | 4 | -0.85 | -3.45 | 1.74 | -0.65 | 0.585 |
| Total grey matter volume | 3 | 3.88 | -15.59 | 23.35 | 0.39 | 0.763 |
| Total white matter volume | 3 | 6.83 | -4.22 | 17.88 | 1.21 | 0.440 |

### Tobacco longitudinal adjusted meta-analysis summary

#### Table 26: Summary of results from adjusted longitudinal meta-analysis of differences in brain volume between people who smoke tobacco (PWST) and controls.

A positive value indicates a greater reduction in PWUT compared to controls (change controls – change PWST).

| **Region of interest** | **Meta-analysis results** | | | | | | |
| --- | --- | --- | --- | --- | --- | --- | --- |
|  | **k** | **Hedge’s g** | **L95%CI** | **U95%CI** | **t** | **p** | **I2 (%)** |
| Total brain volume | 3 | 0.11 | -0.11 | 0.32 | 2.11 | 0.170 | 0.00 |
| Total grey matter volume | 5 | 0.05 | 0.01 | 0.10 | 3.60 | 0.037 | 0.00 |

#### Table 27: Summary of results from Egger’s test for adjusted longitudinal meta-analysis of differences in brain volume between people who smoke tobacco (PWST) and controls.

| **Region of interest** | **k** | **Intercept** | **L95%CI** | **U95%CI** | **t** | **p** |
| --- | --- | --- | --- | --- | --- | --- |
| Total brain volume | 3 | -2.72 | -3.46 | -1.97 | -7.15 | 0.089 |
| Total grey matter volume | 5 | -0.50 | -0.78 | -0.22 | -3.52 | 0.072 |

### Table 28: Cannabis cross-sectional narrative summary for studies not included in any meta-analysis

| **Study ID** | **Main results relevant to review** |
| --- | --- |
| Lisdahl 2016 | People who used cannabis showed significantly smaller left hippocampal volumes (b = -0.18, p – 0.04, FDR corrected p = 0.20). |
| Maple 2019 | Cannabis use was associated with significantly smaller left Rostral Anterior Cingulate Cortex volume (ACC) (t (48) = -3/09, b = -0.33, p = 0.003, FDR corrected p = 0.02). There were no significant differences for the bilateral medial orbitofrontal cortex (OFC), lOFC, pars triangularis, superior temporal, fusiform, rostral ACC, caudal ACC, amygdala, hippocampus or cerebellum. |
| Rapp 2013 | Results demonstrated a significant negative association of cannabis use on the posterior cingulate cortex (PCC), (F (1, 51) = 8.96, partial n^2^ = 0.15, p = 0.004), independent of diagnostic group or hemisphere. There was a trend for an interaction between cannabis use and hemisphere for the ACC, (F (1,51) = 3.95, partial n^2^ = 0.07, p = 0.052), post-hoc comparisons showed a significant effect of cannabis use on the left ACC (F (1,51) = 7.02, partial n^2^ = 0.12, p = 0.011). |
| Scott 2019 | Results showed no significant differences associated with cannabis use for any global or regional brain volume measure. |
| Szeszko 2007 | Patients who used cannabis had significantly smaller volumes of the ACC compared to patients who did not use cannabis (t (1,108) = -2.41, p = 0.018) and controls (t (1, 108) = -2.19, p = 0.031). |
| Thayer 2019b | There were no significant differences in ICV, TBV, TGMV, TWMV in People who use cannabis (all p> 0.13).  People who used cannabis showed greater volume in the left putamen (F (1,53) = 11.49, FDR corrected p = 0.02) than controls but here were no differences for any other subcortical structure. |

### Table 29: Tobacco cross-sectional narrative summary for studies not included in any meta-analysis

| **Study ID** | **Main results relevant to review** |
| --- | --- |
| Chen 2006 | There were no significant differences in relative grey matter volume (a ratio of TGMV to ICV) between people who smoke compared to non-smokers in males (p = 0.401) or females (p = 0.216). |
| Hoogendam 2012 | There were no differences between people who currently smoke, and never smokers in cerebellar volume (SMD = -0.026, 95% CI [-0.093, 0.040]), but there was a significant association for cerebral volume being smaller in people who smoke (SMD = -0.054, 95% CI [-0.092, -0.016]). |
| Jha 2021 | Results identified no significant differences in habenular volume between people who smoke and controls (t (6910 = 0.24, p = 0.814). |
| Lin 2019 | There was no significant difference in ICV between people who heavily smoked and non-smokers (Cohen’s d = 0.155, p = 0.429). |
| Paul 2008 | Results showed no significant differences between people who chronically smoke and non-smokers in white matter volume of the whole corpus callosum (t (18) = 520.40, p = >.05) or across the three subregions (F = 50.71, p = >.05). |
| Pennington 2015 | In people who used multiple substances, those who also smoked had greater right OFC volumes (p <0.035), and trends for larger ACC volume (p <0.053), compared to those who didn’t also smoke. |
| Valsdóttir 2022 | Smoking was associated with smaller relative grey matter volume (b = -0.011, p <0.001). |
| Verde 2015 | The results suggest people who smoke had reduced volume in the left hippocampus compared to non-smokers, and greater right middle posterior gyrus and sulcal volume. |
| Zhao 2019 | Results showed that people who smoked had reduced cortical volume compared to non-smokers. People who currently smoked also had  smaller volumes in the left parahippocampus and inferior temporal cortex. |
| Wang 2019 | There were no significant differences in volume of the anterior and posterior insula between people who smoke, relapse and non-smokers. |

### Table 30: Cannabis longitudinal narrative summary

| **Study ID** | **Main results relevant to review** |
| --- | --- |
| Garimella 2020 | Greater increases in volume between baseline to follow up of the right parasubiculum, right fimbria, and the CA3 in both hemispheres, in people who use cannabis compared to controls. There were no differences in volume changes between baseline to follow up in any other hippocampus subfield volumes, cannabis use was not associated with greater atrophy. |
| Koenders 2017 | There were no differences in hippocampus volume between people who use cannabis and non-users at baseline and follow up. Hippocampus volume increased between baseline to follow up, and there was no difference in this increase between groups (F (1,39) = 0.01, p = 0.93). |
| Luo 2022 | Alcohol, cannabis and alcohol-cannabis co-use were significantly associated with annual volume changes in the caudal middle frontal cortex (cMFC, Bonferroni-corrected *p*-value = 0.013), fusiform gyrus (FG, *p*-value = 0.006), inferior frontal gyrus, pars opercularis (IFGpo, *p*-value = 0.022), superior temporal gyrus (STG, *p*-value = 0.002), and supramarginal gyrus (SMG, *p*-value < 0.001). There were no differences in rate of volumetric decline between cannabis using group and the non-using group in cortical regions. Alcohol and cannabis co-use was associated with faster declines in the cMFC than non-users (−0.094 cm^3^/year, Bonferroni-corrected *p*-value < 0.001), FG (−0.106, *p*-value < 0.001), IFGpo (−0.046, *p*-value = 0.003), STG (−0.131, *p*-value < 0.001), and SMG (−0.138, *p*-value < 0.001). |
| Rais 2008 | Patients with schizophrenia who currently used cannabis showed significantly larger total grey matter volume loss compared to non-using patients (mean difference = –2.67%; se = 1.21%, p = 0.03) and non-using controls (mean difference = –5.09%; se = 1.28%, p <0.001). |
| Wang 2021 | Bilateral, right hippocampus and hippocampus subfield volumes, CA1, CA4/DG, SR/SL/SM and the subiculum significant decreased from baseline to follow up, but not the left hippocampus and CA2/CA3. There was no main effect of cannabis use group, but there was a significant interaction between time and group, for the relative right hippocampal volume (*F* (40) = 7.16, *p* = 0.011) and right CA1 (*F* (40) = 6.25, *p* = 0.017). There was a greater rate of decrease from baseline to follow up for the right hippocampus (*t* = -2.46, *p* = 0.018, Cohen's *d* = -0.769 and right CA1 (*t* = -2.46, *p* = 0.018, Cohen's *d* = -0.768) in the group of people who used cannabis, compared to controls. |
| Welch 2011 | In people at familial high risk for schizophrenia, there was greater right (F = 7.66, P = 0.008) and left (F = 4.47, p = 0.04) thalamic volume loss from baseline to follow up, in the group who used cannabis compared to the non-using group. |
| Xu 2022 | There were no significant interactions between group (cannabis use vs non-use) and time on neocortical grey matter volume, total grey matter volume, white matter volume, total intercranial volume and volumes of subcortical structures. |

### Table 31: Tobacco longitudinal narrative summary

| **Study ID** | **Results** |
| --- | --- |
| Van Haren 2010 | There was no difference between smokers and non-smokers in changes in brain volume between baseline to follow up in the cerebrum (b = 3.01, se = 4.90, p = 0.54), cerebellum (b = -0.85, se = 0.73, p = 0.25), cerebral grey matter (b = -2.00, se = 5.08, p = -0.40) and cerebral white matter (b = -1.01, se = 4.34, p = 0.82) in healthy individuals. There was no difference between smokers and non-smokers in changes in brain volume between baseline to follow up in the cerebrum (b = -6.60, se = 7.00, p = 0.34), cerebellum (b = 0.36, se = 1.04, p = 0.73), cerebral grey matter (b = -0.02, se = 7.21, p = 1.00) and cerebral white matter (b = -6.57, se = 6.16, p = 0.29) in patients with schizophrenia. |
| Duriez 2014 | The annual rate of volume loss for whole brain volume was greater in male current smokers, than male non-smokers (*p* = 0.0094), but there were no differences for women. The annual rate of volume loss for grey matter volume was not different between smoking groups (men *p* = 0.57, women *p* = 0.55), nor for white matter volume (men = *p* = 0.10, women p = 0.47). The annual rate of volume loss in the hippocampus was greater in female smokers compared to female non-smokers (*p* = 0.0053), but differences between user groups were not significant for men ((*p* = 0.078). |
| Kim 2018 | In men, there was a greater volume decrease for current smokers (b = −0.673, se = 0.012, p = 0.013) compared to never smokers. Smoking was not assessed in women due to small sample size. |
| Otsuka 2022 | There was a greater grey matter volume decreased between baseline to follow up in male current smokers compared to non-smokers (b = 0.349, se = 0.16, p = 0.030), but this difference was not seen in women (b = 0.115, se = 0.29, p = 0.693). |

### Table 32: Studies excluded at full-text screening

| **Study** | **Title** | **DOI** | **Exclusion reason** |
| --- | --- | --- | --- |
| Aasly 1993 | Minor structural brain changes in young drug abusers: A magnetic resonance study | 10.1111/j.1600-0404.1993.tb04103.x | Wrong exposure |
| Abush 2018 | Associations between adolescent cannabis use and brain structure in psychosis | 10.1016/j.pscychresns.2018.03.008 | Wrong exposure |
| Akkermans 2017 | Effect of tobacco smoking on frontal cortical thickness development: A longitudinal study in a mixed cohort of ADHD-affected and -unaffected youth | 10.1016/j.euroneuro.2017.07.007 | Wrong exposure |
| Albaugh 2021 | Association of Cannabis Use during Adolescence with Neurodevelopment | 10.1001/jamapsychiatry.2021.1258 | Wrong exposure |
| Albaugh 2023 | Differential associations of adolescent versus young adult cannabis initiation with longitudinal brain change and behavior | 10.1038/s41380-023-02148-2 | Wrong exposure |
| AlejandraInfante 2018 | Adolescent brain surface area pre-and post-cannabis and alcohol initiation | 10.15288/jsad.2018.79.835 | Wrong outcomes |
| Alkan 2023 | Investigation of the effect of ‚ÄúNicotiana rustica/Mara≈ü Otu‚Äù use on gray matter using image processing techniques from brain MRI images | 10.1007/s11760-023-02572-5 | Wrong method (not t-1 weighted MRI) |
| Almeida 2008 | Smoking is associated with reduced cortical regional gray matter density in brain regions associated with incipient alzheimer disease | 10.1097/JGP.0b013e318157cad2 | Wrong outcomes |
| Almeida 2011 | 24-Month effect of smoking cessation on cognitive function and brain structure in later life | 10.1016/j.neuroimage.2011.01.063 | Wrong outcomes |
| Alshehri 2023 | The impact of cigarette smoking on cognitive processing speed and brain atrophy in multiple sclerosis | 10.1177/13524585231172490 | Wrong outcomes |
| Altamura 2017 | Structural and metabolic differentiation between bipolar disorder with psychosis and substance-induced psychosis: An integrated MRI/PET study | 10.1016/j.eurpsy.2016.09.009 | Wrong exposure |
| Ang 2022 | Cannabis (Marijuana) | 10.1007/978-3-031-08774-5_4 | Wrong study design |
| Bagga 2019 | Investigating Sex-Specific Characteristics of Nicotine Addiction Using Metabolic and Structural Magnetic Resonance Imaging | 10.1159/000494260 | Wrong outcomes |
| Bangalore 2008 | Cannabis use and brain structural alterations in first episode schizophrenia - A region of interest, voxel based morphometric study | 10.1016/j.schres.2007.11.029 | Wrong exposure |
| Batalla 2014 | Modulation of brain structure by catechol-O-methyltransferase Val 158Met polymorphism in chronic cannabis users | 10.1111/adb.12027 | Overlapping samples |
| Battistella 2014 | Long-term effects of cannabis on brain structure | 10.1038/npp.2014.67 | Wrong comparator |
| BekinSarƒ±kaya 2023 | Deficits in peripheric and central olfactory measurements in smokers: evaluated by cranial MRI | 10.1007/s00405-022-07700-4 | Wrong outcomes |
| Bell 2024 | Chronic cannabis use associated with subcortical topological reorganization of structural connectivity in adults | 10.1016/j.drugalcdep.2024.111405 | Wrong outcomes |
| Binnewies 2021 | Associations between depression, lifestyle and brain structure: A longitudinal MRI study | 10.1016/j.neuroimage.2021.117834 | Wrong exposure |
| Bjork 2003 | Cross-sectional volumetric analysis of brain atrophy in alcohol dependence: Effects of drinking history and comorbid substance use disorder | 10.1176/appi.ajp.160.11.2038 | Wrong exposure |
| Blumberg 2003 | Amygdala and Hippocampal Volumes in Adolescents and Adults with Bipolar Disorder | 10.1001/archpsyc.60.12.1201 | Wrong exposure |
| Brown 2023 | Associations Between the Wisconsin Inventory of Smoking Dependence Motives and Regional Brain Volumes in Adult Smokers | 10.1093/ntr/ntad097 | Wrong exposure |
| Brown 2023 | Associations between right inferior frontal gyrus morphometry and inhibitory control in individuals with nicotine dependence | 10.1016/j.drugalcdep.2023.109766 | Wrong comparator |
| Bu 2016 | Functional connectivity abnormalities of brain regions with structural deficits in young adult male smokers | 10.3389/fnhum.2016.00494 | Wrong outcomes |
| Bull 1971 | Cerebral atrophy in young cannabis smokers. | 10.1016/s0140-6736(71)90691-x | Wrong method (not t-1 weighted MRI) |
| Burggren 2018 | Subregional Hippocampal Thickness Abnormalities in Older Adults with a History of Heavy Cannabis Use | 10.1089/can.2018.0035 | Wrong exposure |
| Cai 2022 | To explore the mechanism of tobacco addiction using structural and functional MRI: a preliminary study of the role of the cerebellum-striatum circuit | 10.1007/s11682-021-00546-0 | Wrong outcomes |
| Campbell 2023 | Distributed Genetic Effects of the Corpus Callosum Subregions Suggest Links to Neuropsychiatric Disorders and Related Traits | 10.1017/neu.2023.32 | Wrong study design |
| Cao 2021 | Mapping cortical and subcortical asymmetries in substance dependence: Findings from the ENIGMA Addiction Working Group | 10.1111/adb.13010 | Wrong outcomes |
| Chaarani 2019 | Low Smoking Exposure, the Adolescent Brain, and the Modulating Role of CHRNA5 Polymorphisms | 10.1016/j.bpsc.2019.02.006 | Wrong exposure |
| Chang 2024 | Investigating the Relationship Between Smoking Behavior and Global Brain Volume | 10.1016/j.bpsgos.2023.09.006 | Wrong exposure |
| Chen 2024 | The interaction effects of age, APOE and common environmental risk factors on human brain structure | 10.1093/cercor/bhad472 | Wrong exposure |
| Chirokoff 2024 | Identifying the role of (dis)inhibition in the vicious cycle of substance use through ecological momentary assessment and resting-state fMRI | 10.1038/s41398-024-02949-1 | Wrong outcomes |
| Chumachenko 2015 | Brain cortical thickness in male adolescents with serious substance use and conduct problems | 10.3109/00952990.2015.1058389 | Wrong exposure |
| Chye 2017a | Orbitofrontal and caudate volumes in cannabis users: a multi-site mega-analysis comparing dependent versus non-dependent users | 10.1007/s00213-017-4606-9 | Overlapping samples |
| Chye 2019a | Alteration to hippocampal volume and shape confined to cannabis dependence: a multi-site study | 10.1111/adb.12652 | Overlapping samples |
| Chye 2019b | Cortical surface morphology in long-term cannabis users: A multi-site MRI study | 10.1016/j.euroneuro.2018.11.1110 | Overlapping samples |
| Cole 2020 | Multimodality neuroimaging brain-age in UK biobank: relationship to biomedical, lifestyle, and cognitive factors | 10.1016/j.neurobiolaging.2020.03.014 | Wrong outcomes |
| Cong 2022 | Individuals with cannabis use are associated with widespread morphological alterations in the subregions of the amygdala, hippocampus, and pallidum | 10.1016/j.drugalcdep.2022.109595 | Overlapping samples |
| Conti 2021 | Neuroanatomical Correlates of Impulsive Choices and Risky Decision Making in Young Chronic Tobacco Smokers: A Voxel-Based Morphometry Study | 10.3389/fpsyt.2021.708925 | Wrong outcomes |
| Conti 2022 | Chronic tobacco smoking, impaired reward-based decision-making, and role of insular cortex: A comparison between early-onset smokers and late-onset smokers | 10.3389/fpsyt.2022.939707 | Wrong outcomes |
| Copersino 2020 | Interactive effects of age and recent substance use on striatal shape morphology at substance use disorder treatment entry | 10.1016/j.drugalcdep.2019.107728 | Wrong exposure |
| Corley 2019 | Epigenetic signatures of smoking associate with cognitive function, brain structure, and mental and physical health outcomes in the Lothian Birth Cohort 1936 | 10.1038/s41398-019-0576-5 | Wrong exposure |
| Courtney 2020 | The effects of nicotine and cannabis co-use during adolescence and young adulthood on white matter cerebral blood flow estimates | 10.1007/s00213-020-05640-7 | Wrong outcomes |
| Courtney 2022 | The Effects of Nicotine and Cannabis Co-Use During Late Adolescence on White Matter Fiber Tract Microstructure | 10.15288/jsad.2022.83.287 | Wrong outcomes |
| Cousijn 2012 | Grey matter alterations associated with cannabis use: Results of a VBM study in heavy cannabis users and healthy controls | 10.1016/j.neuroimage.2011.09.046 | Overlapping samples |
| Cox 2019 | Associations between vascular risk factors and brain MRI indices in UK Biobank | 10.1093/eurheartj/ehz100 | Wrong exposure |
| Crunelle 2014 | Reduced frontal brain volume in non-treatment-seeking cocaine-dependent individuals: Exploring the role of impulsivity, depression, and smoking | 10.3389/fnhum.2014.00007 | Wrong exposure |
| Cunha 2013 | Corrigendum to "Cannabis use, cognition and brain structure in first-episode psychosis" [Schizophr. Res., 147 (2013), 209-215] | 10.1016/j.schres.2013.10.010 | Wrong exposure |
| Cunha 2013 | Cannabis use, cognition and brain structure in first-episode psychosis | 10.1016/j.schres.2013.04.009 | Wrong exposure |
| Dai 2022 | Longitudinal Assessments of Neurocognitive Performance and Brain Structure Associated With Initiation of Tobacco Use in Children, 2016 to 2021 | 10.1001/jamanetworkopen.2022.25991 | Wrong exposure |
| Daniju 2022 | Prefrontal cortex and putamen grey matter alterations in cannabis and tobacco users | 10.1177/02698811221117523 | Wrong outcomes |
| Das 2012 | Lifetime cigarette smoking is associated with striatal volume measures | 10.1111/j.1369-1600.2010.00301.x | Wrong exposure |
| Dekker 2010 | Cannabis use and callosal white matter structure and integrity in recent-onset schizophrenia | 10.1016/j.pscychresns.2009.06.003 | Wrong exposure |
| Delisi 2006 | A preliminary DTI study showing no brain structural change associated with adolescent cannabis use | 10.1186/1477-7517-3-17 | Wrong exposure |
| delRe 2023 | Characterization of childhood trauma, hippocampal mediation and Cannabis use in a large dataset of psychosis and non-psychosis individuals | 10.1016/j.schres.2023.03.029 | Wrong outcomes |
| Demirakca 2010 | Diminished gray matter in the hippocampus of cannabis users: Possible protective effects of cannabidiol | 10.1016/j.drugalcdep.2010.09.020 | Wrong outcomes |
| Dhana 2023 | Cardiovascular health and cognitive outcomes: Findings from a biracial population-based study in the United States | 10.1002/alz.13421 | Wrong exposure |
| Ding 2015 | Multivariate classification of smokers and nonsmokers using SVM-RFE on structural MRI images | 10.1002/hbm.22956 | Wrong outcomes |
| Dong 2022 | The alterations of cerebral cortex thickness based on surface morphology and its correlation with smoking-related characteristics in severe nicotine addicts | 10.3760/cma.j.cn112137-20220706-01499 | Wrong outcomes |
| Dougherty 2020 | Smoking mediates the relationship between SES and brain volume: The CARDIA study | 10.1371/journal.pone.0239548 | Wrong outcomes |
| Durazzo 2007b | Non-treatment-seeking heavy drinkers: Effects of chronic cigarette smoking on brain structure | 10.1016/j.drugalcdep.2006.08.003 | Overlapping samples |
| Durazzo 2012 | Greater regional brain atrophy rate in healthy elderly subjects with a history of cigarette smoking | 10.1016/j.jalz.2011.10.006 | Wrong exposure |
| Durazzo 2014 | Interactive effects of chronic cigarette smoking and age on brain volumes in controls and alcohol-dependent individuals in early abstinence | 10.1111/j.1369-1600.2012.00492.x | Wrong exposure |
| Durazzo 2015 | Serial longitudinal magnetic resonance imaging data indicate non-linear regional gray matter volume recovery in abstinent alcohol-dependent individuals | 10.1111/adb.12180 | Wrong exposure |
| Durazzo 2018 | Cigarette smoking is associated with cortical thinning in anterior frontal regions, insula and regions showing atrophy in early Alzheimer's Disease | 10.1016/j.drugalcdep.2018.08.009 | Wrong outcomes |
| Enzinger 2005 | Risk factors for progression of brain atrophy in aging: Six-year follow-up of normal subjects | 10.1212/01.WNL.0000161871.83614.BB | Wrong exposure |
| Epstein 2015 | Altered cortical maturation in adolescent cannabis users with and without schizophrenia | 10.1016/j.schres.2014.11.029 | Wrong outcomes |
| Ertl 2023 | Associations between regular cannabis use and brain resting-state functional connectivity in adolescents and adults | 10.1177/02698811231189441 | Wrong outcomes |
| Faria 2023 | Reversible lesion in the splenium of the corpus callosum after intense use of cannabis | 10.1007/s10072-023-06772-2 | Wrong study design |
| Faulkner 2021 | Daily and intermittent smoking are associated with low prefrontal volume and low concentrations of prefrontal glutamate, creatine, myo-inositol, and N-acetylaspartate | 10.1111/adb.12986 | Wrong outcomes |
| Filbey 2014 | Long-term effects of marijuana use on the brain | 10.1073/pnas.1415297111 | Wrong outcomes |
| Filbey 2015 | Preliminary findings demonstrating latent effects of early adolescent marijuana use onset on cortical architecture | 10.1016/j.dcn.2015.10.001 | Wrong comparator |
| Filippi 2021 | Neuroimaging evidence for structural correlates in adolescents resilient to polysubstance use: A five-year follow-up study | 10.1016/j.euroneuro.2021.03.001 | Wrong outcomes |
| Francis 2023 | Functional hyperconnectivity between corticocerebellar networks and altered decision making in young adult cannabis users: Evidence from 7T and multivariate pattern analysis | 10.1016/j.pscychresns.2023.111613 | Wrong exposure |
| Francis 2024 | Multimodal 7T imaging reveals enhanced functional coupling between salience and frontoparietal networks in young adult tobacco cigarette smokers | 10.1007/s11682-024-00882-x | Wrong outcomes |
| French 2015 | Early cannabis use, polygenic risk score for schizophrenia, and brain maturation in adolescence | 10.1001/jamapsychiatry.2015.1131 | Wrong outcomes |
| Frissen 2018 | Evidence that reduced gray matter volume in psychotic disorder is associated with exposure to environmental risk factors | 10.1016/j.pscychresns.2017.11.004 | Wrong exposure |
| Fritz 2014 | Current smoking and reduced gray matter volume - A voxel-based morphometry study | 10.1038/npp.2014.112 | Wrong outcomes |
| Froeliger 2010 | Hippocampal and striatal gray matter volume are associated with a smoking cessation treatment outcome: Results of an exploratory voxel-based morphometric analysis | 10.1007/s00213-010-1862-3 | Wrong comparator |
| Gallinat 2006 | Smoking and structural brain deficits: A volumetric MR investigation | 10.1111/j.1460-9568.2006.05050.x | Wrong outcomes |
| Gazdzinski 2010 | Cerebral white matter recovery in abstinent alcoholics - A multimodality magnetic resonance study | 10.1093/brain/awp343 | Wrong exposure |
| Gazula 2021 | Decentralized Multisite VBM Analysis During Adolescence Shows Structural Changes Linked to Age, Body Mass Index, and Smoking: a COINSTAC Analysis | 10.1007/s12021-020-09502-7 | Wrong exposure |
| Ghosh 2022 | Surface-based brain morphometry in schizophrenia vs. cannabis-induced psychosis: A controlled comparison | 10.1016/j.jpsychires.2022.09.034 | Wrong outcomes |
| Gillespie 2018 | Testing associations between cannabis use and subcortical volumes in two large population-based samples | 10.1111/add.14252 | Wrong exposure |
| Gray 2020 | Associations of cigarette smoking with gray and white matter in the UK Biobank | 10.1038/s41386-020-0630-2 | Wrong exposure |
| Grodin 2021 | Effect of Alcohol, Tobacco, and Cannabis Co-Use on Gray Matter Volume in Heavy Drinkers | 10.1037/adb0000743 | Wrong outcomes |
| Guluzade 2022 | RESULTS OF MRI EXAMINATION OF CEREBRAL WHITE MATTER HYPERINTENSITIES IN PATIENTS WITH TYPE 2 DIABETES AND SMOKERS | 10.34921/amj.2022.2.007 | Wrong outcomes |
| Habets 2011 | Reduced cortical thickness as an outcome of differential sensitivity to environmental risks in schizophrenia | 10.1016/j.biopsych.2010.08.010 | Wrong exposure |
| Halcomb 2024 | Greater ventral striatal functional connectivity in cigarette smokers relative to non-smokers across a spectrum of alcohol consumption | 10.1007/s11682-024-00903-9 | Wrong outcomes |
| Haller 2013 | Combined grey matter VBM and white matter TBSS analysis in young first episode psychosis patients with and without cannabis consumption. | 10.1007/s10548-013-0288-8 | Wrong outcomes |
| Hanlon 2016 | Lower subcortical gray matter volume in both younger smokers and established smokers relative to non-smokers | 10.1111/adb.12171 | Wrong outcomes |
| Happer 2024 | Nicotine use during late adolescence and young adulthood is associated with changes in hippocampal volume and memory performance | 10.3389/fnins.2024.1436951 | Wrong exposure |
| Harper 2021 | Orbitofrontal cortex thickness and substance use disorders in emerging adulthood: causal inferences from a co-twin control/discordant twin study | 10.1111/add.15447 | Wrong outcomes |
| Harper 2021 | The Effects of Alcohol and Cannabis Use on the Cortical Thickness of Cognitive Control and Salience Brain Networks in Emerging Adulthood: A Co-twin Control Study | 10.1016/j.biopsych.2021.01.006 | Wrong exposure |
| Harper 2021 | Testing the consequences of alcohol, cannabis, and nicotine use on hippocampal volume: A quasi-experimental cotwin control analysis of young adult twins | 10.1017/S0033291721004682 | Wrong exposure |
| Harper 2021 | The effects of alcohol and cannabis use on inhibitory control brain networks in emerging adulthood: Causal inferences from a cotwin control study. |  | Wrong exposure |
| Harper 2023 | Testing the consequences of alcohol, cannabis, and nicotine use on hippocampal volume: a quasi-experimental cotwin control analysis of young adult twins. | 10.1017/S0033291721004682 | Wrong exposure |
| Hartberg 2018 | Cortical thickness, cortical surface area and subcortical volumes in schizophrenia and bipolar disorder patients with cannabis use | 10.1016/j.euroneuro.2017.11.019 | Wrong exposure |
| Hawkins 2018 | The effect of age and smoking on the hippocampus and memory in late middle age | 10.1002/hipo.23014 | Wrong exposure |
| HernandezMejia 2024 | The Combined Effects of Nicotine and Cannabis on Cortical Thickness Estimates in Adolescents and Emerging Adults | 10.3390/brainsci14030195 | Wrong outcomes |
| Hill 2016 | Lifetime use of cannabis from longitudinal assessments, cannabinoid receptor (CNR1) variation, and reduced volume of the right anterior cingulate | 10.1016/j.pscychresns.2016.05.009 | Wrong exposure |
| Hirjak 2022 | Multimodal MRI data fusion reveals distinct structural, functional and neurochemical correlates of heavy cannabis use | 10.1111/adb.13113 | Wrong outcomes |
| Ho 2011 | Cannabinoid receptor 1 gene polymorphisms and marijuana misuse interactions on white matter and cognitive deficits in schizophrenia | 10.1016/j.schres.2011.02.021 | Wrong comparator |
| Hoefer 2014 | Genetic and behavioral determinants of hippocampal volume recovery during abstinence from alcohol | 10.1016/j.alcohol.2014.08.007 | Wrong exposure |
| Ibrahim 2022 | Anterior cingulate cortex in individuals with depressive symptoms: A structural MRI study | 10.1016/j.pscychresns.2021.111420 | Wrong outcomes |
| Ikram 2008 | Brain tissue volumes in the general elderly population. The Rotterdam Scan Study | 10.1016/j.neurobiolaging.2006.12.012 | Wrong method (not t-1 weighted MRI) |
| J√∏rgensen 2015 | Cigarette smoking is associated with thinner cingulate and insular cortices in patients with severe mental illness | 10.1503/jpn.140163 | Wrong outcomes |
| Jack√≥w-Nowicka 2021 | The Impact of Common Epidemiological Factors on Gray and White Matter Volumes in Magnetic Resonance Imaging‚ÄìIs Prevention of Brain Degeneration Possible? | 10.3389/fneur.2021.633619 | Wrong exposure |
| Jacobus 2014 | Cortical thickness and neurocognition in adolescent marijuana and alcohol users following 28 days of monitored abstinence | 10.15288/jsad.2014.75.729 | Wrong outcomes |
| Jacobus 2015 | Cortical thickness in adolescent marijuana and alcohol users: A three-year prospective study from adolescence to young adulthood | 10.1016/j.dcn.2015.04.006 | Wrong outcomes |
| Jacobus 2016 | Reprint of ‚ÄúAdolescent cortical thickness pre- and post marijuana and alcohol initiation‚Äù | 10.1016/j.ntt.2016.11.003 | Wrong exposure |
| Jacobus 2016 | Adolescent cortical thickness pre- and post marijuana and alcohol initiation | 10.1016/j.ntt.2016.09.005 | Wrong exposure |
| Jager 2007 | Effects of frequent cannabis use on hippocampal activity during an associative memory task | 10.1016/j.euroneuro.2006.10.003 | Wrong outcomes |
| Jakabek 2016 | An MRI study of white matter tract integrity in regular cannabis users: effects of cannabis use and age | 10.1007/s00213-016-4398-3 | Wrong outcomes |
| Jarvis 2008 | Neuroanatomic comparison of bipolar adolescents with and without cannabis use disorders | 10.1089/cap.2008.033 | Wrong exposure |
| Jensen 2019 | The impact of schizophrenia and intelligence on the relationship between age and brain volume | 10.1016/j.scog.2018.09.002 | Wrong exposure |
| Johansson 2023 | Longstanding smoking associated with frontal brain lobe atrophy: a 32-year follow-up study in women | 10.1136/bmjopen-2023-072803 | Wrong outcomes |
| K√ºhn 2010 | Reduced thickness of medial orbitofrontal cortex in smokers | 10.1016/j.biopsych.2010.08.004 | Wrong outcomes |
| K√ºhn 2012 | Brain grey matter deficits in smokers: Focus on the cerebellum | 10.1007/s00429-011-0346-5 | Wrong outcomes |
| Kaag 2018 | The relation between gray matter volume and the use of alcohol, tobacco, cocaine and cannabis in male polysubstance users | 10.1016/j.drugalcdep.2018.03.010 | Wrong exposure |
| Kalayasiri 2023 | The brain activities of individuals with or without motivation to change: a preliminary study among cigarette smokers | 10.37349/emed.2023.00154 | Wrong outcomes |
| KharabianMasouleh 2018 | Gray matter structural networks are associated with cardiovascular risk factors in healthy older adults | 10.1177/0271678X17729111 | Wrong outcomes |
| Kim 2011 | Effects of smoking on cerebral and ventricular volumes in healthy males | 10.3969/j.issn.1673-5374.2011.01.012 | unable to obtain full text |
| Kirsch 2021 | Childhood maltreatment, prefrontal-paralimbic gray matter volume, and substance use in young adults and interactions with risk for bipolar disorder | 10.1038/s41598-020-80407-w | Wrong exposure |
| Koenders 2016 | Grey matter changes associated with heavy cannabis use: A longitudinal sMRI study | 10.1371/journal.pone.0152482 | Wrong outcomes |
| Kokubun 2021 | Unhealthy lifestyles and brain condition: Examining the relations of BMI, living alone, alcohol intake, short sleep, smoking, and lack of exercise with gray matter volume | 10.1371/journal.pone.0255285 | Wrong outcomes |
| Korponay 2022 | Misconfigured striatal connectivity profiles in smokers. | 10.1038/s41386-022-01366-6 | Wrong outcomes |
| Krishnadas 2013 | Cardio-metabolic risk factors and cortical thickness in a neurologically healthy male population: Results from the psychological, social and biological determinants of ill health (pSoBid) study | 10.1016/j.nicl.2013.04.012 | Wrong outcomes |
| Kunas 2020 | The modulating impact of cigarette smoking on brain structure in panic disorder: A voxel-based morphometry study | 10.1093/scan/nsaa103 | Wrong outcomes |
| Kunas 2023 | Brain alterations related to tobacco smoking: Target points for prevention and intervention. |  | Wrong outcomes |
| Lane 2020 | Associations between Vascular Risk Across Adulthood and Brain Pathology in Late Life: Evidence from a British Birth Cohort | 10.1001/jamaneurol.2019.3774 | Wrong exposure |
| Lee 2022 | Thalamocortical functional connectivity and cannabis use in men with childhood attention-deficit/hyperactivity disorder | 10.1371/journal.pone.0278162 | Wrong outcomes |
| Lesh 2024 | Using Task-fMRI to Explore the Relationship Between Lifetime Cannabis Use and Cognitive Control in Individuals With First-Episode Schizophrenia | 10.1093/schizbullopen/sgae016 | Wrong method (not t-1 weighted MRI) |
| Lewis-delosAngeles 2017 | Lower total and regional grey matter brain volumes in youth with perinatally-acquired HIV infection: Associations with HIV disease severity, substance use, and cognition | 10.1016/j.bbi.2017.01.004 | Wrong exposure |
| Li 2020 | Orbitofrontal cortex volume links polygenic risk for smoking with tobacco use in healthy adolescents | 10.1017/S0033291720002962 | Wrong exposure |
| Li 2022 | Orbitofrontal cortex volume links polygenic risk for smoking with tobacco use in healthy adolescents. | 10.1017/S0033291720002962 | Wrong study design |
| Li 2024 | The relationship between intracranial atherosclerosis and white matter hyperintensity in ischemic stroke patients: a retrospective cross-sectional study using high-resolution magnetic resonance vessel wall imaging | 10.21037/qims-23-64 | Wrong outcomes |
| Li 2024 | Abnormal structural covariance networks in young adults with recent cannabis use | 10.1016/j.addbeh.2024.108029 | Wrong outcomes |
| Liao 2012 | Differences between smokers and non-smokers in regional gray matter volumes: A voxel-based morphometry study | 10.1111/j.1369-1600.2010.00250.x | Wrong outcomes |
| Linli 2022 | Associations between smoking and accelerated brain ageing | 10.1016/j.pnpbp.2021.110471 | Wrong outcomes |
| Lippard 2017 | Brain circuitry associated with the development of substance use in bipolar disorder and preliminary evidence for sexual dimorphism in adolescents | 10.1002/jnr.23901 | Wrong exposure |
| Liu 1998 | Smaller volume of prefrontal lobe in polysubstance abusers: A magnetic resonance imaging study | 10.1016/S0893-133X(97)00143-7 | Wrong exposure |
| Liu 2022 | Brain Magnetic Resonance Imaging Features of Nicotine-Dependent Individuals and Its Correlation with Polymorphisms of Dopamine D Receptor Gene. | 10.1155/2022/2296776 | Wrong outcomes |
| Liu 2023 | Brain Structure and Function Show Distinct Relations With Genetic Predispositions to Mental Health and Cognition | 10.1016/j.bpsc.2022.08.003 | Wrong comparator |
| LongstrethJr 2005 | Incidence, manifestations, and predictors of worsening white matter on serial cranial magnetic resonance imaging in the elderly: The cardiovascular health study | 10.1161/01.STR.0000149625.99732.69 | Wrong outcomes |
| Lor 2023 | Thalamic volume and functional connectivity are associated with nicotine dependence severity and craving | 10.1111/adb.13261 | Wrong comparator |
| Lorenzetti 2015 | Gross morphological brain changes with chronic, heavy cannabis use | 10.1192/bjp.bp.114.151407 | Overlapping samples |
| Macedo 2024 | Light Cannabis Use and the Adolescent Brain: An 8-years Longitudinal Assessment of Mental Health, Cognition, and Reward Processing | 10.1007/s00213-024-06575-z | Wrong outcomes |
| Mackey 2019 | Mega-analysis of gray matter volume in substance dependence: General and substance-specific regional effects | 10.1176/appi.ajp.2018.17040415 | Overlapping samples |
| Malchow 2009 | MR volumetric abnormalties in schizophrenia with comorbid substance abuse |  | Wrong comparator |
| Malchow 2013 | Effects of cannabis and familial loading on subcortical brain volumes in first-episode schizophrenia | 10.1007/s00406-013-0451-y | Wrong comparator |
| Manza 2020 | Brain structural changes in cannabis dependence: association with MAGL | 10.1038/s41380-019-0577-z | Wrong exposure |
| Martz 2022 | Polysubstance Use Plays a Key Role in Midlife Structural Brain Alterations in Long-term Cannabis Users | 10.1016/j.biopsych.2022.09.009 | Wrong study design |
| Mason 2022 | The Reward System: What It Is and How It Is Altered in Cannabis Users | 10.1007/978-3-030-92392-1_71 | Wrong study design |
| Mathew 1998 | Marijuana smoking and brain morphology |  | unable to obtain full text |
| Matochik 2005 | Altered brain tissue composition in heavy marijuana users | 10.1016/j.drugalcdep.2004.06.011 | Wrong outcomes |
| McCorkindale 2019 | Re-investigating the effects of chronic smoking on the pathology of alcohol-related human brain damage | 10.1016/j.alcohol.2018.07.001 | Wrong method (not t-1 weighted MRI) |
| McPherson 2021 | Cannabis Affects Cerebellar Volume and Sleep Differently in Men and Women | 10.3389/fpsyt.2021.643193 | Wrong exposure |
| Mo 2023 | Evaluating the causal effect of tobacco smoking on white matter brain aging: a two-sample Mendelian randomization analysis in UK Biobank | 10.1111/add.16088 | Wrong method (not t-1 weighted MRI) |
| Modabbernia 2021 | Linked patterns of biological and environmental covariation with brain structure in adolescence: a population-based longitudinal study | 10.1038/s41380-020-0757-x | Wrong exposure |
| Mon 2014 | Structural brain differences in alcohol-dependent individuals with and without comorbid substance dependence | 10.1016/j.drugalcdep.2014.09.010 | Wrong exposure |
| Morales 2012 | Gray-matter volume in methamphetamine dependence: Cigarette smoking and changes with abstinence from methamphetamine | 10.1016/j.drugalcdep.2012.02.017 | Wrong outcomes |
| Morales 2014 | Cigarette exposure, dependence, and craving are related to insula thickness in young adult smokers | 10.1038/npp.2014.48 | Wrong outcomes |
| Morales 2015 | Gray-matter volume in methamphetamine dependence: Sources of variability and behavioral relevance. |  | Overlapping samples |
| Moser 2018 | Multivariate associations among behavioral, clinical, and multimodal imaging phenotypes in patients with psychosis | 10.1001/jamapsychiatry.2017.4741 | Wrong outcomes |
| Muller 2016 | Late-life brain volume: A life-course approach. The AGES-Reykjavik study | 10.1016/j.neurobiolaging.2016.02.012 | Wrong exposure |
| Mulugeta 2022 | Healthy Lifestyle, Genetic Risk and Brain Health: A Gene-Environment Interaction Study in the UK Biobank | 10.3390/nu14193907 | Wrong comparator |
| Murdoch 2023 | Neuroimaging and immunological features of neurocognitive function related to substance use in people with HIV | 10.1007/s13365-022-01102-2 | Wrong exposure |
| Neth 2020 | Relationship Between Risk Factors and Brain Reserve in Late Middle Age: Implications for Cognitive Aging | 10.3389/fnagi.2019.00355 | Wrong exposure |
| Niu 2023 | Differences in dynamic functional connectivity density in individuals with light and heavy smoking addiction: a study based on functional MR | 10.3760/cma.j.cn112149-20220730-00403 | Wrong outcomes |
| Noauthorshipindicated 2023 | Correction to: ‚ÄúCigarette smoking is associated with thinner cingulate and insular cortices in patients with severe mental illness‚Äù. |  | Wrong outcomes |
| Ohi 2022 | Common brain cortical abnormality in smoking behavior and bipolar disorder: discriminant analysis using cortical thickness and surface area. | 10.1093/cercor/bhab490 | Wrong outcomes |
| Orr 2016 | Recreational marijuana use impacts white matter integrity and subcortical (but not cortical) morphometry | 10.1016/j.nicl.2016.06.006 | Wrong exposure |
| Orr 2019 | Grey matter volume differences associated with extremely low levels of cannabis use in adolescence | 10.1523/JNEUROSCI.3375-17.2018 | Wrong exposure |
| Pagliaccio 2015 | Shared predisposition in the association between cannabis use and subcortical brain structure | 10.1001/jamapsychiatry.2015.1054 | Wrong exposure |
| Pan 2023 | Adherence to a¬†healthy lifestyle and brain structural imaging markers | 10.1007/s10654-023-00992-8 | Wrong comparator |
| Paul 2018 | Does thinner right entorhinal cortex underlie genetic liability to cannabis use? | 10.1017/S0033291718000417 | Wrong exposure |
| Pavisian 2014 | Effects of cannabis on cognition in patients with MS: A psychometric and MRI study | 10.1212/WNL.0000000000000446 | Overlapping samples |
| Peng 2017 | Brain-volume changes in young and middle-aged smokers: a DARTEL-based voxel-based morphometry study | 10.1111/crj.12393 | Wrong outcomes |
| Peng 2018 | Brain structure alterations in respect to tobacco consumption and nicotine dependence: a comparative voxel-based morphometry study | 10.3389/fnana.2018.00043 | Wrong outcomes |
| Penzel 2021 | Association between age of cannabis initiation and gray matter covariance networks in recent onset psychosis | 10.1038/s41386-021-00977-9 | Wrong exposure |
| Pfefferbaum 2018 | Altered brain developmental trajectories in adolescents after initiating drinking | 10.1176/appi.ajp.2017.17040469 | Wrong exposure |
| Prom-Wormley 2015 | Genetic and environmental contributions to the relationships between brain structure and average lifetime cigarette use. | 10.1007/s10519-014-9704-4 | Wrong exposure |
| Purcell 2023 | Neural outcomes of adolescent substance use. |  | Overlapping samples |
| Purcell 2024 | Hippocampal Gray Matter Volume in Young Adulthood Varies With Adolescent Alcohol Use | 10.1037/pha0000722 | Wrong exposure |
| Qi 2020 | The relevance of transdiagnostic shared networks to the severity of symptoms and cognitive deficits in schizophrenia: a multimodal brain imaging fusion study | 10.1038/s41398-020-0834-6 | Wrong exposure |
| Qi 2022 | Cognition, Aryl Hydrocarbon Receptor Repressor Methylation, and Abstinence Duration-Associated Multimodal Brain Networks in Smoking and Long-Term Smoking Cessation | 10.3389/fnins.2022.923065 | Wrong comparator |
| Qian 2013 | MRI study of functional and structural alterations in the brains of smokers | 10.3760/cma.j.issn.1005-1201.2013.09.024 | Wrong study design |
| Qian 2019 | Brain Gray Matter Volume and Functional Connectivity Are Associated With Smoking Cessation Outcomes | 10.3389/fnhum.2019.00361 | Wrong exposure |
| Qiu 2022 | Reciprocal modulation between cigarette smoking and internet gaming disorder on participation coefficient within functional brain networks. | 10.1007/s11682-022-00671-4 | Wrong outcomes |
| Qiu 2024 | Associations of alcohol and tobacco use with psychotic, depressive and developmental disorders revealed via multimodal neuroimaging | 10.1038/s41398-024-03035-2 | Wrong comparator |
| Quinn 2018 | Impact of substance use disorder on gray matter volume in schizophrenia | 10.1016/j.pscychresns.2018.08.002 | Wrong exposure |
| Rabinowitz 2022 | Shared Genetic Etiology between Cortical Brain Morphology and Tobacco, Alcohol, and Cannabis Use | 10.1093/cercor/bhab243 | Wrong study design |
| Rais 2010 | Cannabis use and progressive cortical thickness loss in areas rich in CB1 receptors during the first five years of schizophrenia | 10.1016/j.euroneuro.2010.08.008 | Wrong outcomes |
| Ranglani 2023 | Testing for associations between HbA1c levels, polygenic risk and brain health in UK Biobank (N = 39 283) | 10.1111/dom.15207 | Wrong exposure |
| Rolls 2023 | Lifestyle risks associated with brain functional connectivity and structure | 10.1002/hbm.26225 | Wrong outcomes |
| Rovio 2022 | The effect of physical activity and other lifestyle factors on dementia, Alzheimer's disease and structural brain changes. |  | Wrong exposure |
| Rutherford 2015 | Investigating maternal brain structure and its relationship to substance use and motivational systems |  | Wrong exposure |
| Sami 2020 | Cannabis use in patients with early psychosis is associated with alterations in putamen and thalamic shape | 10.1002/hbm.25131 | Wrong exposure |
| Schlaepfer 2006 | Decreased frontal white-matter volume in chronic substance abuse | 10.1017/S1461145705005705 | Wrong method (not t-1 weighted MRI) |
| Schneider 2014 | Smoking status as a potential confounder in the study of brain structure in schizophrenia | 10.1016/j.jpsychires.2013.12.004 | Wrong exposure |
| Schnell 2012 | Increased gray matter density in patients with schizophrenia and cannabis use: A voxel-based morphometric study using DARTEL | 10.1016/j.schres.2012.03.021 | Wrong outcomes |
| Scott 2023 | Impact of Adolescent Cannabis Use on Neurocognitive and Brain Development | 10.1016/j.psc.2023.03.012 | Wrong study design |
| Scott 2023 | Impact of Adolescent Cannabis Use on Neurocognitive and Brain Development | 10.1016/j.chc.2022.06.002 | Wrong method (not t-1 weighted MRI) |
| Shen 2018 | Cerebellar gray matter reductions associate with decreased functional connectivity in nicotine-dependent individuals | 10.1093/ntr/ntx168 | Wrong outcomes |
| Shen 2019 | Interactions between monoamine oxidase A rs1137070 and smoking on brain structure and function in male smokers | 10.1111/ejn.14282 | Wrong outcomes |
| Shi 2019 | Study on the mechanism of brain damage based on structural covariant network to evaluate the brain structure of nicotine addicts | 10.3760/cma.j.issn.0376-2491.2019.09.007 | unable to obtain full text |
| Shi 2023 | Effects of current smoking severity on brain gray matter volume in opioid use disorder‚Äìa voxel-based morphometry study | 10.1080/00952990.2023.2169616 | Wrong comparator |
| Shollenbarger 2015 | Impact of cannabis use on prefrontal and parietal cortex gyrification and surface area in adolescents and emerging adults | 10.1016/j.dcn.2015.07.004 | Wrong outcomes |
| Soleimani 2023 | Altered brain structural and functional connectivity in cannabis users | 10.1038/s41598-023-32521-8 | Wrong outcomes |
| Soleimani 2023 | Cortical Morphology in Cannabis Use Disorder: Implications for Transcranial Direct Current Stimulation Treatment | 10.32598/bcn.2021.3400.1 | Wrong outcomes |
| Solowij 2011 | Cerebellar white-matter changes in cannabis users with and without schizophrenia | 10.1017/S003329171100050X | Overlapping samples |
| Solowij 2013 | Alteration to hippocampal shape in cannabis users with and without schizophrenia | 10.1016/j.schres.2012.10.040 | Overlapping samples |
| Srinivasa 2016 | Cardiovascular risk factors associated with smaller brain volumes in regions identified as early predictors of cognitive decline | 10.1148/radiol.2015142488 | Wrong exposure |
| Stoeckel 2016 | Lower gray matter density and functional connectivity in the anterior insula in smokers compared with never smokers | 10.1111/adb.12262 | Wrong outcomes |
| Stone 2012 | Substance use and regional gray matter volume in individuals at high risk of psychosis | 10.1016/j.euroneuro.2011.06.004 | Wrong exposure |
| Sullivan 2018 | The role of aging, drug dependence, and Hepatitis C comorbidity in alcoholism cortical compromise | 10.1001/jamapsychiatry.2018.0021 | Wrong exposure |
| Sullivan 2020 | Assessing the role of cannabis use on cortical surface structure in adolescents and young adults: Exploring gender and aerobic fitness as potential moderators | 10.3390/brainsci10020117 | Wrong outcomes |
| Sullivan 2021 | Cannabis Use and Brain Volume in Adolescent and Young Adult Cannabis Users: Effects Moderated by Sex and Aerobic Fitness | 10.1017/S135561772100062X | Wrong outcomes |
| Sultan 2021 | Neurostructural Correlates of Cannabis Use in Adolescent Bipolar Disorder | 10.1093/ijnp/pyaa077 | Wrong outcomes |
| Teipel 2016 | Association between Smoking and Cholinergic Basal Forebrain Volume in Healthy Aging and Prodromal and Dementia Stages of Alzheimer's Disease | 10.3233/JAD-151100 | Wrong exposure |
| Thames 2017 | Marijuana effects on changes in brain structure and cognitive function among HIV+ and HIV‚àí adults | 10.1016/j.drugalcdep.2016.11.007 | Wrong exposure |
| Thayer 2017 | Structural neuroimaging correlates of alcohol and cannabis use in adolescents and adults | 10.1111/add.13923 | Wrong exposure |
| Thayer 2020 | Recent tobacco use has widespread associations with adolescent white matter microstructure | 10.1016/j.addbeh.2019.106152 | Wrong outcomes |
| Topiwala 2022 | Alcohol consumption and MRI markers of brain structure and function: Cohort study of 25,378 UK Biobank participants | 10.1016/j.nicl.2022.103066 | Overlapping samples |
| Tregellas 2007 | Gray matter volume differences and the effects of smoking on gray matter in schizophrenia | 10.1016/j.schres.2007.08.019 | Wrong comparator |
| VanHaren 2013 | Confounders of excessive brain volume loss in schizophrenia | 10.1016/j.neubiorev.2012.09.006 | Wrong study design |
| Wade 2019 | Orbitofrontal cortex volume prospectively predicts cannabis and other substance use onset in adolescents | 10.1177/0269881119855971 | Wrong outcomes |
| Wallace 2024 | Neurite Orientation Dispersion and Density Imaging (NODDI) of Brain Microstructure in Adolescent Cannabis and Nicotine Use | 10.3390/bs14030231 | Wrong outcomes |
| Wang 2020 | Changes of functional connectivity in executive network during the resting state in adolescent smokers: a research based on independent component analysis | 10.3969/j.issn.1005-202X.2020.03.006 | Wrong outcomes |
| Wang 2021 | Cerebellar thickness changes associated with heavy cannabis use: A 3-year longitudinal study | 10.1111/adb.12931 | Wrong outcomes |
| Weidler 2024 | Resting-state functional connectivity and structural differences between smokers and healthy non-smokers | 10.1038/s41598-024-57510-3 | Wrong outcomes |
| Welch 2011 | The impact of substance use on brain structure in people at high risk of developing schizophrenia | 10.1093/schbul/sbq013 | Wrong exposure |
| Welch 2013 | Tensor-based morphometry of cannabis use on brain structure in individuals at elevated genetic risk of schizophrenia | 10.1017/S0033291712002668 | Wrong exposure |
| Weng 2022 | Assessment of brain connectome alterations in male chronic smokers using structural and generalized q-sampling MRI | 10.1007/s11682-022-00647-4 | Wrong outcomes |
| Wetherill 2015 | Cannabis, cigarettes, and their co-occurring use: Disentangling differences in gray matter volume | 10.1093/ijnp/pyv061 | Wrong outcomes |
| Whitsel 2022 | Long-term associations of cigarette smoking in early mid-life with predicted brain aging from mid- to late life | 10.1111/add.15710 | Wrong exposure |
| Wilson 2000 | Brain morphological changes and early marijuana use: A magnetic resonance and positron emission tomography study | 10.1300/J069v19n01_01 | Wrong exposure |
| Windle 2018 | Age sensitive associations of adolescent substance use with amygdalar, ventral striatum, and frontal volumes in young adulthood | 10.1016/j.drugalcdep.2018.02.007 | Wrong exposure |
| Wittemann 2021 | Cognition and Cortical Thickness in Heavy Cannabis Users | 10.1159/000509987 | Wrong outcomes |
| Wobrock 2009 | Comorbid substance abuse and brain morphology in recent-onset psychosis | 10.1007/s00406-008-0831-x | Wrong exposure |
| Wolf 2021 | Structural correlates of sensorimotor dysfunction in heavy cannabis users | 10.1111/adb.13032 | Wrong outcomes |
| Wu 2020 | Gray matter changes in chronic heavy cannabis users: A voxel-level study using multivariate pattern analysis approach | 10.1097/WNR.0000000000001532 | Wrong outcomes |
| Xiang 2023 | Association between vmPFC gray matter volume and smoking initiation in adolescents | 10.1038/s41467-023-40079-2 | Wrong exposure |
| Xie 2008 | Serial MR evaluation of brain atrophy in patients with cerebral vascular disease |  | unable to obtain full text |
| Xie 2024 | Altered dynamic functional connectivity of insular subdivisions among male cigarette smokers | 10.3389/fpsyt.2024.1353103 | Wrong outcomes |
| Xu 2024 | Abnormal developmental of structural covariance networks in young adults with heavy cannabis use: a 3-year follow-up study | 10.1038/s41398-024-02764-8 | Wrong outcomes |
| Y√ºcel 2006 | Structural brain correlates of alcohol and cannabis use in recreational users | 10.1111/j.1601-5215.2006.00154.x | Wrong exposure |
| Yang 2023 | An augmented Mendelian randomization approach provides causality of brain imaging features on complex traits in a single biobankscale dataset | 10.1371/journal.pgen.1011112 | Wrong outcomes |
| Yang 2023 | Assessing the interaction effects of brain structure longitudinal changes and life environmental factors on depression and anxiety | 10.1002/hbm.26153 | Wrong outcomes |
| Ye 2020 | Characterizing the Structural Pattern of Heavy Smokers Using Multivoxel Pattern Analysis | 10.3389/fpsyt.2020.607003 | Wrong outcomes |
| Ye 2021 | Characterizing the structural pattern of heavy smokers using multivoxel pattern analysis. | 10.3389/fpsyt.2020.607003 | Wrong outcomes |
| Yeh 2007 | Hierarchical linear modeling (HLM) of longitudinal brain structural and cognitive changes in alcohol-dependent individuals during sobriety | 10.1016/j.drugalcdep.2007.05.027 | Wrong exposure |
| Yeshaw 2024 | Uncovering Predictors of Low Hippocampal Volume: Evidence from a Large-Scale Machine-Learning-Based Study in the UK Biobank | 10.1159/000538565 | Wrong outcomes |
| Yokoyama 2018 | Additive Effect of Cigarette Smoking on Gray Matter Abnormalities in Schizophrenia | 10.1093/schbul/sbx092 | Wrong outcomes |
| Yu 2011 | Regional grey and white matter changes in heavy male smokers | 10.1371/journal.pone.0027440 | Wrong outcomes |
| Yu 2020 | Cannabis-Associated Psychotic-like Experiences Are Mediated by Developmental Changes in the Parahippocampal Gyrus | 10.1016/j.jaac.2019.05.034 | Wrong exposure |
| Yucel 2008 | Regional brain abnormalities associated with long-term heavy cannabis use | 10.1001/archpsyc.65.6.694 | Overlapping samples |
| Yucel 2016 | Hippocampal harms, protection and recovery following regular cannabis use | 10.1038/TP.2015.201 | Overlapping samples |
| Zanchi 2015 | Cigarette smoking leads to persistent and dose-dependent alterations of brain activity and connectivity in anterior insula and anterior cingulate | 10.1111/adb.12292 | Wrong outcomes |
| Zhang 2011 | Factors underlying prefrontal and insula structural alterations in smokers | 10.1016/j.neuroimage.2010.08.008 | Wrong outcomes |
| Zhang 2011 | Anatomical differences and network characteristics underlying smoking cue reactivity | 10.1016/j.neuroimage.2010.07.063 | Overlapping samples |
| Zhang 2017 | Structural changes of brain gray matter in male long-term smokers under magnetic resonance imaging | 10.3760/cma.j.issn.0376-2491.2017.45.010 | unable to obtain full text |
| Zhang 2023 | Distinct resting-state functional connectivity patterns of Anterior Insula affected by smoking in mild cognitive impairment | 10.1007/s11682-023-00766-6 | Wrong outcomes |
| Zhang 2023 | Integrative brain structural and molecular analyses of interaction between tobacco use disorder and overweight among male adults | 10.1002/jnr.25141 | Wrong outcomes |
| Zhang 2024 | Analysis of the interactive effects between smoking addiction and overweight on brain gray matter volume | 10.3760/cma.j.cn113661-20230728-00018 | Wrong outcomes |
| Zhou 2023 | The changes of intrinsic connectivity contrast in young smokers | 10.1111/adb.13347 | Wrong outcomes |
| Zhu 2021 | Polygenic Risk for Schizophrenia, Brain Structure, and Environmental Risk in UK Biobank | 10.1093/schizbullopen/sgab042 | Wrong exposure |
| Zivadinov 2009 | Smoking is associated with increased lesion volumes and brain atrophy in multiple sclerosis | 10.1212/WNL.0b013e3181b2a706 | Wrong outcomes |
| Zorlu 2017 | Effects of cigarette smoking on cortical thickness in major depressive disorder | 10.1016/j.jpsychires.2016.09.009 | Wrong outcomes |
